# AAV9 Gene Therapy in GM1 Gangliosidosis Type II: A Phase 1/2 Trial

**DOI:** 10.1101/2025.07.28.25332074

**Authors:** Connor J. Lewis, Precilla D’Souza, Jean M. Johnston, Maria T. Acosta, Cristan Farmer, Eva H. Baker, Anna Crowell, Yoliann Mojica, Sumaiya Rahman, Lisa Joseph, Adam Hartman, Gilbert Vézina, Zenaide Quezado, Muhammad H. Yousef, Amelia Luckett, Zeynep Vardar, Mohammad Salman Shazeeb, Manuela Corti, Meghan Blackwood, Kirsten Coleman, Audrey Thurm, Erika De Boever, William A. Gahl, Barry J. Byrne, Terence R. Flotte, Xuntian Jiang, Amanda L. Gross, Allison M. Keeler, Heather Gray-Edwards, Douglas R. Martin, Miguel Sena-Esteves, Cynthia J. Tifft

## Abstract

**Background:** GM1 gangliosidosis, caused by biallelic variants in *GLB1*, results from deficiency of lysosomal β-galactosidase, the enzyme primarily responsible for degradation of GM1 ganglioside. This progressive neurodegenerative disease is uniformly fatal with no approved therapies, but preclinical studies utilizing gene therapy have shown promising results.

**Methods:** This phase 1-2 open label dose escalation study utilized a single intravenous administration of adeno-associated virus serotype 9 (AAV9) encoding β-galactosidase in the first nine enrolled Type II GM1 gangliosidosis participants. The primary endpoint was safety after 3 years; secondary and exploratory efficacy outcomes included cerebrospinal fluid (CSF) GM1 ganglioside and β-galactosidase, clinical assessments, and neuroimaging changes.

**Results:** One serious adverse event was attributed to the vector, i.e., vomiting requiring rehospitalization for IV hydration. Serum aspartate and alanine aminotransferase levels increased following gene transfer but returned to baseline by 18 months. Per protocol analysis found stability in Vineland Expressive Communication and Gross Motor and significant declines in Fine Motor and Receptive Communication. Median CGI-Improvement scores at 2 and 3 years after gene transfer were “minimally improved” or “no change”. CSF β-galactosidase increased and GM1 ganglioside decreased in all participants. MRI showed improved myelination by differential tractography and declines in cerebral atrophy. MRS showed reduced loss of *N*-acetylaspartate+*N*-acetylaspartyl glutamate (NAA) compared with historical controls.

**Conclusions:** A single IV infusion of AAV9 encoding β-galactosidase was well-tolerated among the first nine Type II GM1 gangliosidosis participants. Secondary and exploratory outcomes suggested improvements in biochemical markers and neuroimaging and stabilized or reduced rates of developmental deterioration (NCT03952637).

## INTRODUCTION

GM1 gangliosidosis (GM1) is a rare, progressive neurodegenerative lysosomal storage disorder caused by biallelic variants in *GLB1* encoding β-galactosidase, an enzyme that catabolizes GM1 ganglioside.^1^ Because neurons have a high content of GM1 ganglioside, the clinical manifestations of GM1 largely involve the central nervous system (CNS). Type I GM1 is the most severe subtype and Type III is the mildest. Type II GM1 patients present with developmental delays, ataxia, dystonia, dysarthria, low bone mineral density, and progressive brain atrophy.^2–5^ Two Type II GM1 subtypes, late-infantile and juvenile, have different clinical trajectories, but all patients experience a progressive, uniformly fatal course.^6,7^

There are no approved therapies for GM1, making gene replacement attractive, especially since preclinical studies of intravenous adeno-associated virus serotype 9 (AAV9) gene therapy in a feline GM1 model resembling Type II disease demonstrated increased survival, repletion of cerebrospinal fluid (CSF) enzyme levels, and correction of magnetic resonance imaging (MRI) and magnetic resonance spectroscopy (MRS) abnormalities.^8,9^ We performed a clinical trial of *GLB1* delivery utilizing AAV9 in 12 children with Type II GM1 (NCT03952637).

## METHODS

### Study Design

This 3-year phase 1-2 open label dose escalation study had 2 years of additional follow-up. Eligible children were age 6 months - 12 years with biallelic *GLB1* variants, β-galactosidase deficiency, a Type II GM1 gangliosidosis phenotype, and a Vineland-3 Adaptative Behavior Composite standard score ≥ 40 (Appendix A). Individuals were excluded if their anti-AAV9 titer was > 1:50. All participants received AAV9-GLB1 (1.5×10^13^ or 4.5×10^13^ vector genomes/kg) intravenously 1 mL/min (Appendix B). Immune suppression was achieved with rapamycin and Rituximab 3 weeks before gene transfer, methylprednisolone 1-2 hours before gene transfer, and oral prednisone following gene therapy (Appendix C). Blood draws, lumbar punctures, Clinical Global Impression Improvement (CGI-I) scores, Vineland-3 Adaptive Behavior Scales, and MRI and MRS images were obtained periodically (Appendicies D, E, and F).

### End Points

The primary end point was safety at 3 years post-infusion, i.e., adverse events (AE) or serious adverse events (SAE). Key secondary outcomes included the CGI-Improvement (CGI-I) rating and Vineland Adaptive Behavior Scales (Appendix E) at 2 years post-infusion, changes in brain MRI (volumetric analysis, diffusion tensor imaging [DTI]), and MRS (Appendix F). Exploratory endpoints included CSF GM1 ganglioside, CSF and serum β-galactosidase activity, and H3N2b (a unique oligosaccharide in GM1) in CSF, plasma, and urine (Appendix D).^10^

### Statistical Analysis

For this rare condition, the planned sample size ceiling was based on feasibility; no a priori power calculation was performed. The statistical analysis plan dictated an intent-to-treat mixed model for repeated measures controlling for baseline age to evaluate change from baseline on the Vineland GSV at Year 2; the Year 3 timepoint was also included (Appendix E). Alpha was set at 5%, uncorrected for multiplicity. Longitudinal biochemical and neuroimaging data were descriptive. Post-hoc exploratory analysis of safety and biochemical outcomes was performed by dosage group and analysis of developmental and neuroimaging by subtype due to known differential clinical trajectories.^5,6,11,12^

## Results

### Study Population; Baseline Values (Table 1)

Nine children (5 males) were enrolled between August 19, 2019 and October 24, 2022; at the cutoff date for this interim analysis (June 1, 2025), GT17 had not reached 3 years of follow-up. Five participants (four late-infantile) received low dose and four (all juvenile) received high dose in order of enrollment. Individual Vineland and CGI scores reflected marked GM1 disease at baseline in most participants (Table 1). The mean CSF β-galactosidase activity ranged from 0-10% of normal and the mean CSF GM1 ganglioside levels were 2-5-fold normal. Baseline AST values ranged from 39-146 U/L (normal, ≤41), while ALT and GGT levels were normal. All participants had an AAV9 titer ≤ 1:50.

**Table 1.** Baseline Patient Characteristics and Vector Dosage. Specific ages were redacted per MedArXiv requirements.

| Participant | GT04 | GT06 | GT07 | GT08 | GT03 | GT10 | GT11 | GT12 | GT17 |
| --- | --- | --- | --- | --- | --- | --- | --- | --- | --- |
| Sex | M | M | F | F | M | F | F | M | M |
| Age (yr) | 0-5 | 6-10 | 0-5 | 0-5 | 0-5 | 6-10 | 0-5 | 6-10 | 6-10 |
| Weight (kg) | 17.9 | 17.0 | 14.9 | 14.4 | 18.3 | 22.9 | 21.7 | 13.0 | 20.1 |
| Disease Subtype | Late-Infantile | Late-Infantile | Late-Infantile | Late-Infantile | Juvenile | Juvenile | Juvenile | Juvenile | Juvenile |
| Attained Running* | No | No | No | No | Yes | Yes | Yes | Yes | Yes |
| Attained 3 Word Sentence<sup>τ</sup> | No<sup>ω</sup> | No | No | No | Yes | Yes | Yes | Yes | Yes |
| Baseline Vineland<sup>†</sup> | 52 | 63 | 73 | 76 | 81 | 76 | 80 | 79 | 73 |
| Baseline CGI-S<sup>‡</sup> | 5 | 4 | 4 | 4 | 2 | 3 | 3 | 3 | 3 |
| β-galactosidase activity (fold normal)<sup>Ω</sup> | 0.00 | 0.01 | 0.00 | 0.04 | 0.07 | 0.06 | 0.01 | 0.00 | 0.10 |
| GM1 ganglioside levels (ng/mL)<sup>Ω</sup> | 175 | 75.8 | 163 | 172 | 155 | 128 | 67.4 | 97.6 | 125 |
| AST (U/L)<sup>υ</sup> | 146 | 63 | 55 | 57 | 71 | 39 | 51 | 70 | 64 |
| ALT (U/L)<sup>υ</sup> | 25 | 11 | 9 | 10 | 19 | 13 | 19 | 12 | 18 |
| GGT (U/L)<sup>υ</sup> | 14 | 8 | 10 | 8 | 14 | 14 | 14 | 13 | 14 |
| Baseline AAV9 Titer | 1:50 | < 1:25 | < 1:25 | < 1:25 | < 1:25 | < 1:25 | < 1:25 | < 1:25 | 1:50 |
| IVGT Dose<sup>¶</sup> | Low | Low | Low | Low | Low | High | High | High | High |

\*Ability to run assessed at baseline assessment. Neurotypical children begin running between 18- and 24-months.
<sup>τ</sup>Ability to form a 3-word sentence assessed at baseline assessment. Neurotypical children formulating 3-word sentences between 24- and 36-months.
<sup>ω</sup>GT04 was nonverbal at his baseline evaluation, indicating he had already missed the developmental milestone.
<sup>†</sup>Vineland scored between 36 and 7 days prior to AAV9-GLB1 administration.
<sup>‡</sup>CGI-S scored between 28 and 21 days prior to AAV9-GLB1 administration.
<sup>Ω</sup>Baseline GM1 and β-galactosidase were taken from CSF between 30 and 15 days prior to AAV9-GLB1 administration. Normal β-galactosidase activity was calculated from 10 pediatric controls as described in Section C of the Supplementary Appendix.
<sup>υ</sup>Baseline AST (upper limit of normal is 41 U/L), ALT (upper limit of normal is 30), and GGT (upper limit of normal is 16) were taken at 28 days prior to treatment.
<sup>¶</sup>The Low Dose Cohort received $1.5 \times 10^{13}$ vg/kg and the high dose cohort received $4.5 \times 10^{13}$ vg/kg.

### Safety

One SAE (GT17) involved vomiting requiring hospitalization for IV fluids on day 3; this was definitely related to the treatment. Four other SAEs (Supplement Table G1) were considered unrelated to AAV9-GLB1, with two attributable to GM1 disease progression, one to the immunosuppression regimen, and one to protocol-related procedures.

There were 113 other AEs (Supplement Table G2); 72 were Grade 1, 28 Grade 2, and 13 Grade 3. Thirty AEs were considered possibly-, probably-, or definitely-related to the vector (Table 2). There were 8 gastrointestinal events potentially related to the vector, including vomiting (5), retching (1), and reduced appetite (2). One instance of tachycardia was possibly related to the vector.

**Table 2:** Adverse Events during treatment possibly, probably, or definitely related to the gene therapy treatment.

| Body System<sup>a</sup><br>Adverse Event<sup>b</sup> | Low-Dose<br>(N) | High-Dose<br>(N) | Total<br>(N) | Relatedness to gene therapy<sup>c</sup> |  |  |
| --- | --- | --- | --- | --- | --- | --- |
|  |  |  |  | Possibly<br>(N) | Probably<br>(N) | Definitely<br>(N) |
| Cardiac disorders |  |  | 1 |  |  |  |
| Tachycardia |  | 1 | 1 | 1 |  |  |
| Gastrointestinal disorders |  |  | 8 |  |  |  |
| Vomiting |  | 5* | 5 |  | 3 | 2 |
| Retching |  | 1 | 1 |  | 1 |  |
| Decreased appetite |  | 2 | 2 |  | 2 |  |
| Laboratory abnormalities |  |  | 21 |  |  |  |
| Aspartate aminotransferase increased | 4 | 5 | 9 | 5 | 3 | 1 |
| Fibrin D Dimer increased | 2 | 3 | 5 |  | 4 | 1 |
| Serum ferritin increased |  | 3 | 3 |  | 2 | 1 |
| C-reactive protein increased |  | 1 | 1 |  |  | 1 |
| Alanine aminotransferase increased |  | 2 | 2 | 1 | 1 |  |
| Positive Elispot |  | 1 | 1 |  |  | 1 |

<sup>a</sup> Preferred Term
<sup>b</sup> Common Terminology Criteria for Adverse Events (CTCAE, v5.0)
<sup>c</sup> Based on investigator assessment
\*Serious Adverse Event

There were 21 laboratory abnormalities related to the vector, including one elevated Elispot that resolved. Other AEs were elevated aspartate aminotransferase (AST, 9), D-dimer (5), ferritin (3), alanine aminotransferase (ALT, 2), or C-reactive protein (1). All 9 participants had elevated AST levels at baseline (Table 1), typical for the underlying disease. Mean AST levels were further elevated following gene transfer; elevations were similar between the low and high dose participants (Figure 1A). All AST levels fell to baseline 18 months after treatment (Supplementary Figure H1). ALT levels also increased post AAV9-GLB1 infusion but were generally resolved between week 52 and 78 following gene transfer (Supplement Figure H2). Gamma glutamyl transferase (GGT) levels were well controlled except in GT03, whose central catheter became infected on day 18 (Supplement Figure H3). Viral genomes were detected in saliva, urine, and feces following AAV9-GLB1 administration; all returned to low levels by day 30 (Supplement Figures I1-I3).

**Figure 1.**
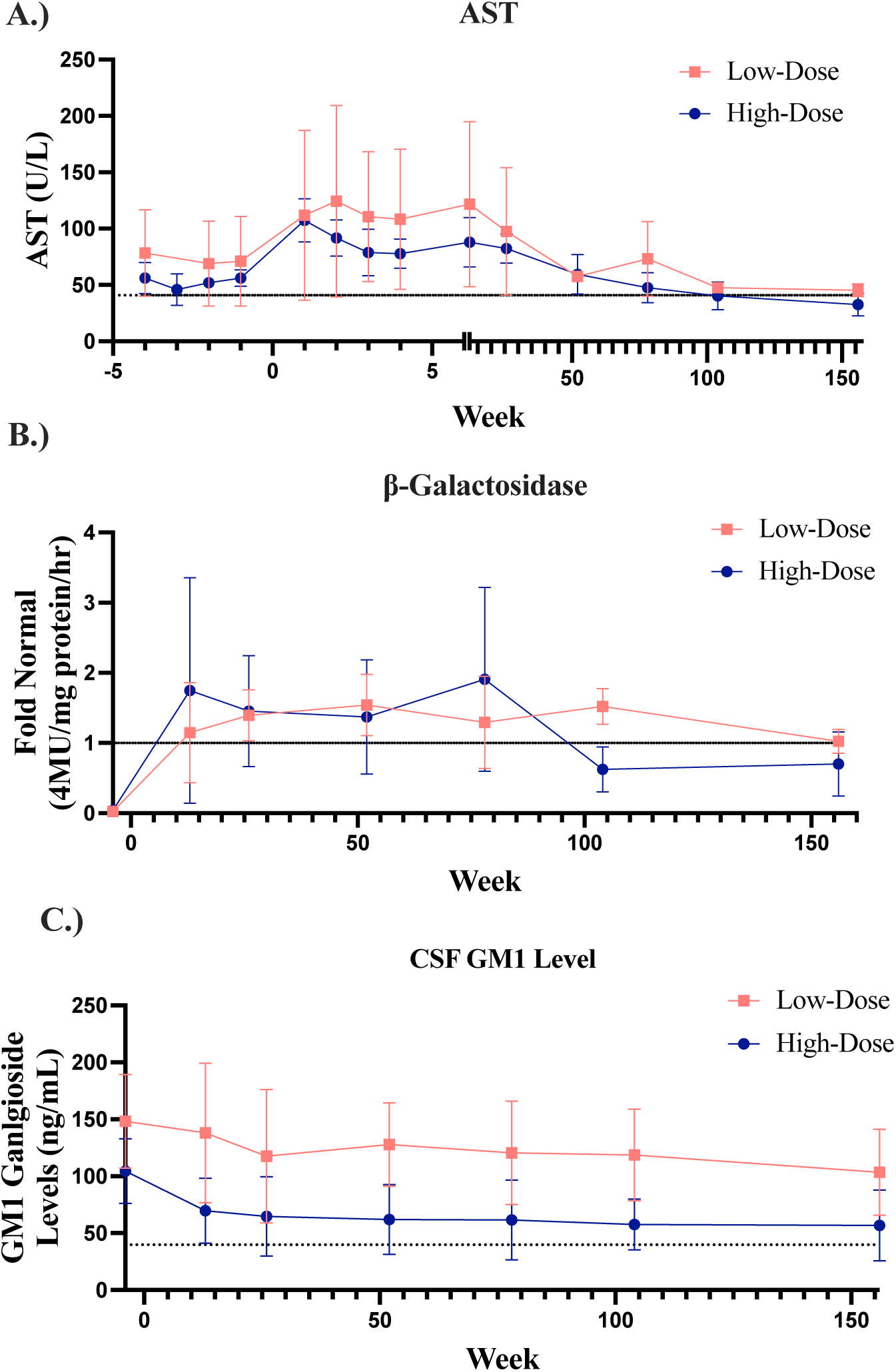
Biochemical Outcomes. **A.** AST levels in participants after receiving low dose or high dose gene transfer (mean +/− SD). **B.** CSF β-galactosidase levels after low dose or high dose gene transfer. **C.** CSF GM1 levels after gene transfer. The dotted line represents the upper limit of normal for AST (41 U/L) and GM1 ganglioside levels in CSF (39.9 ng/mL at 1 standard deviation above normal),^8^ and represents the normal activity from pediatric controls for β-galactosidase (*n*=9 up to Week 78, *n*=8 up to Week 104 (Year 2), *n*=7 at Week 156 (Year 3).

### Immune Responses to AAV9-GLB1

Complement (C3 and C4) was not depleted post gene transfer (Supplement Figures H4 and H5). Platelets declined slightly but remained above 100,000/mcL and returned to normal (Supplement Figure H6). There were transient increases in D-dimer (Supplement Figure H7). Anti-vector IgG typically peaked at 6-9 months (Supplemental Figure J1).

Anti-AAV9 IgM generally peaked 2-10 weeks post gene transfer (Supplemental Figure J2) and returned to baseline. Although Rituximab depleted circulating B-cells for at least 6 months, all participants developed anti-AAV9 neutralizing antibodies within 2 weeks after gene transfer, with an increase at 6 months and then stabilization for years (Supplement Figure J3).

Capsid-specific IFN-γ producing cells peaked in the first 10 weeks post treatment (Supplement Figure J4), corresponding to ALT and AST increases in most participants. Secondary capsid-specific IFN-γ immune responses occurred in GT06, GT08, GT10, and GT17 at 6 months to 1 year post AAV delivery and did not correspond to increases in ALT and AST. Increases in capsid-specific IFN-γ in GT10 and GT17 corresponded to loss of enzymatic activity in the serum but not in the CSF (Supplement Figure K3). Transgene-specific immune responses were not observed in any participant at any timepoint (Supplement Figure J5).

### Efficacy

#### Clinical Outcome Assessments

##### Vineland

Vineland scores of untreated GM1 participants decline with age.^11^ At years 2 and 3 (*n*=8), most gene transfer-treated participants’ Vineland-3 GSVs were not statistically different from their baseline scores (Figure 2A). However, the per protocol group-level analysis indicated statistically significant mean loss of skills in Fine Motor (Year 2: *t*(39) = −2.81, *p* = 0.008; Year 3: *t*(39) = −3.43, *p* = 0.001) and Receptive Communication (Year 2: *t*(39) = −2.15, *p* = 0.038; Year 3: n.s.) (Figure 2B). No significant changes in Expressive Communication and Gross Motor were observed. Individual Vineland Adaptive Behavior Growth Scale Values are shown in Supplementary Figures L1-L4.

**Figure 2.**
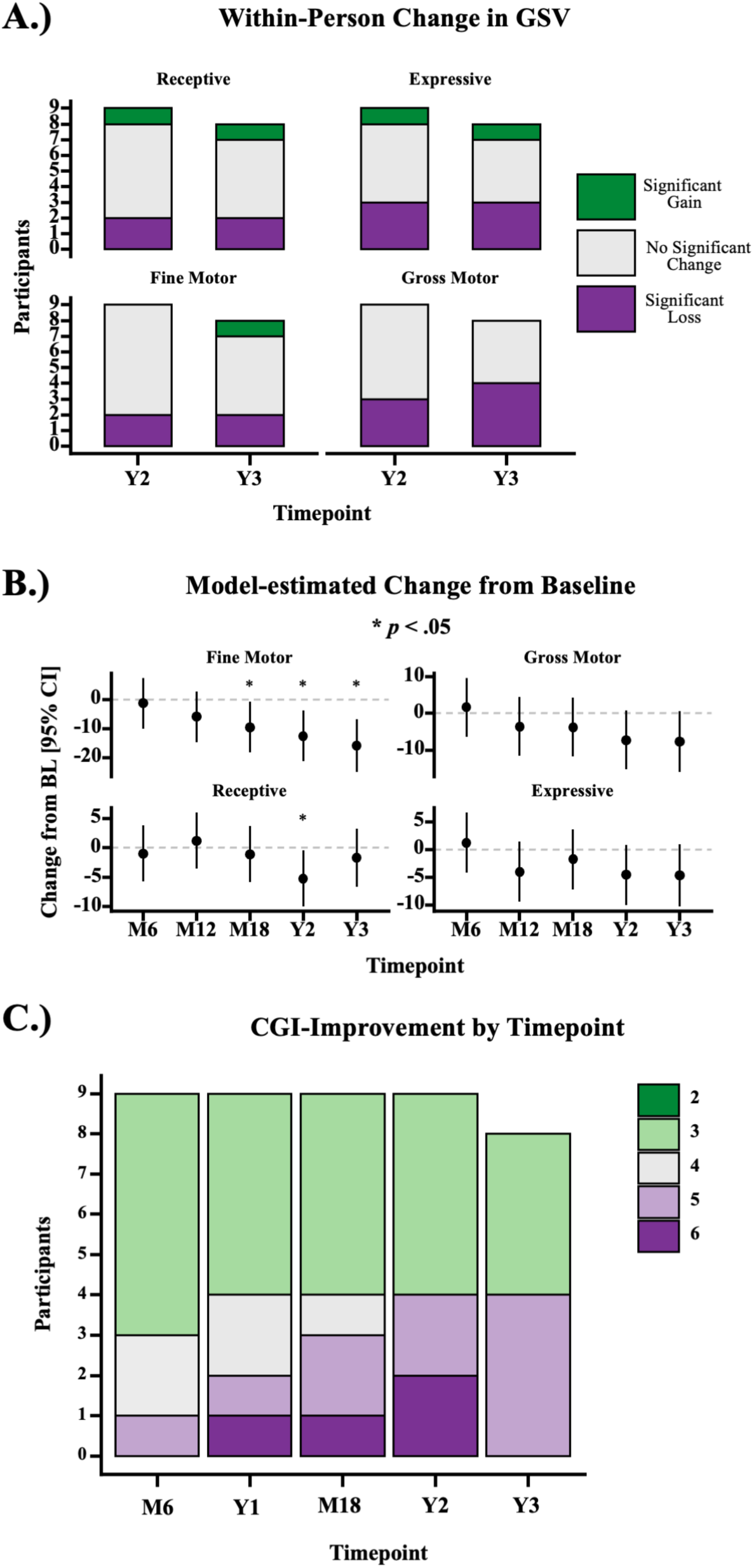
Clinical Outcome Assessments. **A.** N-of-1 analysis of change in Vineland-3 GSVs. The number of participants with significant (*p*<0.05) increases, decreases, or no significant change in GSV, based on the conditional standard error of measurement is shown by timepoint. **B.** Estimated marginal mean change from baseline at each timepoint, from the per protocol mixed model for repeated measures. Asterisks indicate statistically significant (*p*<0.05) change. **C.** The number of participants with each CGI-Improvement rating is shown by timepoint (*n*=9 up to Year 2, *n*=8 at Year 3).

##### CGI-Improvement

In untreated GM1, the mean annualized change in CGI-I scores of late-infantile participants was + 0.61 (SE=0.07) and of juvenile GM1 participants + 0.17 (SE=0.11),^12^ indicating worsening function. In the 9 treated participants, the median CGI-I at Year 2 was 3 (“Minimally Improved”) [IQR = 3, 5] and at Year 3 (*n*=8) was 4 (“No Change”) [IQR = 3, 5] (Figure 2C). Individual participant scores are shown in Supplementary Figure L3.

#### Enzyme repletion

Mean CSF β-galactosidase levels increased from ∼zero at baseline to approximately normal values for participants receiving either low or high dose gene transfer (**Figure 1B**). Mean CSF GM1 fell below baseline in both groups (**Figure 1C**).

Within 13-26 weeks of receiving low-dose AAV9-GLB1, all 4 late-infantile subjects showed normal CSF β-galactosidase levels; these remained above normal at the efficacy end point or the most recent evaluation (Supplement Figure K1). CSF GM1 ganglioside levels fell below baseline in all 4 participants at both one and two years post gene transfer (Supplement Figure K1). Serum β-galactosidase activity increased, peaking at 13-weeks following gene transfer in all 4 late-infantile participants (Supplement Figure K2). CSF, serum, and urine H3N2b levels decreased in all 4 late-infantile participants following gene transfer (Supplement Figure K3-K5).

CSF β-galactosidase activity increased in all five juvenile participants (*n*=4 high dose), reaching ≥normal levels in at 13 weeks and declining 2 years post gene transfer (Supplement Figure K1). CSF GM1 decreased in all 5 juvenile participants and typically reached a minimum between 6 months and 1 year following gene transfer (Supplement Figure K1). Serum β-galactosidase activity increased in all 5 juvenile participants and was higher in participants receiving the higher dose (Supplement Figure K2). H3N2b levels in CSF, serum, and urine decreased in all 5 juvenile participants (Supplement Figure K3-K5).

#### Neuroimaging; Differential Tractography

Differential tractography has documented consistent declines in neuronal tract numbers and volume in untreated GM1.^13^ One example is provided by the image of the untreated older sibling of GT17 over a one year period (Figure 3A; year 1), showing widespread decrements (red).

**Figure 3.**
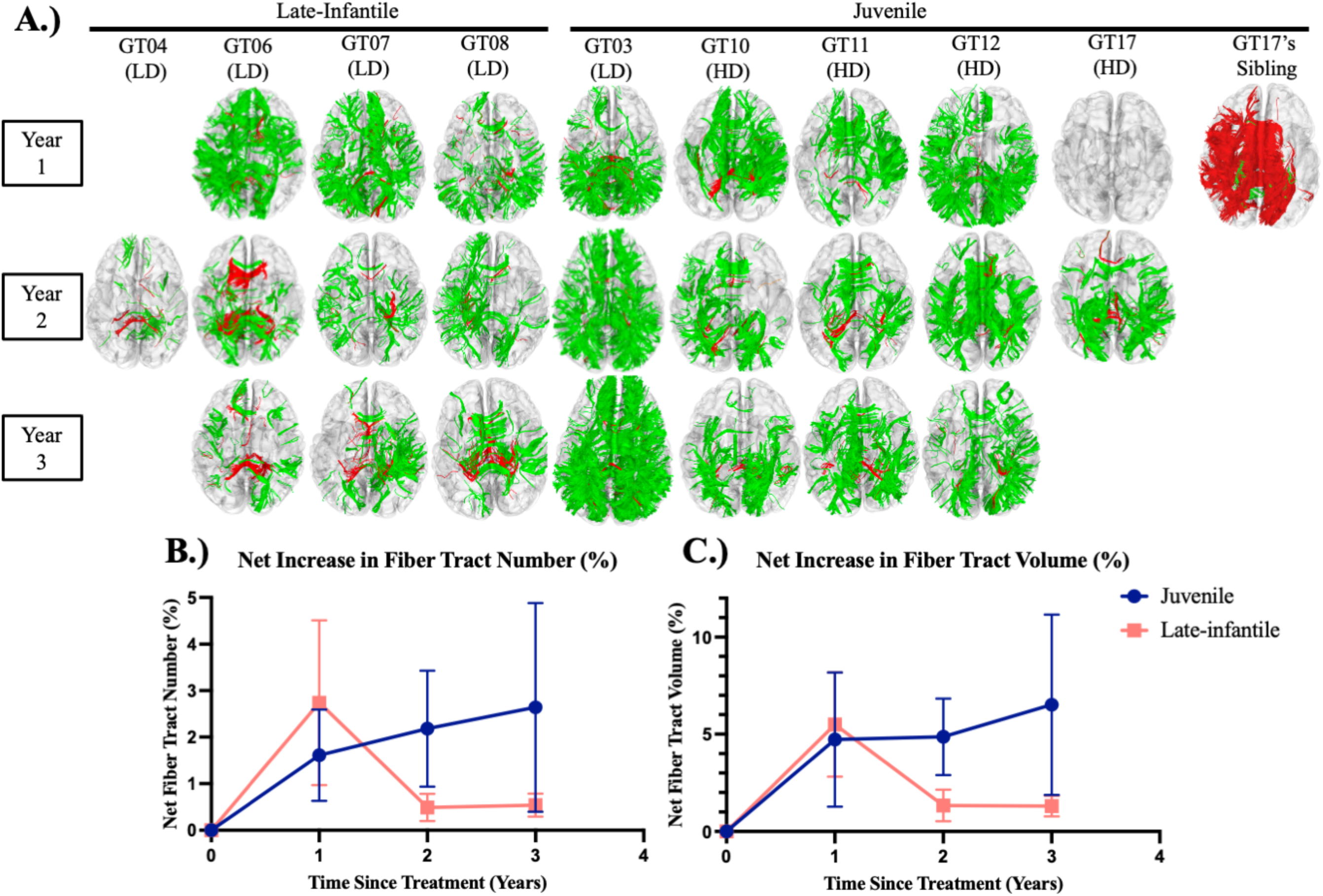
Neuroimaging Results. A.) Differential fiber tractography assessed fiber tract gains (green) and fiber tract losses (red) B.) Net fiber tract number in both cohorts. C.) Net fiber tract volume in both cohorts. LD = Low-Dose, HD = High-Dose (*n*=9 up to Year 2, *n*=8 at Year 3).

In contrast, all four late-infantile (low dose) GM1 participants showed gains in neuronal tracts 2 years post gene transfer, and GT06, GT07, and GT08 also had improvements at years 1 and 3 (Figure 3A). The greatest neuronal growth appeared in the first year after gene transfer. For the late-infantile subtype, differential tractography showed a mean net gain in fiber tract number of 2.7 ± 1.8 percent in the first year after gene transfer, falling to 0.5 ± 0.3 percent and 0.5 ± 0.2 percent by years 2 and 3 (Figure 3B). The mean net gain in fiber tract volume was 5.5 ± 2.7 percent, 1.3 ± 0.8 percent, and 1.3 ± 0.5 percent in years 1, 2, and 3 (Figure 3C). Neuroimaging of GT06, GT07, and GT08 showed reductions in the rate of global brain atrophy compared to the natural history (Section M of the Supplementary Appendix). All 4 late-infantile participants showed improvements in the rate of ventricle enlargement and thalamic atrophy compared to the natural history. GT07 and GT08 also showed reductions in the loss of NAA compared to historical controls, assessed by MRS (Supplement Figure M5).

Similar differential tractography results were obtained for the juvenile subtype (*n*=5; *n=*4 high dose) (Figure 3A). GT03, in particular, showed marked growth of neuronal tracts at years 1, 2, and 3. For the 5 GM1 juvenile subtype participants, differential tractography showed a mean net gain of 1.6 ± 1.0 percent, 2.2 ± 1.2 percent, and 2.6 ± 2.2 percent in fiber tract number at 1, 2, and 3 years after gene transfer (Figure 3B. The mean net gains in fiber tract volume were 4.7 ± 3.5 percent, 4.9 ± 2.0 percent, and 6.5 ± 4.6 percent in years 1, 2, and 3 (Figure 3C). GT03 and GT12 showed improvements in the rate of global brain atrophy compared to the natural history (Section M of the Supplementary Appendix). GT03, GT10, GT12, and GT17 showed improvements in the rate of ventricle enlargement compared to historical controls. GT03 and GT12 showed improvements in the rate of thalamic atrophy compared to the natural history. GT03, GT10, and GT12 also showed improvements in the loss of NAA compared to historical controls (Supplement Figure M5).

## DISCUSSION

GM1 gangliosidosis, a rare fatal monogenic disease with a well characterized natural history, represents a promising disorder for gene therapy.^11^ This phase 1-2 study provides evidence that a single intravenous infusion of AAV9 gene therapy carrying β-galactosidase is well-tolerated, and suggests that it increases β-galactosidase activity, decreases GM1 storage, slows brain atrophy, and allows for neuronal growth.

No substantial safety concerns arose from this clinical trial. Most adverse events appeared unrelated to AAV9-GLB1 administration and were moderate in severity. Only one serious adverse event was considered related to the vector. All participants were alive at the cutoff date. There was no evidence of thrombotic microangiopathy (TMA), hemophagocytic lymphohistiocytosis (HLH), or acute respiratory distress syndrome (ARDS), which have occurred during AAV therapeutics involving more than twice the viral load our high dose cohort received.^14–19^

AAV based gene therapies invoke immune responses,^20–24^ and all 9 participants in this report developed persistent neutralizing antibodies to the vector. Since Type II GM1 affects children, assessment of long-term transgene expression and innovative immunosuppression approaches are warranted for potential re-administration of the vector in the developing brain.^21,25^

Biochemical parameters provided preliminary evidence for efficacy. In the first nine participants, serum and CSF β-galactosidase levels increased above baseline levels and those of historical controls,^11^ reflecting *GLB1* transduction. Furthermore, reductions in the pentasaccharide biomarker, H3N2b, in urine, serum, and CSF, along with reductions in the cytotoxic substrate (GM1 ganglioside), indicated systemic increases in enzyme activity.^10^

Regarding development, the *a priori* hypothesis of improvement in Vineland-3 GSV was not supported. However, at Year 3, three of the four subdomains of interest remained not significantly different from baseline. The natural history of GM1 involves eventual declines among all affected individuals,^7,11^ so no change in 3 years may be a clinically meaningful outcome. Indeed, despite the statistically significant declines in some subdomains of the Vineland-3, the median CGI-I score was “minimally improved,” so some loss of skills may represent a clinically meaningful positive result.

Improvement in secondary radiologic outcomes corroborated the biochemical and clinical findings. Progressive brain atrophy with enlargement of the lateral ventricles is a neuroimaging hallmark of GM1 gangliosidosis;^5,11^ our gene therapy participants exhibited reduced rates of this atrophy, indicating structural CNS improvements. Differential tractography, reflecting myelination, supported this result, showing white matter trajectorial improvements in the AAV9-GLB1-treated individuals compared to historical controls.^13^

Baseline AAV9 antibody was higher in GT17 (1:50) than in other participants (< 1:25), and GT17 had a less beneficial response to the vector. In addition, GT04 was the youngest, most severely affected participant, with the highest GM1 ganglioside level, and was losing skills rapidly, as evidenced by baseline clinical outcome assessments. His poor response resembled that observed in metachromatic leukodystrophy where participants entering phases of rapid decline did not experience therapeutic benefit from gene therapy.^26^ In contrast, GT03, appeared pre-symptomatically in some areas due to the diagnosis in an older sibling, was the least severe at baseline and showed marked improvements in clinical outcomes and neuroimaging.

The apparent dose-related response to AAV9-GLB1 in terms of serum β-galactosidase activity was not observed in CSF enzyme activity, clinical outcome assessments, or neuroimaging. One possible explanation involves saturation of the blood brain barrier’s active transport of AAV9,^27^ while expression of *GLB1* from the liver could determine the dose-response in serum. The serum/CSF difference was not observed in our preclinical murine or feline models,^28,29^ and there are no human studies mentioning this phenomenon. Future investigations could involve the use of novel approaches for delivering AAV to the CNS, including administration into the cisterna magna, lateral ventricles, subarachnoid space, striatum, and the thalamus.^22,30,31^ Even more promising may be a new technology to target AAV capsids to the human transferrin receptor, greatly facilitating crossing of the blood brain barrier.^32^

Study limitations include its open-label design and clinical outcome assessment by unblinded clinicians. The small sample size prevented formal comparison of results between doses and/or subtype groups; further, subtype and dose groups were confounded. While the largest available natural history study^11^ is an important source of comparison, especially for CGI-I ratings,^12^ that study had older and more severely impacted participants than those in the current trial. Finally, 5 of the 9 participants were from 2 families (Table 1), perhaps limiting generalizibility.

This study provides preliminary evidence that administration of a single dose of AAV9-GLB1 is safe and well tolerated and suggests that it may lead to biochemical and neuroimaging improvements in Type II GM1 gangliosidosis. Future studies are needed to evaluate clinical benefits of earlier gene therapy and longer follow-up with respect to development as well as non-neurological manifestations of GM1, including its ophthalmologic, musculoskeletal, and hepatic symptoms.^11^ In addition, it will be important to determine the appropriate baseline anti-AAV antibody and clinical severity cutoff for AAV therapeutics in GM1 participants.

## Acknowledgements

We thank the participants and their families for the generosity of their time and efforts. We are also grateful to many staff members and care providers who contributed their expertise over the years.

## Data Availability

The data from this study is available from the corresponding author (CJT) at reasonable request upon completion of the study.

## Ethics Declaration

The Institutional Review Board at the NIH approved this protocol (19-HG-0101). Informed consent was completed with the parents and legal guardians of all participants. All paticipants were assessed in their ability to provide assent, and none we deemed capable.

## Supplementary Appendices

### List of Investigators

1 Audrey Thurm, Ph.D., Neurodevelopmental and Behavioral Phenotyping Service, National Institute of Mental Health, Bethesda, MD, USA

2 Barry J Byrne, M.D., Ph.D., Powell Gene Therapy Center, University of Florida

3 Terrence R. Flotte M.D. Ph.D., Department of Pediatrics, University of Massachusetts Chan Medical School

4 Xuntian Jiang, Ph.D., Department of Medicine, Washington University School of Medicine

5 Douglas R. Martin, Ph.D., Department of Anatomy, Physiology, & Pharmacology, Auburn, University College of Veterinary Medicine

6 Miguel Sena-Esteves Ph.D., Department of Neurology, University of Massachusetts Chan Medical School

7 Cynthia J. Tifft, M.D. Ph.D., Office of the Clinical Director and Medical Genetics Branch, National Human Genome Research Institute, National Institutes of Health

### Supplementary Methods

#### Supplement A Study Design and Inclusion/Exclusion Criteria

The study protocol (19-HG-0101; Included with Submission) was approved by the National Institutes of Health (NIH) institutional review board and U.S. Food and Drug Administration. Written informed consent was provided by the parents or legal guardian of each patient, none of whom was deemed capable of providing informed assent. The full inclusion/exclusion criteria are included below. Cohort one (low dose) received 1.5×10^13^ vg/kg, and cohort two (high dose) received 4.5×10^13^ vg/kg. Twelve pediatric participants were enrolled in the dose-ranging phase of this study (six in each cohort). However, only the results of the 9 children with at least 2 years of follow-up are included in this report; 3 participants were recently enrolled and have not completed the study.

For some analyses, Type II GM1 participants were further sub-typed into late-infantile (earlier onset of symptoms; never achieved running) and juvenile (able to run by age 2).^1–3^

##### Inclusion Criteria

- Vineland-3 Adaptive Behavior composite standard score greater than or equal to 40
- Male or female subjects ≥ 1 day old and < 12 years old at time of full ICF signing
- Biallelic variants in *GLB1*
- Documented deficiency of β-galactosidase enzyme by clinical laboratory testing
- Phenotype consistent with a diagnosis of Type II GM1 gangliosidosis, with symptom onset after the first year of life
- AAV9 antibody titers ≤1:50
- Agree to reside within 50 miles of the study site for at least 1 month following treatment

##### Exclusion Criteria

- AAV9 antibody titers >1:50
- Contraindications to concomitant medications
- Serious illness that would not allow travel to the study site
- Unwilling to undergo study interventions as outlined in the Schedule of Events
- Subjects receiving other unapproved, off-label or experimental therapies for GM1 gangliosidosis (e.g., miglustat, *N*-acetyl-leucine) within the last 60 days
- Any prior participation in a study in which a gene therapy vector or stem cell transplantation was administered
- Pregnant or lactating subjects
- Immunizations of any kind in the month prior to screening
- Evidence of cardiomyopathy on history, exam, or additional testing (echocardiogram or electrocardiogram) or other cardiac disease that in the opinion of the investigator would deem the subject unsafe to participate in the trial
- Indwelling ferromagnetic devices that would preclude MRI/fMRI/MRS imaging
- Ongoing medical condition that is deemed by the Principal Investigator to interfere with the conduct or assessments of the study
- History of infection with human immunodeficiency virus (HIV), hepatitis A, B, or C, or tuberculosis.
- History of or current chemotherapy, radiotherapy or other immunosuppressive therapy within the past 30 days. Corticosteroid treatment may be permitted at the discretion of the PI.
- Abnormal laboratory values considered clinically significant per the investigator
- Failure to thrive, defined as falling 20 percentiles (20/100) in body weight in the 3 months preceding Screening/Baseline
- Underlying defect in immune function
- History of multiple and severe life-threatening infections

#### Supplement B Vector Design and Preparation; Plasmid Sequence

##### Vector Design

As described in the protocol (attached), the AAV9-GLB1 therapy utilized in this study is a single stranded recombinant adeno-associated virus (AAV) vector encoding human lysosomal acid beta-galactosidase (Figure B1). The AAV9-GLB1 vector carries the human wild type *GLB1* open reading frame (Genbank: M34423.1) under control of the CAG promoter (CMV enhancer/chicken beta-actin promoter) followed by a polyadenylation signal derived from the simian 40 virus (SV40). The full plasmid sequence is provided below.

**Figure B1.**
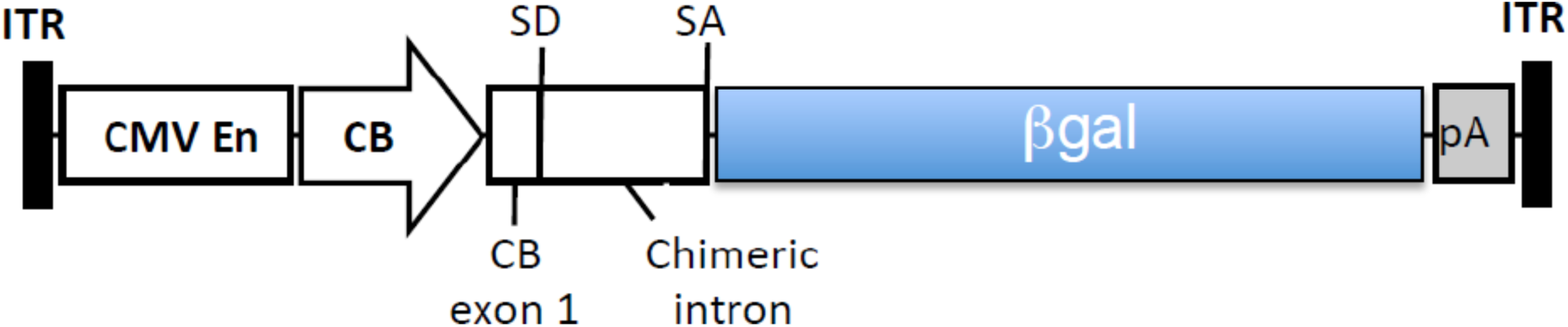
Schematic genome of AAV9-GLB1 vector

##### Vector Manufacturing

The AAV9-GLB1 vector was produced in the Clinical Manufacturing Facility at the Research Institute at Nationwide Children’s Hospital according to current good manufacturing practice (cGMP). The vector was produced by triple plasmid transient transfection of adherent HEK293 cells and purified by iodixanol gradient ultracentrifugation followed by ion-exchange column chromatography. The vector was formulated in 20 mM Tris (pH 8) 1 mM MgCl_2_, 200 mM NaCl containing 0.001% (w/v) poloxamer 188 with a final physical titer of 1.81×10^13^ vg/mL (digital droplet PCR), and an infectious unit titer of 3.6×10^9^ IU/mL (TCID_50_ assay). The full:empty capsid ratio was 26:1 (96.7% full) determined by analytical ultracentrifugation. The total protein content was 212.5 µg/mL with a total host cell protein content < 8 ng/mL and residual BSA < 20 ng/mL. AAV9-GLB1 vector was stored in 2.0 mL vials at < −60°C until use.

##### Vector Preparation on Infusion Day

The full details of the vector preparation are described in the pharmacy manual (included in this submission). After the work surface was prepped and appropriate protective gowning and protective measures donned, the study agent was thawed at room temperature and maintained at 2-8°C until delivered to the investigator. Then using a 30- or 50-mL syringe (depending on the total vector needed to administer the patient-specific total vg) and an 18-gauge needle, the necessary volume of vector was withdrawn. The syringe was then flipped to mix the contents, the needle removed and a 0.22 μm syringe filter attached to the syringe. The filter was then connected to a sterile line attached at the other end another equally sized syringe. The contents of the first syringe were transferred to the second syringe, filtering the solution. A small-bore extension set was then attached and primed. Finally, the syringe was labeled, placed on ice, and delivered to the investigator or study team.

##### Vector Delivery

As described in the protocol, participants received AAV9-GLB1 intravenously through a peripherally inserted central catheter (PICC) line, similar central line, or peripheral IV. The vector was administered at a rate of 1 mL/min using a standard syringe pump. The line was flushed after the vector was administered with 5 or 10 mL of saline at a rate of 1 mL/min.

##### Plasmid Sequence

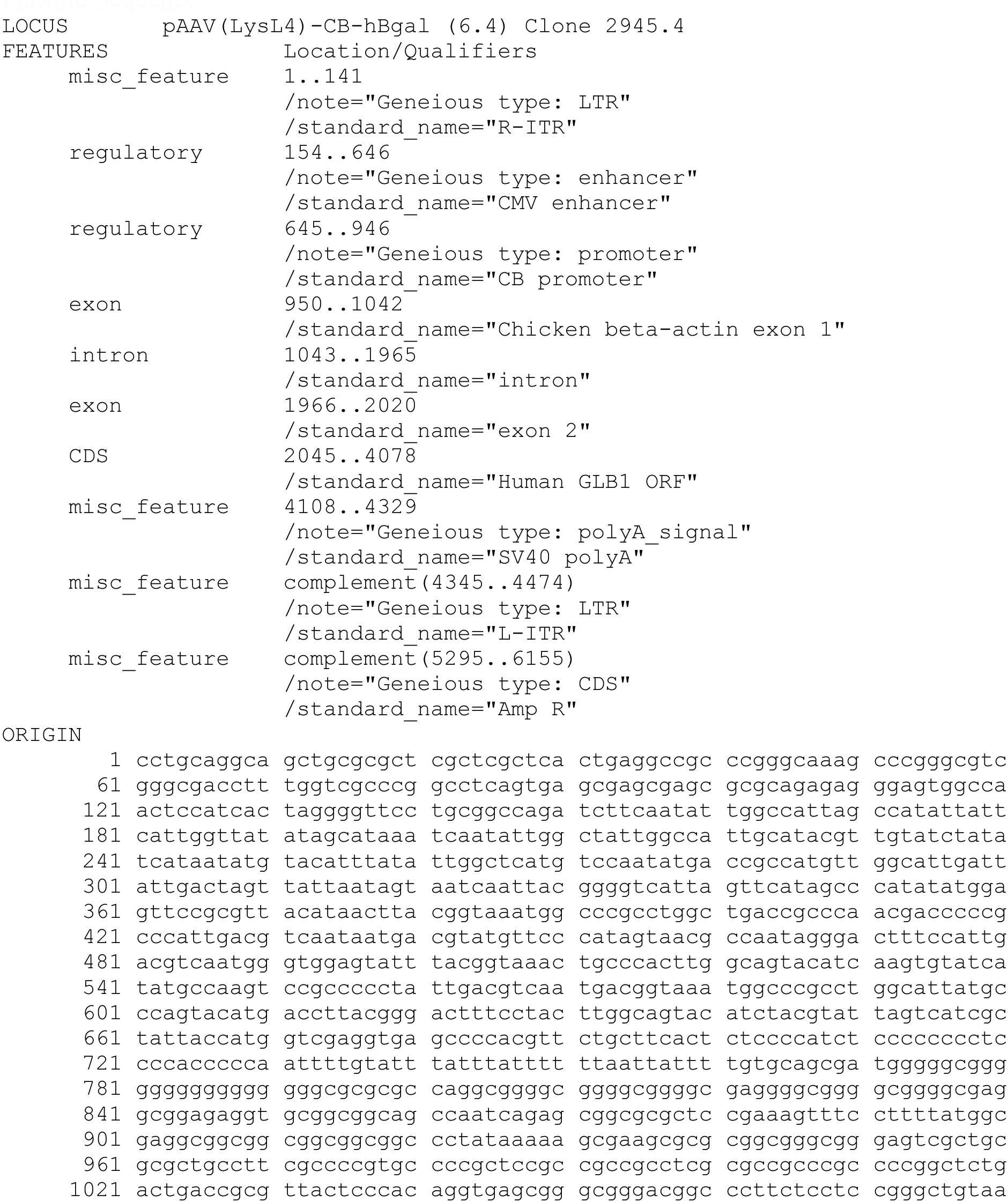
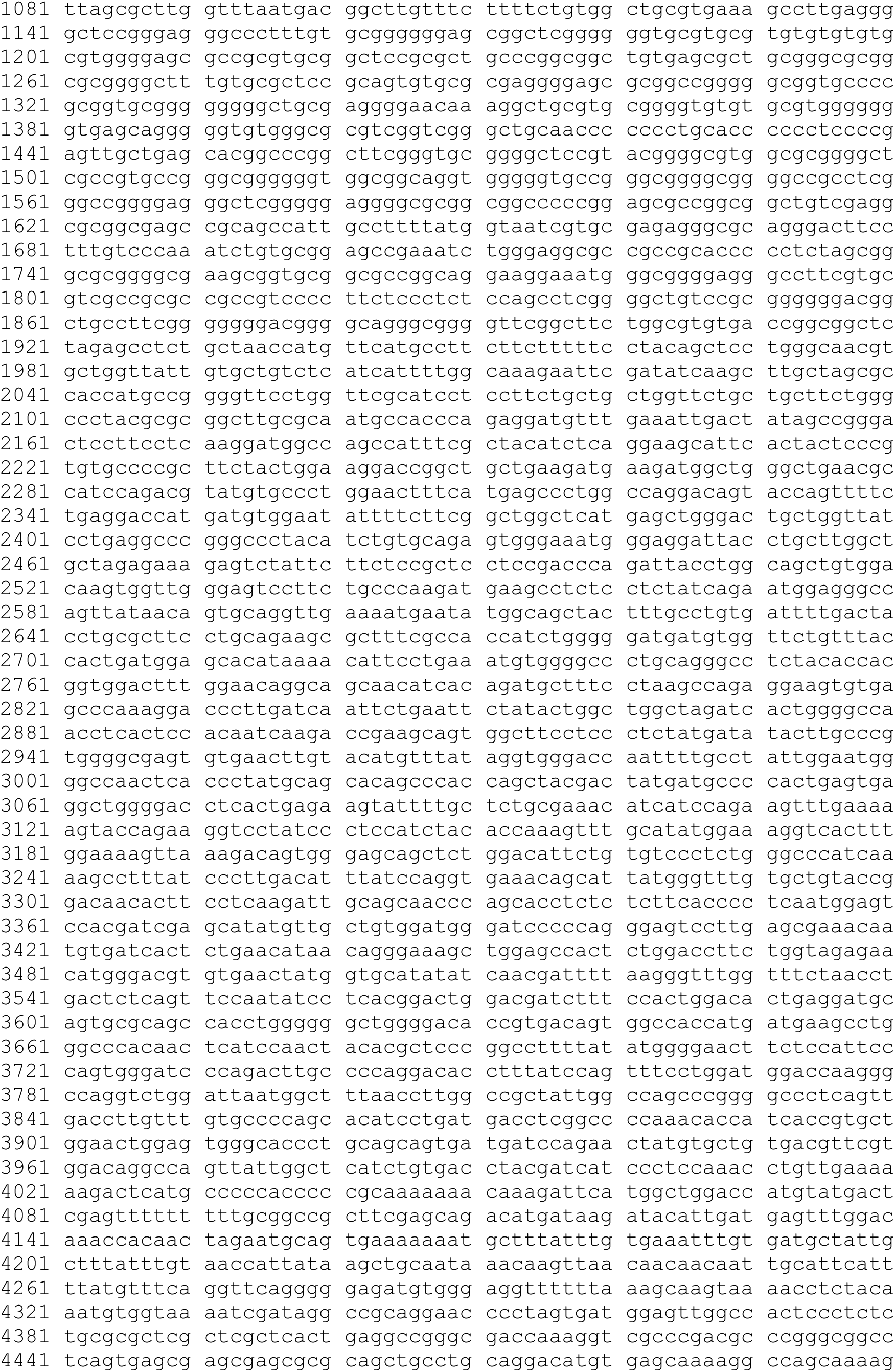
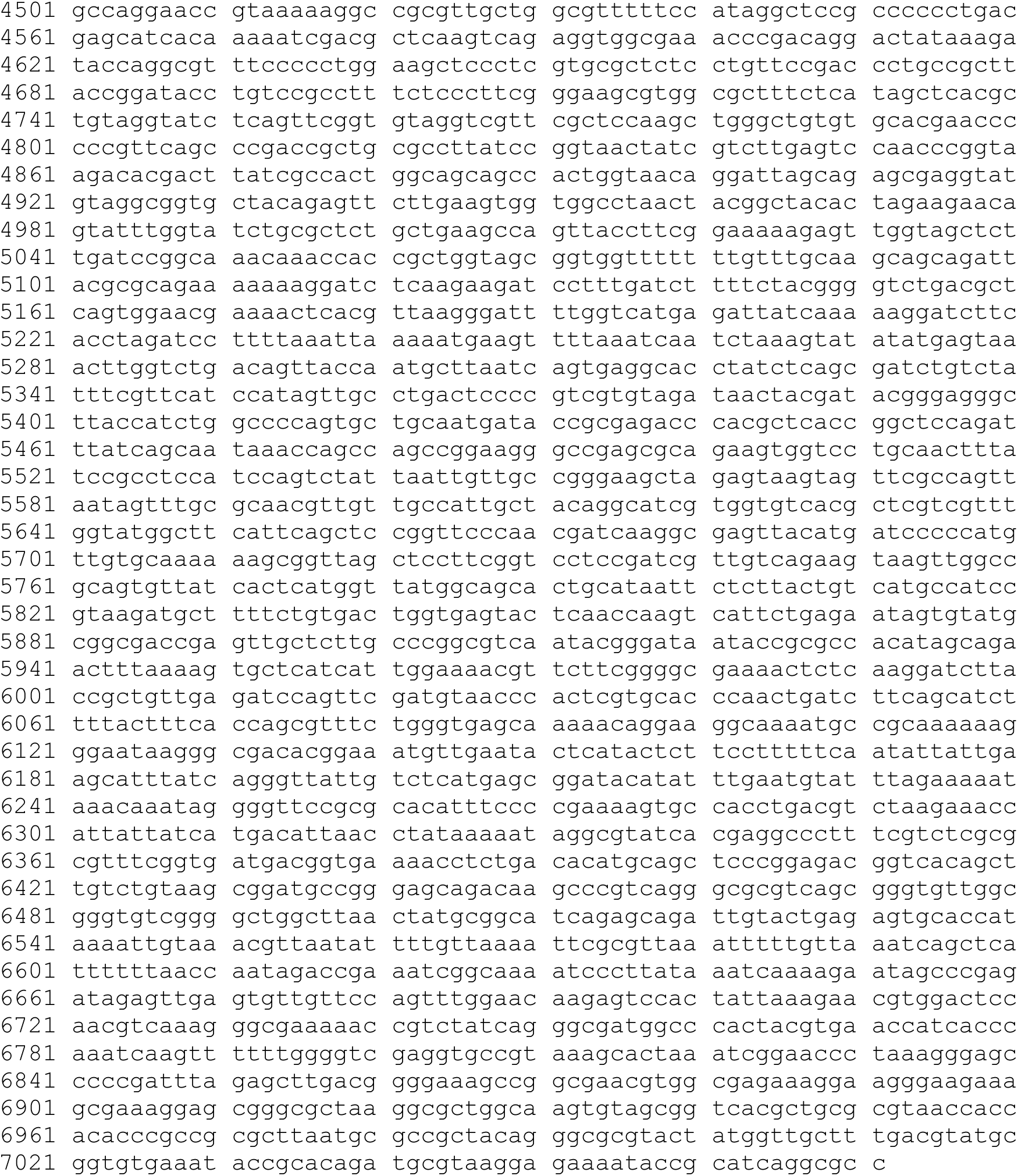

#### Supplement C Immunosuppression Treatment Regimen

The immunosuppression treatment schedule for NCT03952637 is displayed in Figure C1. Participants received daily rapamycin (0.5-1 mg/m^2^) beginning 3 weeks prior to AAV9-GLB1 administration until 6 months after administration; the dosage was adjusted to maintain a serum trough level of 7-12 ng/mL. Participants also received 3-4 doses (375 mg/m^2^) weekly of Rituximab starting 3 weeks prior to AAV9-GLB1 administration to deplete CD20^+^ B-cells. Participants received intravenous methylprednisolone 1 mg/kg 1-2 h prior to administration of AAV9-GLB1 gene transfer and began daily oral prednisolone 1 mg/kg starting on the first day after treatment and ending at day 4.

There were deviations in the immunosuppression treatment regimen since GT17 did not receive one of the four doses of rituximab (day -14, Table C1) following an infusion associated reaction to the first dose. In addition, following a positive Elispot, GT17 received additional daily oral prednisolone from day 14 to day 90 compared to the rest of the cohort. Lastly, five of the participants received intravenous immunoglobulin (IVIG) injections at their 6-month evaluation as clinically indicated.

**Figure C1.**
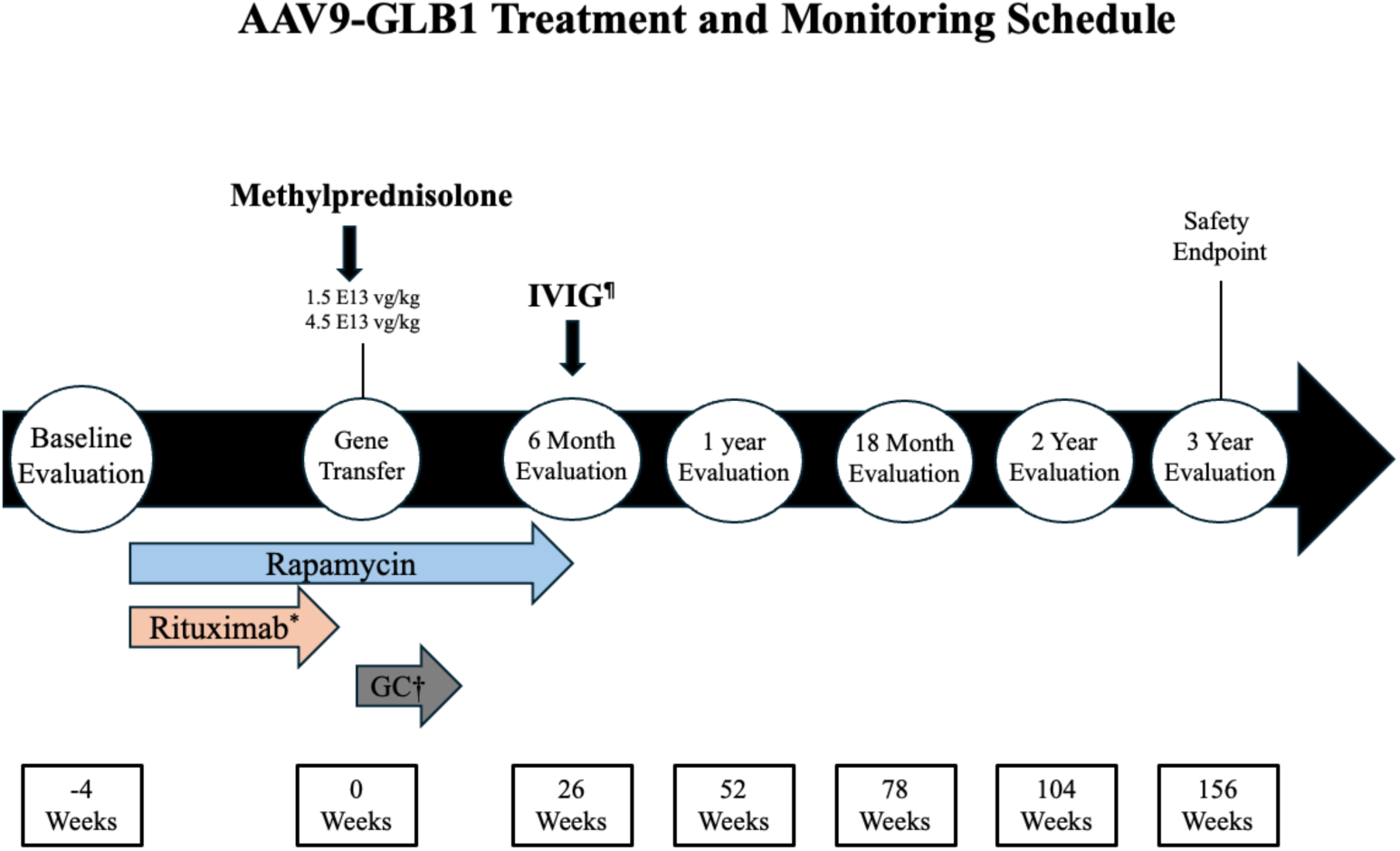
Immunosuppression treatment regimen and monitoring schedule. *4 Rituximab injections were given on days -21, -14, -7, and -1. GT17 did not receive a Rituximab dose on day -14 following his infusion associated reaction from the day -21 injection. ^¶^ Intravenous Immunoglobulin (IVIG) Injections were given for GT06, GT07, GT08, GT10, and GT11 as clinically indicated. † Oral glucocorticoids (GC) in the form of oral prednisolone were given daily starting on day one and ending after day four. GT17 received additional daily oral prednisolone from day 14 to day 90 due to a positive Elispot.

**Table C1.** Immunosuppression Regimen.

| Study ID | Rituximab infusion dates (Day) | Sirolimus start date (Day) | Methylprednisolone IV | Prednisone PO | Gamunex C – Immune Globulin Inj 10% (Visit timepoint) |
| --- | --- | --- | --- | --- | --- |
| GT03 | -21, -14, -7, -1 | -21 | Day 0 – One time dose | Day +1 – given daily for three days | n/a |
| GT04 | -21, -14, -7, -1 | -21 | Day 0 – One time dose | Day +1 – given daily for three days | n/a |
| GT06 | -21, -14, -7, -1 | -21 | Day 0 – One time dose | Day +1 – given daily for three days | Month 6 visit |
| GT07 | -21, -14, -7, -1 | -21 | Day 0 – One time dose | Day +1 – given daily for three days | Month 6 visit |
| GT08 | -21, -14, -7, -1 | -21 | Day 0 – One time dose | Day +1 – given daily for three days | Month 6 visit |
| GT10 | -21, -14, -7, -1 | -21 | Day 0 – One time dose | Day +1 – given daily for three days | Month 6 visit |
| GT11 | -21, -14, -7, -1 | -21 | Day 0 – One time dose | Day +1 – given daily for three days | Month 6 visit |
| GT12 | -21, -14, -7, -1 | -21 | Day 0 – One time dose | Day +1 – given daily for three days | n/a |
| GT17 | -21, -7, -1 | -21 | Day 0 – One time dose | Day +1 – given daily for three days<br>Then again from Day 14 to Day 90 | n/a |

#### Supplement D Biochemical Monitoring

Clinical laboratory assessments were primarily performed at the National Institutes of Health (NIH) Clinical Center, and specialized laboratory tests were performed at academic and contract research organizations (see below). As described in the protocol, all participants had extensive biochemical monitoring including blood draws, lumbar punctures, and saliva, feces, and urine collections. Blood draws were completed at days -30, -21, -14, -7, -1, 0 (before gene transfer), 1, 2-3, 4-6, 7, 14, 21, 30, 45, 60, 75, 90, 6 months, 1 year, 18 months, 2 years, and 3 years (after gene transfer). Lumbar punctures were performed on participants at baseline, 90 days, 6 months, 1 year, 18 months, 2 years, and 3 years post gene transfer. Data analysis and graph generation were performed in GraphPad Prism software for macOS, version 10.1.0 (GraphPad Software).

##### Local Clinical Laboratory Measures - NIH

Serum samples were obtained at the NIH Clinical Center at days -30, -21, -14, -7, -1, 0 (before gene transfer), 1, 2-3, 4-6, 7, 14, 21, 30, 90, 6 months, 1 year, 18 months, 2 years, and 3 years. Serum samples were analyzed at local laboratories for days 45, 60, and 75. Serum was analyzed for safety including aspartate aminotransferase (AST), alanine aminotransferase levels (ALT), and Gamma glutamyl transferase (GGT), and platelet counts. Per the protocol schedule of events, safety labs were analyzed on day -30 to -15, day -14, day -7, day -1, day 2-3, day 7-8, day 14, day 21, day 30, day 45, day 60, day 75, day 90, day 180, year 1, 18 motnhs, 2 years, and 3 years relative to gene transfer. D-dimer levels were analyzed on day -30 to -15, day 1, and day 2-3 relative to gene transfer. Complement C3 and C4 were analyzed on day -30 to -15 and day 7-8. Data collection and missing data are described for each participant in Tables D1-D9.

##### Baseline Anti-AAV9 Antibodies - ATHENA

Detection of baseline Anti-AAV9 antibodies in serum was performed by ATHENA DIAGNOSTICS INC (Worcester, MA) utilizing a CLIA certified (CLIA# 22D0069726) Enzyme Linked Immunosorbent Assay (ELISA). GT04’s baseline AAV9 antibody titer was performed by the University of Florida (BJB)

##### β-galactosidase activity – Auburn University

The activities of β-galactosidase in serum and cerebrospinal fluid (CSF) were measured by Auburn University (ALG) by incubating samples with 100 µL of 0.5 mM 4-methylumbelliferyl (4MU) β-D-galactoside, pH 3.8 (Sigma) in citrate-phosphate buffer (0.05 M citric acid monohydrate, 0.05 M disodium phosphate heptahydrate, 0.1 M sodium chloride) at 37°C for 1 h. Enzyme activity in the serum was determined using 10 µL of sample and activity in CSF was determined using 30 µL of sample. Enzyme activity was quenched with 3 mL of cold glycine carbonate buffer, pH 10.0 (0.17 M glycine, 0.17 M sodium carbonate, anhydrous) and fluorescence was measured with an excitation of 360 nm and emission of 450 nm on a BioTek Synergy H1 plate reader. Specific activity was expressed as nmol 4MU cleaved/mg protein/h after normalization to protein concentration as determined by the Lowry method. The controls utilized to calculate normal β-galactosidase activity consisted of 10 non-neurological participants (7 females) aged 6.35 ± 2.97 years who were enrolled in an NIH clinical protocol that allowed for use of research specimens. CSF was analyzed for β-galactosidase activity at baseline, day 90, day 180, year 1, 18 months, 2 years, and 3 years relative to gene transfer. Serum was analyzed for β-galactosidase activity at baseline, day 90, day 180, year 1, 2 years, and 3 years relative to gene transfer. GT04 did not have β-galactosidase activity analyzed for year 2 in CSF. GT17 did not have β-galactosidase activity analyzed for year 2 in serum and both serum and CSF in year 3 post gene transfer.

##### ELISPOT/Neutralizing Antibodies - University of Massachusetts Chan Medical School

Blood samples were sent to the University of Massachusetts Chan Medical School (ALK) for ELISPOT and analysis of Neutralizing Antibodies (NAb). Serum was screened for neutralizing antibodies as previously described.^4^ Luminescence was measured using a BioTek Synergy HTX reader. NAb titers are expressed as the highest dilution that inhibits β-galactosidase expression by at least 50% when compared to a negative mouse serum control. INF-y ELISpot assays were run as previously published.^4^ Spot forming unit (SFU) number was determined using Mabtech IRIS reader. Responses were considered positive when the number of SFU per 1e6 cells were >50 and at least 3-fold higher than the unstimulated control condition.

##### GM1 Ganglioside Levels – Pharmaron

CSF samples were sent to Pharmaron Lab Services LLC (Exton, PA) for analysis of GM1 ganglioside levels. A validated liquid chromatography-tandem mass spectrometry (LC-MS/MS) method was used to determine GM1 in human CSF samples. The standard calibration curve was composed of eight standards ranging from 5 to 500 ng/mL for GM1, and the quality control (QC) samples were prepared at the Low QC (15.0 ng/mL), Mid QC (100 ng/mL), and High QC (400 ng/mL) concentrations for GM1. For each sample analysis batch, 2 replicates at each QC concentration were analyzed with two sets of standard curves. One accuracy and precision batch with four levels of QCs, lower limit of quantification QC (5.00 ng/mL), Low QC (15.0 ng/mL), Mid QC (100 ng/mL), and High QC (400 ng/mL) concentrations for GM1 was performed before initiating sample analysis to qualify the analyst and instrument used. 7 paticipants (GT04, GT06, GT07, GT08, GT03, GT10, GT11, and GT12) had CSF analyzed for GM1 ganglioside levels at baseline, day 90, day 180, year 1, 18 months, year 2, and year 3 relative to gene transfer. GT04 did not have GM1 ganglioside levels analyzed in CSF for year 3 post gene transfer. GT17, did not have CSF analyzed for GM1 ganglioside levels analyzed at 2 and 3 years post gene transfer.

##### Viral Shedding - Pharmaron

###### Sample Collection

Viral shedding sample collection in all media began with GT06. Stored urine and serum were available for GT03 and GT04 for a retrospective analysis of viral shedding. Saliva, stool, and urine samples for viral shedding analysis were collected at baseline and after intravenous gene transfer at various timepoints, such as day 0, 7, 14, 21, 30, 45, 90, 180, 365, and 18 months for most participants. Saliva and urine were collected in a sterile specimen container, and stool was collected either in a sterile specimen container or a Norgen stool nucleic acid preservation tube. All specimens were kept at optimally cold temperatures, on wet ice or kept refrigerated throughout aliquoting into storage tubes, which were all stored in a −80°C freezer. All samples were transported on dry ice to Pharmaron in insulated dry ice shipping boxes and all were received in good, frozen condition for viral shedding analysis.

###### Sample Analysis

The qPCR reaction (rxn) contained PrimeTime® Gene Expression Master Mix (IDT), 400 nM forward and reverse primers, 200 nM probe, and nuclease free water (IDT). Primers and probe sequences and plasmid are proprietary. 10 µL of purified matrix was assayed in a 50 µL qPCR reaction. The qPCR reactions were assayed on an ABI QuantStudio 7 Pro Real-Time PCR System (ABI). The cycling parameters were as follows: 95°C for 10 min followed by 40 cycles of 95°C for 15 s and 64°C for 30 s. After cycling was complete, the plate was kept at 4°C. A standard curve made from linearized pDNA containing the target sequence (range: 25 to 1 × 10^7^ copies/rxn) was used to quantify the amount of target sequence in the sample (copies/rxn). The number of copies/rxn quantified in the sample was multiplied by 2 to calculate viral genomes (vg)/rxn. This corrects for the quantification of the single-stranded DNA (ssDNA) viral genomes relative to the double stranded DNA (dsDNA) standard curve. The qPCR assay was qualified with an LLOQ of 50 copies/rxn (100 vg/rxn) and a ULOQ of 1 × 10^6^ copies/rxn (2 × 10^6^ vg/rxn).

Urine, feces, and saliva were stored at −80°C. Nucleic acids were extracted from urine, saliva, and feces using the QIAamp® 96 Virus QIAcube® HT Kit (Qiagen) and the QIAcube HT (Qiagen) automated, high-throughput nucleic acid purification system following manufacturer’s instructions. The isolated nucleic acids were eluted with 130 µL of AVE buffer (Qiagen) and the final eluate was approximately 100 µL. This eluate is referred to as purified matrix. The purified matrix was stored at −80°C.

Prior to extraction, urine and feces were pretreated following the manufacture’s protocol (QIAamp 96 Virus QIAcube HT Handbook, Qiagen). Saliva had no pretreatment. Urine was pretreated by combining 200 µL of urine with 15 µL of Proteinase K and 60 µL of ATL buffer (Qiagen). 200 µL of this 275 µL pretreated urine was extracted. Feces was pretreated by combining 100 µL (100 mg) of feces with 900 µL of 0.90% NaCl. The suspension was vortexed vigorously followed by centrifugation at 20K ×g for 1 min. 200 µL of the supernatant was extracted.

The conversion of copies/rxn to vg/mL of original matrix used the following equations, which account for the dilution of the sample by pretreatment and the volume (100 µL) of eluate.

Saliva: (Copies/rxn × 2 (vg correction) / 20.0 µL) x 1,000 µL/mL = vg/mL matrix. 20.0 µL of original matrix used: (200 µL saliva extracted / 100 µL of eluate) × 10 µL used in PCR.

Urine: (Detected copies × 2 (vg correction) / 14.5 µL) x 1,000 µL/mL = vg/mL matrix. 14.5 µL of original matrix used: (200 µL urine / 275 µL pretreated urine) × 200 µL pretreated urine extracted / 100 µL of eluate × 10 µL used in PCR.

Feces: (Detected copies × 2 (vg correction) / 2.00 µL) x 1,000 µL/mL = vg/mL matrix. (100 µL(mg) feces / 1000 µL pretreated feces) × 200 µL pretreated feces extracted / 100 µL of eluate × 10 µL used in PCR.

###### DNA Extraction, Quantification, and Quantitative PCR

DNA was extracted from serum samples collected at various time points post-dosing using a DNeasy Blood and Tissue kit (Qiagen, Valencia, CA) according to manufacturer’s and in-house protocols. DNA was quantified using the NanoDrop One Microvolume UV-Vis Spectrophotometer (Thermo Fisher) according to manufacturer’s and in-house protocols. Quantitative PCR (qPCR) was performed on a QuantStudio 3 (Applied Biosystems) using the QuantStudio Design and Analysis Software v1.4.1 (Applied Biosystems) according to manufacturer’s instructions and in-house protocols. A primer and probe set were designed to the chicken beta actin (CB) promoter that is found in the target plasmid, pAAV-CB-hβgal. Briefly, an 8-log standard curve covering a range of 1E+8 through 1E+0 copies was generated from a stock plasmid and Ct values for each standard were plotted against the log concentration of copies per sample. The linear regression of the resultant plot was utilized to calculate the copies within each sample. Each 20 µL reaction contained 13 µL of TaqPath™ ProAmp™ Master Mix (ThermoFisher Scientific), up to 0.01 µg of sample DNA, 700 nM of the forward and reverse primers, and 100 nM of FAM labeled probe. Each sample was tested in triplicate, with one replicate containing 10 copies of the target plasmid spiked in to test for inhibition of the reaction. Cycling followed manufacturer recommended guidelines, 5 min at 95 °C for initial denaturation and enzyme activation, followed by 40 cycles altering between a 15 s period at 95 °C for denaturing and a 60 s period at 60 °C for annealing and extension. Results were averaged across two replicates and normalized to vector genome copies microgram of DNA.

CB Forward:

5’-CATCTACGTATTAGTCATCGCTATTACCA-3’

CB Reverse:

5’-CCCATCGCTGCACAAAATAATTA -3’

CB Probe:

6FAM-CCACGTTCTGCTTCACTCTCCCCATC-TAMRA

##### H3N2B Levels – Washington University

CSF, serum, and urine collections were sent to Washington University St. Louis (XJ) to analyze H3N2b levels. As described in Pell et al.^5^ liquid chromatography-tandem mass spectrometry (LC-MS/MS) was performed on all samples to determine the concentration of H3N2b utilizing validated methodology. Urine H3N2b levels were normalized to creatine levels as described in Leonard et al.^6^ CSF was analyzed for H3N2b levels at baseline, day 90, day 180, year 1, 18 months, 2 years, and 3 years relative to gene transfer. Serum and urine were analyzed for H3N2b levels at baseline, day 30, day 90, day 180, year 1, 18 months, 2 years, and 3 years relative to gene transfer. 7 paticipants (GT04, GT06, GT07, GT08, GT03, GT10, GT11, and GT12) had complete H3N2b studies. GT04 did not have serum, urine, or CSF H3N2b levels analyzed in serum, CSF, or urine for year 3 following gene transfer. GT17 did not have year 2 and year 3 CSF H3N2b levels analyzed, year 3 urine H3N2b levels, or year 3 serum H3N2b levels analyzed.

##### IgG and IgM – University of Florida

Blood samples were sent to the University of Florida Powell Gene Therapy Center (MC, BJB), for analysis of serum IgG and IgM. The first 7 participants (GT04, GT06, GT07, GT08, GT03, GT10, and GT11) had serum analyzed for IgG and IGM analyzed at baseline (day -30), day 7, day 14, day 21, day 30, day 45, day 60, day 75, day 90, day 180, and day 365. One participant (GT12), had serum analyzed for IgG and IGM analyzed at baseline (day -30), day 7, day 14, day 21, day 30, day 45, day 60, day 75, day 90, and day 180. GT17 did not have serum analyzed for IgG or IgM.

###### Antibody Assay

Total anti-AAV9 IgG and IgM levels in serum were evaluated by enzyme-linked immunosorbent assays (ELISA). Serum samples from the subjects were assayed for total (neutralizing and non-neutralizing) antibodies to AAV9 capsid as previously described.^7^ Briefly, 96-well plates were coated with 1 × 10^9^ AAV9 particles per well in sodium bicarbonate buffer, pH 8.4, overnight at 4 °C. Subsequently, the plates were washed with a solution containing phosphate-buffered saline (PBS) and 0.05% Tween-20 (PBS-T) and then blocked with 10% fetal bovine serum (FBS; Cellgro) for 2 h at 37 °C. After being washed with PBS T, the samples were serially diluted from 1:10 to 1:10,240 with a known positive human standard and allowed to bind overnight at 4 °C. The plates were washed again, followed by addition of a secondary antibody (goat anti-human IgG or IgM conjugated with horseradish peroxidase [HRP]; Invitrogen) at a dilution of 1:20,000 for 2 h at 37 °C. Finally, the plates were washed and incubated with 3,3′,5,5′-tetramethylbenzidine (TMB) peroxidase substrate (Seracare Life Sciences) in the dark. Reactions were stopped with 0.1M phosphoric acid. The reaction product was measured by spectrophotometric absorbance at 450 nm using Gen5 Microplate Reader and Imager Software (BioTek Instruments). Sample titers were calculated using the mean absorbance of up to three sequential dilutions that were within the linear region of a 4-parameter logistic standard curve generated by a known positive human standard.

**Table D1.** GT04 Biochemical Analysis Testing.

|  | AST | ALT | GGT | Platelet | D-Dimer | C3 | C4 |
| --- | --- | --- | --- | --- | --- | --- | --- |
| Day -30 | X | X | X | X |  |  |  |
| Day -21 | X | X | X | X |  |  |  |
| Day -14 | X | X | X | X |  |  |  |
| Day -7 | X | X | X | X |  |  |  |
| Day -1 | X | X | X | X |  |  |  |
| Day 0 | X | X | X |  | X | X | X |
| Day 1 |  |  |  | X | X |  |  |
| Day 2-3 | X | X | X | X | X | X | X |
| Day 4-6 | X | X | X | X | X | X | X |
| Day 7 | X | X | X | X | X |  |  |
| Day 14 | X | X | X | X | X |  |  |
| Day 21 | X | X | X | X |  |  |  |
| Day 30 | X | X | X | X | X |  |  |
| Day 45 | X | X | X | X |  |  |  |
| Day 60 | X | X | X | X |  |  |  |
| Day 75 | X | X | X | X |  |  |  |
| Day 90 | X | X | X | X |  |  |  |
| Day 180 | X | X | X | X |  | X | X |
| Year 1 |  | X |  | X | X |  |  |
| Week 78 | X | X | X | X | X | X | X |
| Year 2 |  |  |  |  |  |  |  |
| Year 3 |  |  |  |  |  |  |  |

X = test completed, Red box indicates test was not completed according to protocol.

**Table D2.** GT06 Biochemical Analysis Testing.

|  | AST | ALT | GGT | Platelet | D-Dimer | C3 | C4 |
| --- | --- | --- | --- | --- | --- | --- | --- |
| Day -30 | X | X | X | X | X | X | X |
| Day -21 | X | X | X | X |  | X | X |
| Day -14 | X | X | X | X |  |  |  |
| Day -7 | X | X | X | X |  |  |  |
| Day -1 | X | X | X | X |  |  |  |
| Day 0 |  |  |  |  | X |  |  |
| Day 1 | X | X | X | X | X | X | X |
| Day 2-3 | X | X | X | X | X | X | X |
| Day 4-6 | X | X | X |  |  | X | X |
| Day 7 | X | X | X | X | X | X | X |
| Day 14 | X | X | X | X | X | X | X |
| Day 21 | X | X | X | X | X | X | X |
| Day 30 | X | X | X | X | X | X | X |
| Day 45 | X | X | X | X | X | X | X |
| Day 60 | X | X | X | X |  | X | X |
| Day 75 | X | X | X | X | X | X | X |
| Day 90 | X | X | X | X |  |  |  |
| Day 180 | X | X | X | X | X | X | X |
| Year 1 | X | X | X | X | X | X | X |
| Week 78 | X | X | X | X |  | X | X |
| Year 2 | X | X | X | X | X |  |  |
| Year 3 | X | X | X | X | X | X | X |

**Table D3.** GT07 Biochemical Analysis Testing.

|  | AST | ALT | GGT | Platelet | D-Dimer | C3 | C4 |
| --- | --- | --- | --- | --- | --- | --- | --- |
| Day -30 | X | X | X | X | X | X | X |
| Day -21 | X | X | X | X |  | X | X |
| Day -14 | X | X | X | X |  |  |  |
| Day -7 | X | X | X | X |  |  |  |
| Day -1 | X | X | X | X |  |  |  |
| Day 0 |  |  |  |  | X |  |  |
| Day 1 | X | X | X | X | X | X | X |
| Day 2-3 | X | X | X | X | X | X | X |
| Day 4-6 |  |  |  |  |  | X | X |
| Day 7 | X | X | X | X | X | X | X |
| Day 14 | X | X | X | X | X | X | X |
| Day 21 | X | X | X | X | X | X | X |
| Day 30 | X | X | X | X | X | X | X |
| Day 45 | X | X | X | X | X | X | X |
| Day 60 | X | X | X | X |  | X | X |
| Day 75 | X | X | X | X | X | X | X |
| Day 90 | X | X | X | X |  |  |  |
| Day 180 | X | X | X | X | X | X | X |
| Year 1 | X | X | X | X | X | X | X |
| Week 78 | X | X | X | X |  | X | X |
| Year 2 | X | X | X | X | X |  |  |
| Year 3 | X | X | X | X | X | X | X |

**Table D4.** GT08 Biochemical Analysis Testing.

|  | AST | ALT | GGT | Platelet | D-Dimer | C3 | C4 |
| --- | --- | --- | --- | --- | --- | --- | --- |
| Day -30 | X | X | X | X | X | X | X |
| Day -21 | X | X | X | X |  |  |  |
| Day -14 | X | X | X | X |  |  |  |
| Day -7 | X | X | X | X |  |  |  |
| Day -1 | X | X | X | X | X |  |  |
| Day 0 |  |  |  | X | X |  |  |
| Day 1 | X | X | X | X | X | X | X |
| Day 2-3 | X | X | X | X | X | X | X |
| Day 4-6 | X | X | X |  |  | X | X |
| Day 7 | X | X | X | X | X | X | X |
| Day 14 | X | X | X | X | X | X | X |
| Day 21 | X | X | X | X | X | X | X |
| Day 30 | X | X | X | X | X | X | X |
| Day 45 | X | X | X | X | X | X | X |
| Day 60 | X | X | X | X |  | X | X |
| Day 75 | X | X | X | X | X | X | X |
| Day 90 | X | X | X | X |  |  |  |
| Day 180 | X | X | X | X | X | X | X |
| Year 1 | X | X | X | X | X | X | X |
| Week 78 | X | X | X | X |  | X | X |
| Year 2 | X | X | X | X | X |  |  |
| Year 3 | X | X | X | X | X | X | X |

**Table D5.**
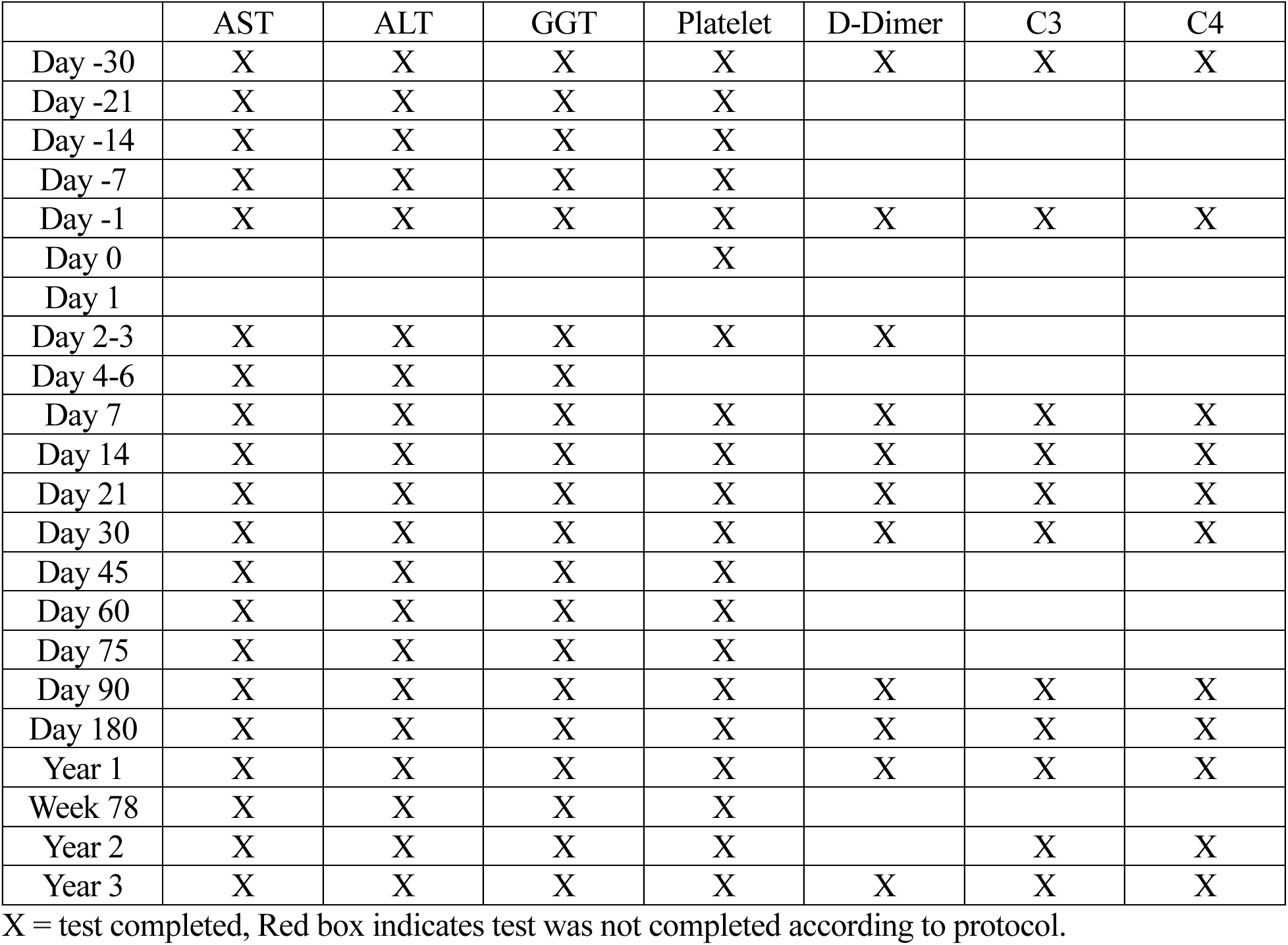
GT03 Biochemical Analysis Testing.

**Table D6.** GT10 Biochemical Analysis Testing.

|  | AST | ALT | GGT | Platelet | D-Dimer | C3 | C4 |
| --- | --- | --- | --- | --- | --- | --- | --- |
| Day -30 | X | X | X | X |  | X | X |
| Day -21 | X | X | X | X |  |  |  |
| Day -14 | X | X | X | X |  |  |  |
| Day -7 | X | X | X | X |  |  |  |
| Day -1 | X | X | X | X | X | X | X |
| Day 0 |  |  |  |  |  |  |  |
| Day 1 | X | X | X | X | X | X | X |
| Day 2-3 | X | X | X | X | X | X | X |
| Day 4-6 | X | X | X | X | X | X | X |
| Day 7 | X | X | X | X | X | X | X |
| Day 14 | X | X | X | X | X | X | X |
| Day 21 | X | X | X | X | X | X | X |
| Day 30 | X | X | X | X | X | X | X |
| Day 45 | X | X | X | X | X | X | X |
| Day 60 | X | X | X | X | X | X | X |
| Day 75 | X | X | X | X | X | X | X |
| Day 90 | X | X | X | X | X | X | X |
| Day 180 | X | X | X | X | X | X | X |
| Year 1 | X | X | X | X | X | X | X |
| Week 78 | X | X | X | X | X |  |  |
| Year 2 | X | X | X | X | X | X | X |
| Year 3 | X | X | X | X | X | X | X |

**Table D7.**
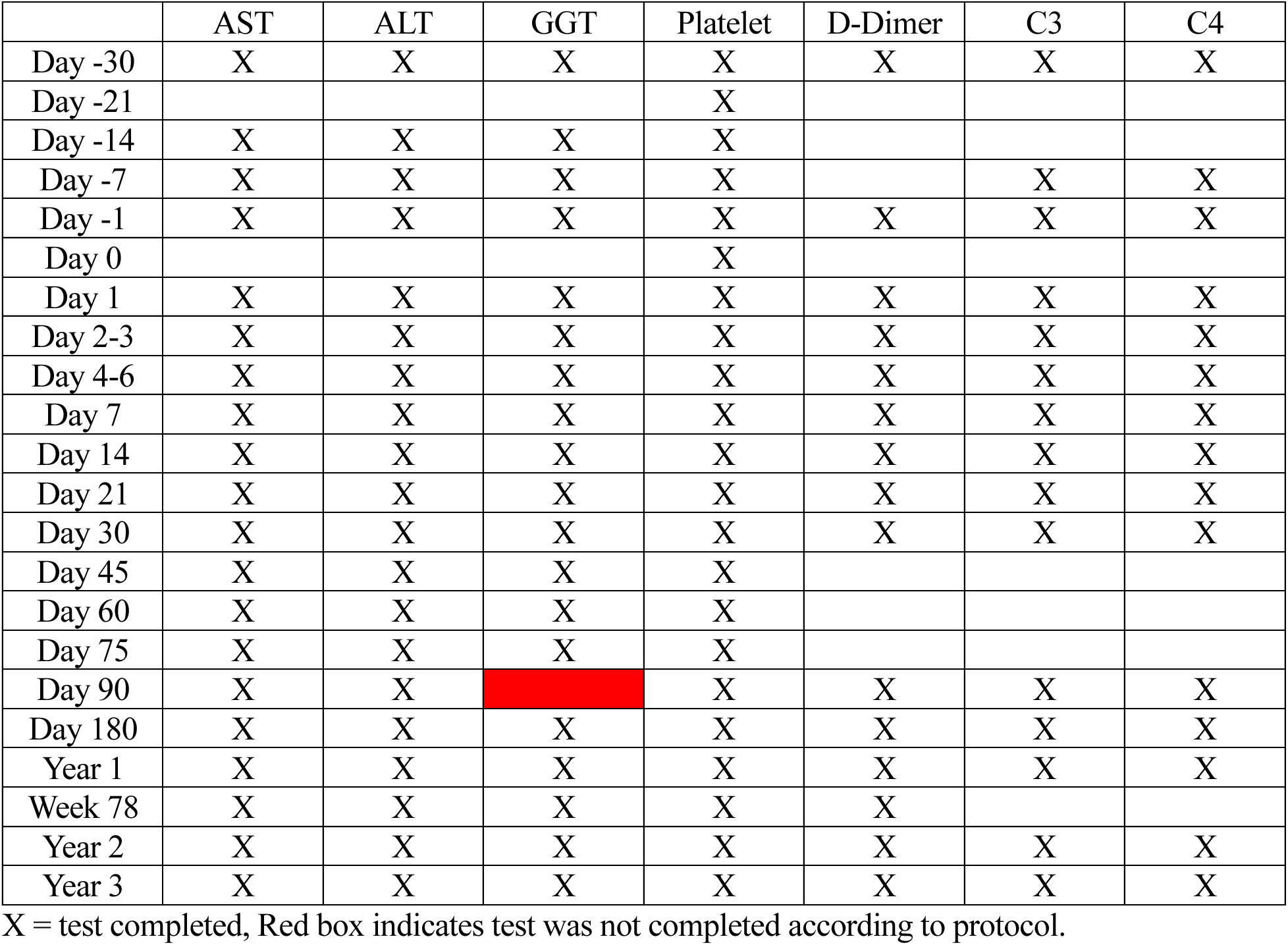
GT11 Biochemical Analysis Testing.

**Table D8.** GT12 Biochemical Analysis Testing.

|  | AST | ALT | GGT | Platelet | D-Dimer | C3 | C4 |
| --- | --- | --- | --- | --- | --- | --- | --- |
| Day -30 | X | X | X | X |  |  |  |
| Day -21 |  |  |  | X |  |  |  |
| Day -14 | X | X | X | X |  |  |  |
| Day -7 | X | X | X | X |  |  |  |
| Day -1 |  |  |  |  |  |  |  |
| Day 0 | X | X | X | X | X | X | X |
| Day 1 |  |  |  |  |  |  |  |
| Day 2-3 | X | X | X | X | X | X | X |
| Day 4-6 | X | X | X | X |  | X | X |
| Day 7 | X | X | X | X | X | X | X |
| Day 14 | X | X | X | X |  | X | X |
| Day 21 | X | X | X | X |  | X | X |
| Day 30 | X | X | X | X | X | X | X |
| Day 45 | X | X | X | X |  |  |  |
| Day 60 | X | X | X | X |  |  |  |
| Day 75 | X | X | X | X |  |  |  |
| Day 90 | X | X | X | X |  | X | X |
| Day 180 | X | X | X | X | X | X | X |
| Year 1 | X | X | X | X | X |  |  |
| Week 78 | X | X | X | X | X | X | X |
| Year 2 | X | X | X | X | X | X | X |
| Year 3 | X | X | X | X | X |  |  |

X = test completed, Red box indicates test was not completed according to protocol.

**Table D9.** GT17 Biochemical Analysis Testing.

|  | AST | ALT | GGT | Platelet | D-Dimer | C3 | C4 |
| --- | --- | --- | --- | --- | --- | --- | --- |
| Day -30 | X | X | X | X | X | X | X |
| Day -21 | X | X | X | X | X | X | X |
| Day -14 |  |  |  |  |  |  |  |
| Day -7 | X | X | X | X |  |  |  |
| Day -1 | X | X | X | X | X | X | X |
| Day 0 | X | X | X | X | X | X | X |
| Day 1 | X | X | X | X | X | X | X |
| Day 2-3 | X | X | X | X | X | X | X |
| Day 4-6 | X | X | X | X | X | X | X |
| Day 7 | X | X | X | X | X | X | X |
| Day 14 | X | X | X | X | X | X | X |
| Day 21 | X | X | X | X | X | X | X |
| Day 30 | X | X | X | X | X | X | X |
| Day 45 |  |  |  |  |  |  |  |
| Day 60 |  |  |  |  |  |  |  |
| Day 75 |  |  |  |  |  | X | X |
| Day 90 | X | X | X | X | X | X | X |
| Day 180 | X | X | X | X | X | X | X |
| Year 1 | X | X | X | X | X | X | X |
| Week 78 | X | X | X | X | X |  |  |
| Year 2 | X | X | X | X | X | X | X |
| Year 3 |  |  |  |  |  |  |  |

X = test completed, Red box indicates test was not completed according to protocol.

#### Supplement E Clinical Outcome Assessments

##### Vineland Adaptive Behavior Composite (ABC)

Administration of the Vineland Adaptive Behavior Scale was completed as described in D’Souza et al.^1^ as a semi-structured interview with the participant’s parent/caregiver. All AAV9-GLB1 participants received only the third edition of the Vineland (Vineland-3).^8^ The Vineland outputs both a composite score and domain level scores (including Communication, Socialization, Daily Living, and Motor). The Vineland subdomains of interest were Receptive Communication, Expressive Communication, Fine Motor, and Gross Motor^1^. Performance was operationalized using the growth scale value (GSV), a version of the raw score that has been transformed to an interval-level scale. GSVs have conditional standard errors of measurement, allowing for the statistical evaluation of individual change,^9^ and a monotonic relationship with change in the underlying construct.^10^ Too few natural history data in the age range of the trial participants were available to support external comparison.

Six administrations of the Vineland were given during the main portion of the study, i.e.,: (1) between 36 and 7 days prior to AAV9-GLB1 administration; (2) at approximately 6 months post AAV9-GLB1 administration; (3) at approximately 1 year post AAV9-GLB1 administration; (4) at approximately 18 months post AAV9-GLB1 administration; (5) at approximately 2 years post AAV9-GLB1 administration; and (6) at approximately 3 years post AAV9-GLB1 administration. Here we report Vineland-3 GSV for fine motor skills, gross motor skills, receptive communication skills, and expressive communication skills.

The statistical analysis plan dictated a mixed model for repeated measures for the evaluation of change from baseline on the Vineland GSV at Year 2; the Year 3 timepoint was also included as an exploratory analysis. Fixed effects were study visit (Baseline, Month 6, Month 12, Month 18, Year 2, Year 3) and the *a priori* covariate chronological age at baseline. A random subject-level intercept and an unstructured variance-covariance matrix were used. This was an intent-to-treat analysis, so the participant with no Year 3 data was included in the analysis. The a priori hypothesis was that a significant increase from baseline in GSV score would be observed at Year 2, and alpha was set to 0.05. The mixed models were evaluated using lme4^11^ and uncorrected *p*-values with Satterthwaite’s approximation for degrees of freedom) were calculated using lmerTest.^12^

##### Clinical Global Impression (CGI)

Prospective evaluations of the Clinical Global Impression (CGI) scales were performed for each patient. The CGI scales are a brief assessment of a patient’s global clinical presentation as determined from the clinician’s view.^13,14^ The CGI Severity (CGI-S) scale, completed at baseline only, is scored from 1 (normal) to 7 (among the most extremely ill) and assesses the patient’s global severity (Table E1). The CGI Improvement (CGI-I) scale, completed at all post-infusion assessments, evaluates the participants’ change in clinical presentation from the baseline evaluation and is scored from 1 (very much improved) to 7 (very much worse) with 4 corresponding to “no change” in the patient’s clinical presentation (Table E2). As described in Lewis et al.,^15^ CGI scores were based on a consensus reached among three researchers (PD, MTA, and CJT in this study) and were obtained within two weeks of the patient’s weeklong evaluation at the NIH.

CGI-S were evaluated only at the baseline evaluation for each patient. 7 evaluations of CGI-I were conducted, i.e.,: (1) at 1 month post AAV9-GLB1 administration; (2) at 3 months post AAV9-GLB1 administration; (3) at 6 month post AAV9-GLB1 administration; (4) at 1 year post AAV9-GLB1 administration; (5) at 18 months post AAV9-GLB1 administration; (6) at 2 years post AAV9-GLB1 administration, and (7) at 3 years post AAV9-GLB1 administration.

**Table E1.** Clinical Global Impression Severity (CGI-S)

| **CGI-S Score** | **Baseline Severity** |
| --- | --- |
| 1 | Normal |
| 2 | Borderline ill |
| 3 | Midly ill |
| 4 | Moderately ill |
| 5 | Markedly ill |
| 6 | Severely ill |
| 7 | Among the most extremely ill |

**Table E2.** Clinical Global Impression Change (CGI-I)

| **CGI-I Score** | **Change Severity** |
| --- | --- |
| 1 | Very much improved |
| 2 | Much improved |
| 3 | Minimally improved |
| 4 | Normal |
| 5 | Minimally worse |
| 6 | Much worse |
| 7 | Very much worse |

#### Supplement F MRI/DTI/MRS Acquisition and Analysis

Gene therapy treated GM1 gangliosidosis participants underwent MRI studies at baseline (day - 30), 1 year, 2 years, and 3 years relative to gene transfer. MRI studies included T1-weighted scans, diffusion tensor imaging (DTI), and magnetic resonance spectroscopy (MRS) studies in the same session. Changes in brain magnetic resonance imaging were assessed in relation to historical GM1 controls (see below). Volumetric analysis was performed on T1-weighted scans, differential fiber tractography was performed on DTI scans to assess changes in myelination, and MRS was analyzed for changes in *N*-acetylaspartate+*N*-acetylaspartyl glutamate (NAA), a marker of neuronal health. MRI imaging acquisition and processing are described below. Longitudinal neuroimaging data were descriptive in nature. When appropriate, data are reported as means (± SD). Data analysis and graph generation were performed in GraphPad Prism software for macOS, version 10.1.0 (GraphPad Software).

##### NHGRI Natural History Study (NCT00029965)^16^

The NIH clinical protocol, “Natural History of Glycosphingolipid Storage Disorders and Glycoprotein Disorders”, encompasses a wider patient population including those with GM1 gangliosidosis, GM2 gangliosidosis, sialidosis, and galactosialidosis. Data included in this study were a subset of those from the aforementioned study, including only those with a GM1 gangliosidosis diagnosis, and a phenotype consistent with Type II disease. Diagnoses were made by documented enzyme deficiency and/or mutation analysis in a CLIA-approved laboratory.

Type II GM1 gangliosidosis participants were further sub-typed into late-infantile (earlier onset of symptoms; never achieved running) and juvenile (able to run by age 2).^1–3^ All Type II GM1 gangliosidosis participants who completed a sedated MRI under the Natural History protocol were included in MRI portion of this study. Participants who were initially enrolled in the Natural History protocol (NCT00029965)^16^ and later enrolled in the gene therapy treated protocol (NCT03952637)^17^ were excluded from the Natural history MRI and MRS datasets. Cross-sectional evaluations were performed with the most recent MRI study for late-infantile participants with repeated MRI collection to best overlap with age of the gene therapy treated cohort. For juvenile participants, the earliest MRI collection was utilized to best overlap with the gene therapy treated cohort. 11 MRI scans from 11 late-infantile natural history participants were included in this analysis (Table F1) and 18 MRI scans from 18 juvenile natural history participants were included in this analysis (Table F2). 9 MRS scans from 9 late-infantile natural history participants were included in the MRS analysis (Table F3). 17 MRS scans from 17 juvenile natural history participants were included in the MRS analysis (Table F4). MRS scans did not always reach a sufficient signal to noise ratio, leading some scans to be discarded. For participants with multiple MRS scans, an earlier scan was used for late-infantile participants, and a later scan was used for juvenile participants given they met the signal to noise ratio cutoff.

##### MRI Acquisition

All magnetic resonance imaging, including T1-weighted, Diffusion Tensor Imaging (DTI), and Magnetic Resonance Spectroscopy (MRS), was performed on a 3-Tesla Phillips (Philips Healthcare, Best, The Netherlands) Achieva System with an 8-channel SENSE head coil.^1,18,19^ All MRI acquisition for both the gene therapy treated GM1 participants and untreated natural history study GM1 participants was performed under anesthesia.

##### T1-Weighted Acquisition and Analysis

T1- weighted images for both the gene therapy treated and NHGRI natural history participants were acquired using a 3D T1-weighted protocol with a slice thickness of 1 mm, repetition time (TR) of 11 ms, and an echo time (TE) of 7 ms, a flip angle of 6 degrees, field of view of 220 mm, and two signal averages or number of excitations (NEX). Unprocessed digital imaging and communications in medicine (DICOM) images were converted to NIfTI using *dcm2niix*.^20^ T1-weighted MRI scans were then sent through volBrain’s *vol2Brain* to calculate bilateral volumes of the whole brain, lateral ventricles, and thalamus.^21^ As described in Lewis et al.,^22^ we have previously shown that *vol2Brain* offers accurate automated volumetric segmentation of T1-weighted MRI data in this cohort. A separate simple linear regression was performed for both late-infantile and juvenile cross-sectional natural history data.

##### Diffusion Tensor Imaging Acquisition and Analysis

Diffusion tensor imaging (DTI) was acquired during the same session for both cohorts with the following parameters: TR/TE = 6400/100 ms, 15-gradient directions (30-directions for the Natural History protocol), b-values = 0 and 1000 s/mm^2^, slice thickness = 2.5 mm, acquisition matrix = 128 × 128, NEX = 1, FOV = 24 cm. One natural history patient, GT17’s untreated older sibling’s (Figure 3) DTI acquisition was performed utilizing the 15-direction gene therapy protocol.

DTI were analyzed by differential tractography, a subject level analysis technique utilized to compare changes in DTI calculated metrics over time.^19,23^ We used differential tractography to compare changes in fractional anisotropy (FA) in gene therapy treated GM1 participants for three years post intravenous administration of AAV9-GLB1. Differential tractography was performed as described in Lewis et al.^19^ First, unprocessed DTI digital imaging and communications in medicine (DICOM) images were converted to NIfTI using *dcm2niix*.^20^ DTI were then preprocessed using MRtrix3’s (MRtrix, v3.0.4)^24^ *dwifslpreproc*^25,26^ command leveraging MRtrix3’s *dwi2mask*^27^ function followed by FSL’s (FSL, v6.0.5)^28,29^ *eddy*^25^ function. Preprocessed DTI images were then imported into DSI Studio (DSI Studio, v November 6, 2023) where the DTI images were quality checked for bad slices; a U-Net mask was created to remove non-brain regions and generalized q-sampling imaging (GQI)^30^ based reconstruction was performed with a diffusion sampling length ratio of 1.25.

As described in Lewis et al.,^19^ a baseline whole brain tractography was performed with an angular threshold of 60 degrees, a step size of one millimeter, one million seeds, a maximum length threshold of 200 millimeters, and a minimum length threshold of 30 millimeters. The fractional anisotropy (FA) map for each patient’s baseline scan was exported to a NIfTI file and utilized for differential fiber tractography. Whole brain differential tractography was calculated utilizing the same parameters as the baseline whole brain tractography using a 30 percent FA threshold. Follow-up scans were compared to the original baseline scan where fiber tract gains were determined in fiber tracts where the difference in FA between the follow-up scan and the baseline scan exceeded the FA threshold. Fiber tract losses were determined where the difference between the baseline scan and the follow-up scan exceeded the same threshold. Net fiber tract metrics for the number of fiber tracts and the volume of those tracts were determined as the difference between the fiber tract gains and fiber tract losses relative to the baseline whole brain tractography as a percentage change.

##### Magnetic Resonance Spectroscopy Acquisition and Analysis

As described in D’Souza et al.^1^ single voxel magnetic resonance spectroscopy (MRS) was performed on the left centrum semiovale (LCSO, Figure F1) during the same sedated MRI acquisition session. Voxels were graphically prescribed on the 3-D T1 weighted image in three planes. 1H-MRS was acquired with PRESS localization, CHESS water suppression, and the following parameters: TE = 38 ms, TR = 2,000 ms, and NEX = 128. A water spectrum was acquired with TE = 38 ms, TR = 5,000 ms, and NEX = 16 at the LCSO. A heavily T2 weighted image was also acquired at the LCSO with TE = 500 ms, TR = 3,000 ms, ETL = 8 to correct for CSF. A water containing phantom was placed in the field-of-view of the CSF correction image.

Post-processing was performed using LCModel,^31^ followed by correcting for estimated water tissue,^32^ and T1 of the metabolites in the tissue.^33^ No correction was made for T2 decay of the metabolites. Based on the results from the natural history study,^5^ *N*-acetylaspartate + *N*-acetylaspartyl glutamate (NAA) was analyzed for all the participants. Post-processing for CSF was performed in accordance with previously described reports.^34–36^

Reference curves were calculated using data from 38 individuals undergoing the same MRS acquisition protocol at the National Institutes of Health Clinical Center for other study protocols who were either asymptomatic or neurologically pre-symptomatic. Reference curves were generated by fitting the model. NAA concentrations for untreated and treated GM1 participants were subtracted from the age-specific reference concentration to quantify the age-appropriate difference from the reference curve. Acquisition and analysis of the MRS data was uniform between the reference data, untreated Natural History GM1 participants, and gene therapy treated GM1 gangliosidosis participants.

**Table F1:**
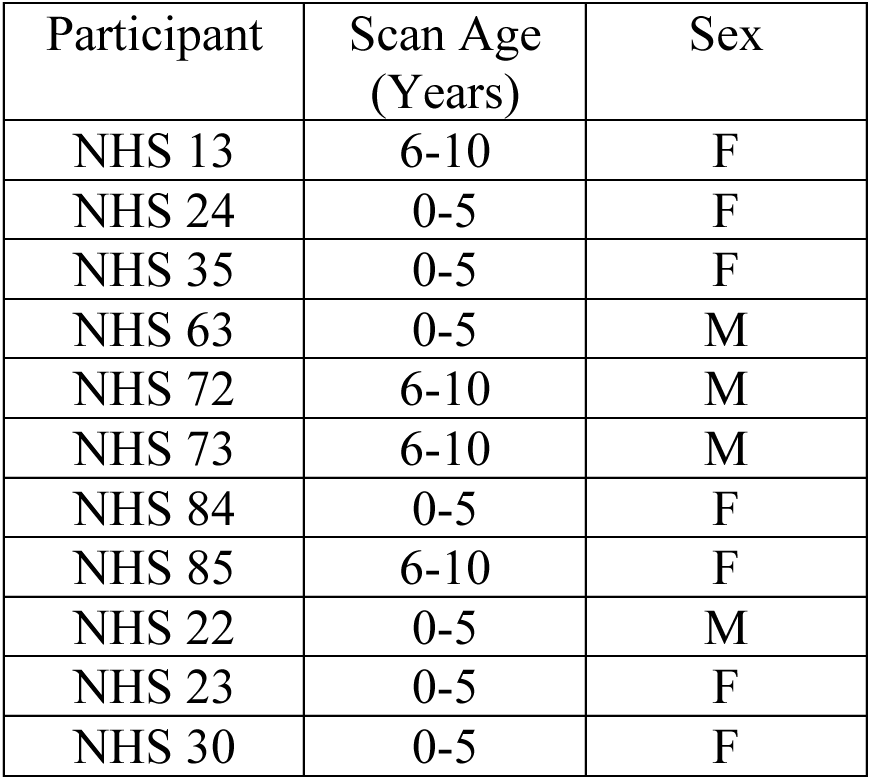
NHGRI GM1 Natural History Study Late-Infantile GM1 Gangliosidosis T1-weighted MRI Cohort (*n* = 11).

| Participant | Scan Age (Years) | Sex |
| --- | --- | --- |
| NHS 13 | 6-10 | F |
| NHS 24 | 0-5 | F |
| NHS 35 | 0-5 | F |
| NHS 63 | 0-5 | M |
| NHS 72 | 6-10 | M |
| NHS 73 | 6-10 | M |
| NHS 84 | 0-5 | F |
| NHS 85 | 6-10 | F |
| NHS 22 | 0-5 | M |
| NHS 23 | 0-5 | F |
| NHS 30 | 0-5 | F |

**Table F2:** NHGRI GM1 Natural History Study Juvenile GM1 Gangliosidosis T1-weighted MRI Cohort (*n* = 18).

| Participant | Scan Age (Years) | Sex |
| --- | --- | --- |
| NHS 03 | 16-20 | M |
| NHS 09 | 11-15 | M |
| NHS 10 | 11-15 | F |
| NHS 11 | 11-15 | M |
| NHS 20 | 11-15 | F |
| NHS 25 | 11-15 | F |
| NHS 26 | 11-15 | M |
| NHS 27 | 6-10 | M |
| NHS 28 | 16-20 | F |
| NHS 54 | 6-10 | F |
| NHS 58 | 11-15 | F |
| NHS 71 | 11-15 | F |
| NHS 93 | 6-10 | F |
| NHS 94 | 6-10 | M |
| NHS 118 | 10-15 | F |
| NHS 53 | 10-15 | F |
| NHS 69 | 0-5 | F |
| NHS 97 | 0-5 | F |

**Table F3:** NHGRI GM1 Natural History Study Late-Infantile GM1 Gangliosidosis Magnetic Resonance Spectroscopy Cohort (*n* = 9).

| Participant | Scan Age (Years) | Sex |
| --- | --- | --- |
| NHS 13 | 6-10 | F |
| NHS 22 | 0-5 | M |
| NHS 23 | 6-10 | F |
| NHS 30 | 0-5 | F |
| NHS 63 | 0-5 | M |
| NHS 72 | 6-10 | M |
| NHS 73 | 6-10 | M |
| NHS 84 | 0-5 | F |
| NHS 85 | 6-10 | F |

**Table F4:** NHGRI GM1 Natural History Study juvenile GM1 Gangliosidosis Magnetic Resonance Spectroscopy Cohort (*n* = 17).

| Participant | Scan Age (Years) | Sex |
| --- | --- | --- |
| NHS 03 | 16-20 | M |
| NHS 09 | 11-15 | M |
| NHS 10 | 11-15 | F |
| NHS 11 | 11-15 | M |
| NHS 20 | 11-15 | F |
| NHS 25 | 11-15 | F |
| NHS 26 | 11-15 | M |
| NHS 27 | 6-10 | M |
| NHS 28 | 16-20 | F |
| NHS 54 | 6-10 | F |
| NHS 58 | 11-15 | F |
| NHS 64 | 11-15 | F |
| NHS 69 | 6-10 | F |
| NHS 71 | 11-15 | F |
| NHS 82 | 16-20 | F |
| NHS 93 | 6-10 | F |
| NHS 94 | 6-10 | M |

**Figure F1.**
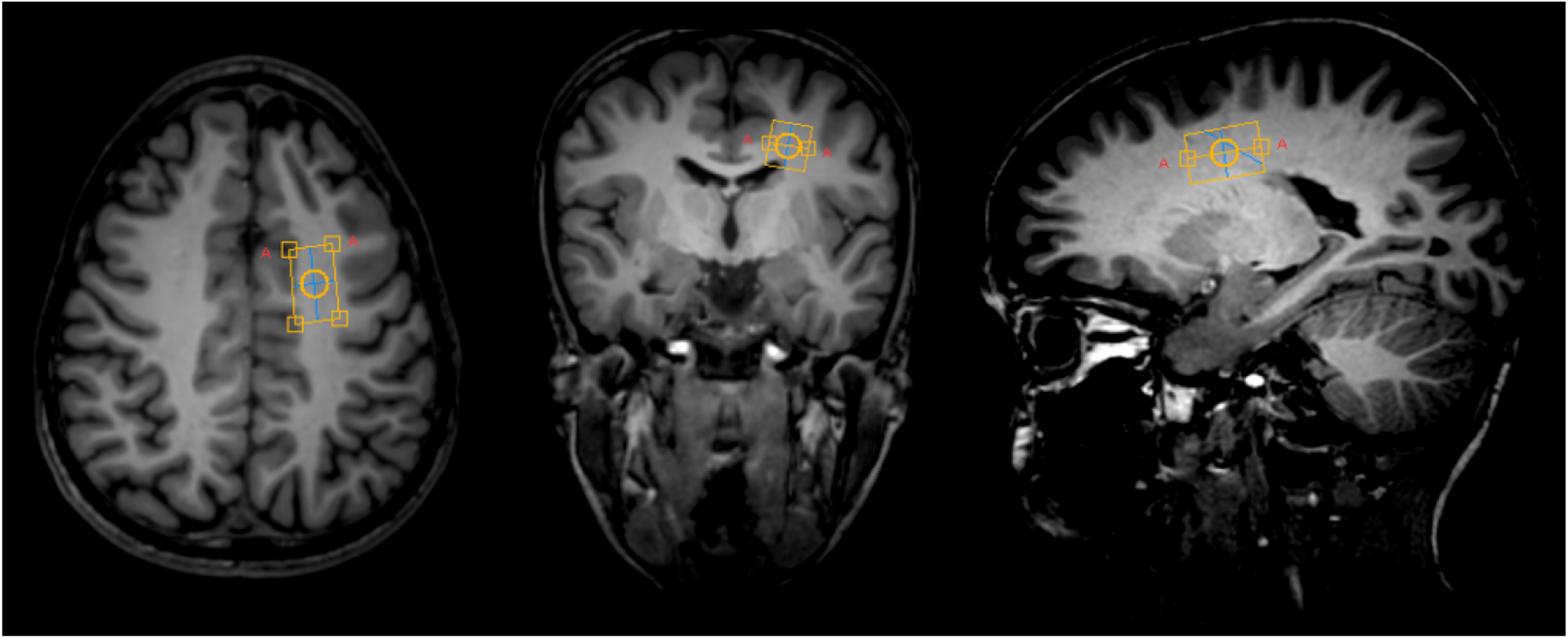
Magnetic resonance spectroscopy left centrum semiovale (LCSO) voxel location in a gene therapy treated GM1 patient.

### Supplementary Results

#### Supplement G Adverse Events

**Table G1:** Cumulative Summary of Serious Adverse Events.

| Participant | Onset | Body System<sup>a</sup> | AE Term<sup>b</sup> | Severity (CTCAE grade)<sup>c</sup> | AE Outcome | Causality to Study Treatment<sup>d</sup> | Causality to Disease Progression<sup>d</sup> | Causality to Immunosuppression regimen<sup>d</sup> | Causality to Protocol-related procedure |
| --- | --- | --- | --- | --- | --- | --- | --- | --- | --- |
| GT03 | Day 18 | Infections and infestations | Bacterial Sepsis | Grade 3 – Severe | Resolved | Not related | Not related | Not related | Definitely Related |
| GT04 | Day 261 | Neurologic | Seizures | Grade 3 Severe | Resolved | Not related | Definitely Related | Not related | Not related |
| GT04 | Day 1081 | Neurologic | Worsening Seizures | Grade 3 Severe | Not resolved | Not related | Definitely Related | Not related | Not related |
| GT17 | Day -22 | Immune system | Anaphylactic reaction | Grade 4 Life threatening | Resolved | Not related | Not related | Definitely Related | Not related |
| GT17 | Day 3 | Gastrointestinal disorders | Vomiting | Grade 3 Severe | Resolved | Definitely related<sup>e</sup> | Not related | Not related | Not related |

<sup>a</sup> MedDRA Dictionary version 27.0
<sup>b</sup> Preferred Term
<sup>c</sup> Common Terminology Criteria for Adverse Events (CTCAE) version 5.0
<sup>d</sup> Based on Investigator assessment
<sup>e</sup> The event was not unexpected

**Table G2:** Cumulative Summary of AE’s in Clinical Study 19-HG-0101.

| **Subject ID** | **Onset** | **AE Term<sup>a</sup>** | **Severity<sup>b</sup>** | **AE Outcome** | **Causality to Study Treatment<sup>c</sup>** |
| --- | --- | --- | --- | --- | --- |
| GT03 | Day -30 | Skin abrasion | Grade 1 Mild | Resolved | Not related |
| GT03 | Day -21 | Elevated triglycerides | Grade 1 Mild | Resolved | Not related |
| GT03 | Day 18 | Bacterial sepsis | Grade 3 Severe | Resolved | Not related |
| GT03 | Day 21 | Iron deficiency anemia | Grade 1 Mild | Resolved | Not related |
| GT03 | Day 93 | Pyrexia | Grade 1 Mild | Resolved | Not related |
| GT03 | Day 186 | Skin laceration | Grade 2 Moderate | Resolved | Not related |
| GT03 | Day 363 | Amblyopia | Grade 2 Moderate | Not resolved | Not related |
| GT03 | Day 363 | Astigmatism | Grade 2 Moderate | Not resolved | Not related |
| GT03 | Day 575 | Presumed COVID-19 Infection | Grade 1 Mild | Resolved | Not related |
| GT03 | Day 1095 | Obstructive sleep apnea | Grade 2 Mild | Not resolved | Not related |
| GT04 | Day -1 | Platelet count decreased | Grade 1 Mild | Resolved | Not related |
| GT04 | Day 2 | Fibrin D dimer increased | Grade 1 Mild | Resolved | Probably related |
| GT04 | Day 3 | Aspartate aminotransferase increased | Grade 3 Severe | Resolved | Possibly related |
| GT04 | Day 90 | Stomatitis | Grade 2 Moderate | Resolved | Not related |
| GT04 | Day 90 | Iron deficiency anemia | Grade 1 Mild | Resolved | Not related |
| GT04 | Day 180 | Stomatitis | Grade 2 Moderate | Resolved | Not related |
| GT04 | Day 261 | Seizures | Grade 3 Severe | Resolved | Not related |
| GT04 | Day 262 | Focal seizures | Grade 3 Severe | Not resolved | Not related |
| GT04 | Day 1081 | Worsening Seizures | Grade 3 Severe | Not resolved | Not related |
| GT06 | Day 7 | Aspartate aminotransferase increased | Grade 1 Mild | Resolved | Possibly related |
| GT06 | Day 30 | Iron deficiency anemia | Grade 1 Mild | Resolved | Not related |
| GT06 | Day 90 | Stomatitis | Grade 1 Mild | Resolved | Not related |
| GT06 | Day 358 | Drug eruption | Grade 1 Mild | Resolved | Not related |
| GT06 | Day 605 | Corona virus infection | Grade 1 Mild | Resolved | Not related |
| GT06 | Day 672 | Gastrointestinal virus | Grade 1 Mild | Resolved | Not related |
| GT06 | Day 728 | Activated partial thromboplastin time prolonged | Grade 1 Mild | Resolved | Not related |
| GT06 | Day 734 | Obstructive sleep apnea | Grade 1 Mild | Resolved | Not related |
| GT07 | Day -7 | Complication associated with device | Grade 1 Mild | Resolved | Not related |
| GT07 | Day 15 | Aspartate aminotransferase increased | Grade 1 Mild | Resolved | Possibly related |
| GT07 | Day 30 | Iron deficiency anemia | Grade 1 Mild | Resolved | Not related |
| GT07 | Day 184 | Vomiting | Grade 2 Moderate | Resolved | Not related |
| GT07 | Day 180 | Low Vitamin D | Grade 1 Mild | Resolved | Not related |
| GT07 | Day 605 | Corona virus infection | Grade 1 Mild | Resolved | Not related |
| GT07 | Day 672 | Gastrointestinal virus | Grade 1 Mild | Resolved | Not related |
| GT07 | Day 728 | Activated partial thromboplastin time prolonged | Grade 1 Mild | Resolved | Not related |
| GT07 | Day 728 | Obstructive sleep apnea | Grade 3 Severe | Resolved | Not related |
| GT08 | Day -21 | Pyrexia | Grade 1 Mild | Resolved | Not related |
| GT08 | Day 0 | Tachycardia | Grade 1 Mild | Resolved | Not related |
| GT08 | Day 0 | Hypertension | Grade 3 Severe | Resolved | Not related |
| GT08 | Day 7 | Low Vitamin D | Grade 1 Mild | Resolved | Not related |
| GT08 | Day 15 | Aspartate aminotransferase increased | Grade 1 Mild | Resolved | Possibly related |
| GT08 | Day 31 | Fibrin D dimer increased | Grade 1 Mild | Resolved | Probably related |
| GT08 | Day 53 | Stomatitis | Grade 2 Moderate | Resolved | Not related |
| GT08 | Day 75 | Iron deficiency anemia | Grade 1 Mild | Resolved | Not related |
| GT08 | Day 88 | Stomatitis | Grade 2 Moderate | Resolved | Not related |
| GT08 | Day 605 | Corona virus infection | Grade 1 Mild | Resolved | Not related |
| GT08 | Day 672 | Gastrointestinal virus | Grade 1 Mild | Resolved | Not related |
| GT08 | Day 728 | Activated partial thromboplastin time prolonged | Grade 1 Mild | Resolved | Not related |
| GT08 | Day 728 | Leukocytosis | Grade 1 Mild | Resolved | Not related |
| GT08 | Day 728 | Fibrin D dimer increased | Grade 1 Mild | Resolved | Not related |
| GT08 | Day 735 | Upper respiratory infection | Grade 1 Mild | Resolved | Not related |
| GT08 | Day 738 | Obstructive sleep apnea | Grade 3 Mild | Resolved | Not related |
| GT10 | Day -14 | Blood cholesterol increased | Grade 1 Mild | Resolved | Not related |
| GT10 | Day -14 | Neutrophil count decreased | Grade 2 Moderate | Resolved | Not related |
| GT10 | Day -14 | Hypertriglyceridemia | Grade 1 Mild | Resolved | Not related |
| GT10 | Day 1 | Tachycardia | Grade 1 Mild | Resolved | Possibly related |
| GT10 | Day 2 | Decreased appetite | Grade 2 Moderate | Resolved | Probably related |
| GT10 | Day 4 | Retching | Grade 2 Moderate | Resolved | Probably related |
| GT10 | Day 4 | Vomiting | Grade 2 Moderate | Resolved | Probably related |
| GT10 | Day 4 | Alanine aminotransferase increased | Grade 1 Mild | Resolved | Probably related |
| GT10 | Day 7 | Elevated Ferritin | Grade 1 Mild | Resolved | Probably related |
| GT10 | Day 22 | Neutrophil count decreased | Grade 3 Severe | Resolved | Not related |
| GT10 | Day 180 | Vomiting | Grade 2 Moderate | Resolved | Not related |
| GT10 | Day 180 | Pyrexia | Grade 1 Mild | Resolved | Not related |
| GT10 | Day 365 | Hypertriglyceridemia | Grade 1 Mild | Resolved | Not related |
| GT10 | Day 635 | Corona virus infection | Grade 1 Mild | Resolved | Not related |
| GT10 | Day 1095 | Hypercapnia during sleep study | Grade 1 Mild | Not resolved | Not related |
| GT11 | Day -14 | Neutrophil count decreased | Grade 3 Severe | Resolved | Not related |
| GT11 | Day 2 | Vomiting | Grade 2 Moderate | Resolved | Probably related |
| GT11 | Day 2 | Decreased appetite | Grade 2 Moderate | Resolved | Probably related |
| GT11 | Day 2 | Device occlusion | Grade 2 Moderate | Resolved | Not related |
| GT11 | Day 3 | Constipation | Grade 1 Mild | Resolved | Not related |
| GT11 | Day 3 | Aspartate aminotransferase increased | Grade 1 Mild | Resolved | Probably related |
| GT11 | Day 5 | Neutrophil count decreased | Grade 3 Severe | Resolved | Unlikely related |
| GT11 | Day 7 | Catheter site related reaction | Grade 1 Mild | Resolved | Not related |
| GT11 | Day 7 | Iron deficiency anemia | Grade 1 Mild | Resolved | Not related |
| GT11 | Day 7 | Hypogammaglobulinemia | Grade 1 Mild | Resolved | Not related |
| GT11 | Day 30 | Low Vitamin D | Grade 1 Mild | Resolved | Not related |
| GT11 | Day 51 | Aspartate aminotransferase increased | Grade 1 Mild | Resolved | Probably related |
| GT11 | Day 63 | Fibrin D dimer increased | Grade 1 Mild | Resolved | Possibly related |
| GT11 | Day 90 | Iron deficiency anemia | Grade 1 Mild | Resolved | Not related |
| GT11 | Day 90 | Neutrophil count decrease | Grade 1 Mild | Resolved | Not related |
| GT11 | Day 180 | Vomiting | Grade 1 Mild | Resolved | Not related |
| GT11 | Day 270 | Vomiting | Grade 1 Mild | Resolved | Not related |
| GT11 | Day 309 | Obstructive sleep apnea | Grade 2 Moderate | Resolved | Not related |
| GT11 | Day 605 | Corona virus infection | Grade 1 Mild | Resolved | Not related |
| GT12 | Day -28 | Headache | Grade 1 - Mild | Resolved | Not related |
| GT12 | Day -28 | Vomiting | Grade 2 - Moderate | Resolved | Not related |
| GT12 | Day -24 | Headache | Grade 1 - Mild | Resolved | Not related |
| GT12 | Day -23 | Vomiting | Grade 2 - Moderate | Resolved | Not related |
| GT12 | Day -14 | Neutrophil count decreased | Grade 2 - Moderate | Resolved | Not related |
| GT12 | Day 0 | Neutrophil count decreased | Grade 2 - Moderate | Resolved | Not related |
| GT12 | Day 2 | Fibrin D dimer increased | Grade 1 - Mild | Resolved | Probably related |
| GT12 | Day 2 | Serum ferritin increased | Grade 1 - Mild | Resolved | Probably related |
| GT12 | Day 2 | Vomiting | Grade 2 - Moderate | Resolved | Possibly related |
| GT12 | Day 7 | Aspartate aminotransferase increased | Grade 1 - Mild | Resolved | Probably related |
| GT12 | Day 14 | Neutrophil count decreased | Grade 3 - Severe | Resolved | Not related |
| GT12 | Day 255 | Corona virus infection | Grade 1 Mild | Resolved | Not related |
| GT12 | Day 545 | Upper respiratory infection | Grade 1 Mild | Resolved | Not related |
| GT17 | Day -22 | Anaphylactic reaction | Grade 4 Life threatening | Resolved | Not related |
| GT17 | Day -20 | Skin abrasion | Grade 1 Mild | Resolved | Not related |
| GT17 | Day 0 | PICC line occlusion | Grade 2 Moderate | Resolved | Not related |
| GT17 | Day 1 | Premature ventricular contractions | Grade 1 Mild | Resolved | Not related |
| GT17 | Day 2 | Vomiting | Grade 2 Moderate | Resolved | Definitely related |
| GT17 | Day 3 | Vomiting | Grade 3 Severe | Resolved | Definitely related |
| GT17 | Day 3 | Aspartate aminotransferase increased | Grade 1 Mild | Resolved | Definitely related |
| GT17 | Day 3 | Fibrin D dimer increased | Grade 1 Mild | Resolved | Definitely related |
| GT17 | Day 3 | Ferritin increased | Grade 1 Mild | Resolved | Definitely related |
| GT17 | Day 3 | C-reactive protein increased | Grade 1 Mild | Resolved | Definitely related |
| GT17 | Day 8 | PICC line flush resistance | Grade 2 Moderate | Resolved | Not related |
| GT17 | Day 10 | Gastrointestinal virus | Grade 3 Severe | Resolved | Not related |
| GT17 | Day 14 | Neutrophil count decrease | Grade 3 Severe | Resolved | Not related |
| GT17 | Day 14 | Elispot positive | Grade 2 Moderate | Resolved | Definitely related |
| GT17 | Day 21 | Hypertriglyceridemia | Grade 1 Mild | Resolved | Not related |
| GT17 | Day 86 | Aspartate aminotransferase increased | Grade 1 Mild | Resolved | Possibly related |
| GT17 | Day 86 | Neutrophil count decrease | Grade 2 Moderate | Resolved | Not related |
| GT17 | Day 97 | Neutrophil count decrease | Grade 3 Severe | Resolved | Not related |
| GT17 | Day 97 | Alanine aminotransferase increased | Grade 3 Severe | Resolved | Possibly related |

<sup>a</sup>Common Terminology Criteria for Adverse Events (CTCAE, v5.0)
<sup>b</sup>Scored according to Common Terminology Criteria for Adverse Events (CTCAE) version 5.0.
<sup>c</sup>As determined by the principal investigator.

#### Supplement H Clinical Laboratory Studies

##### Aspartate Aminotransferase (AST)

**Figure H1.**
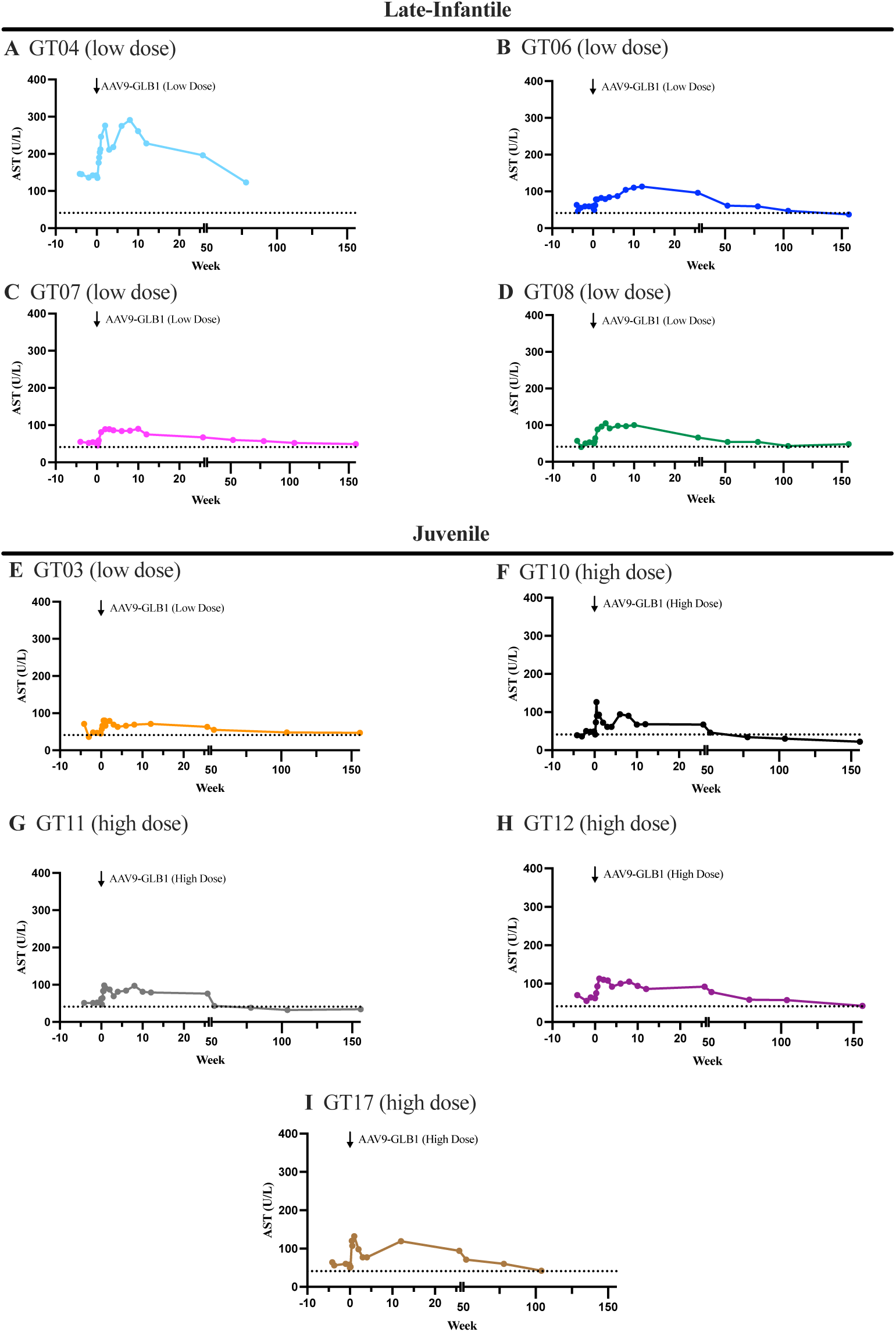
Aspartate aminotransferase levels (AST) in serum for each patient over the duration of the study. AAV-GLB1 gene transfer was completed on week 0 for each patient. The dotted line on each graph represents the upper limit of normal for AST (41 U/L).

##### Alanine Aminotransferase (ALT)

**Figure H2.**
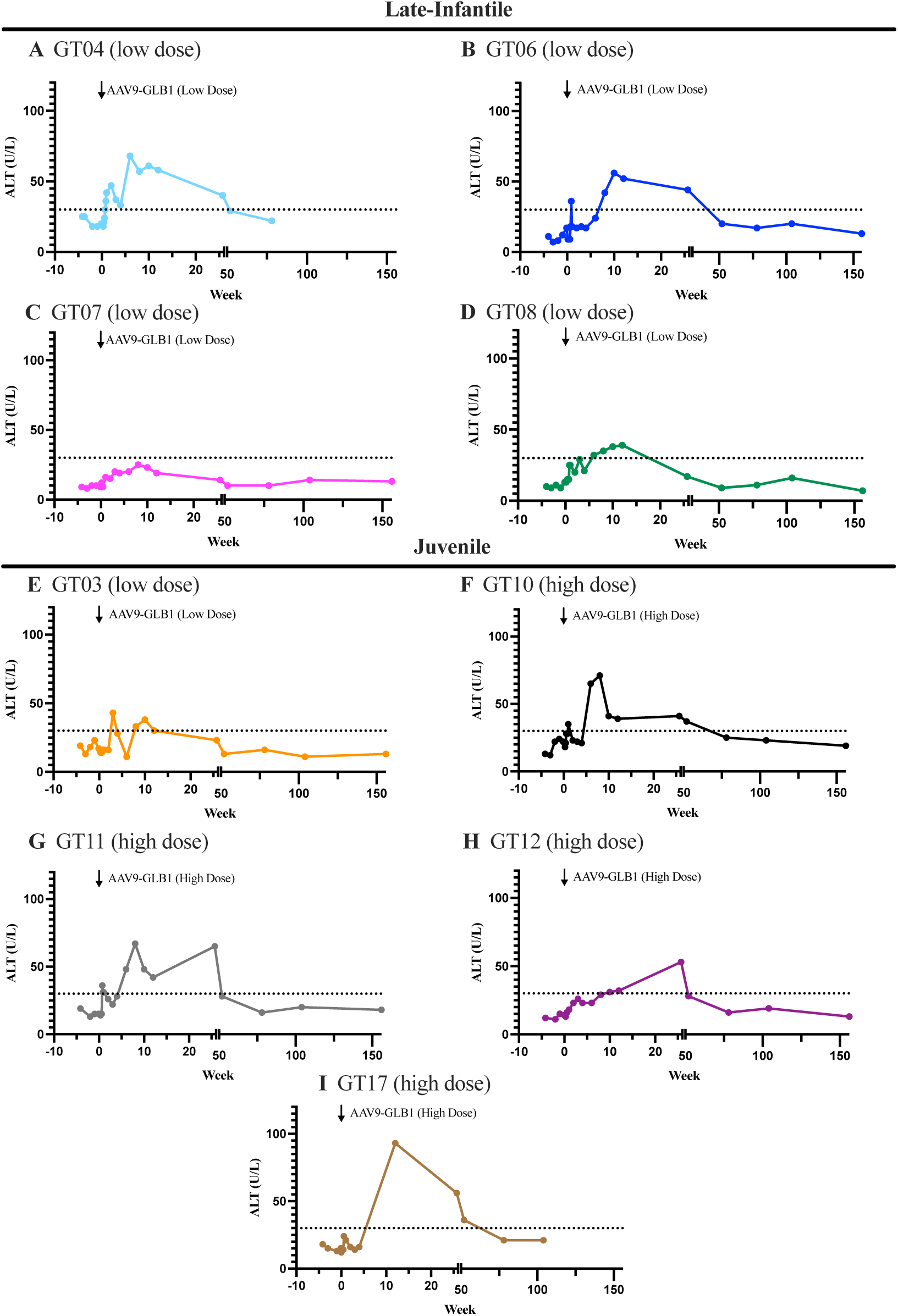
Alanine aminotransferase levels (ALT) in serum for each patient over the duration of the study. AAV-GLB1 gene transfer was completed on week 0 for each patient. The dotted line on each graph represents the upper limit of normal for ALT (30 U/L).

##### Gamma Glutamyl Transferase (GGT)

**Figure H3.**
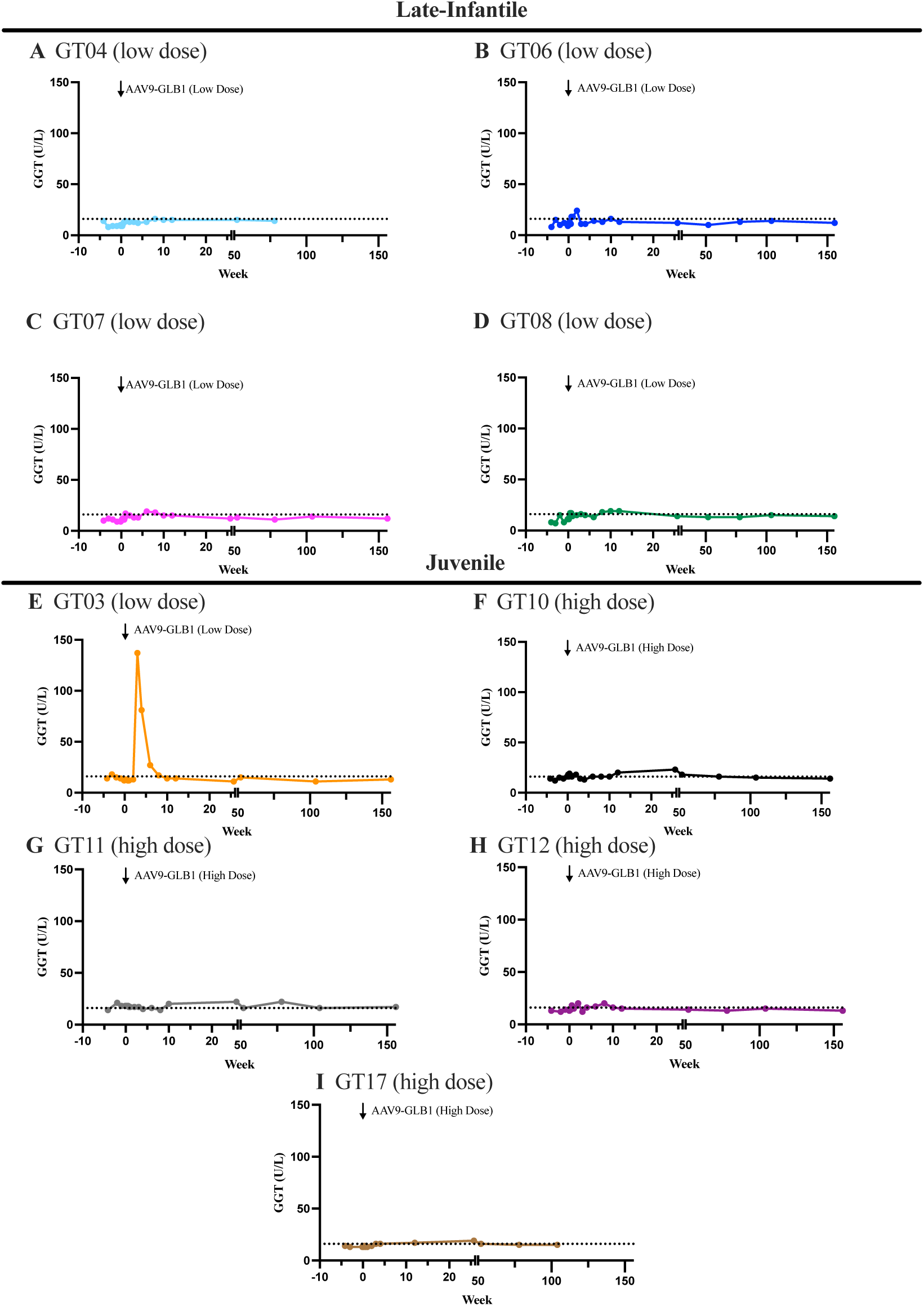
Gamma glutamyl transferase levels (GGT) in serum for each patient over the duration of the study. AAV-GLB1 gene transfer was completed on week 0 for each patient. The dotted line on each graph represents the upper limit of normal for GGT (16 U/L).

##### C3

**Figure H4.**
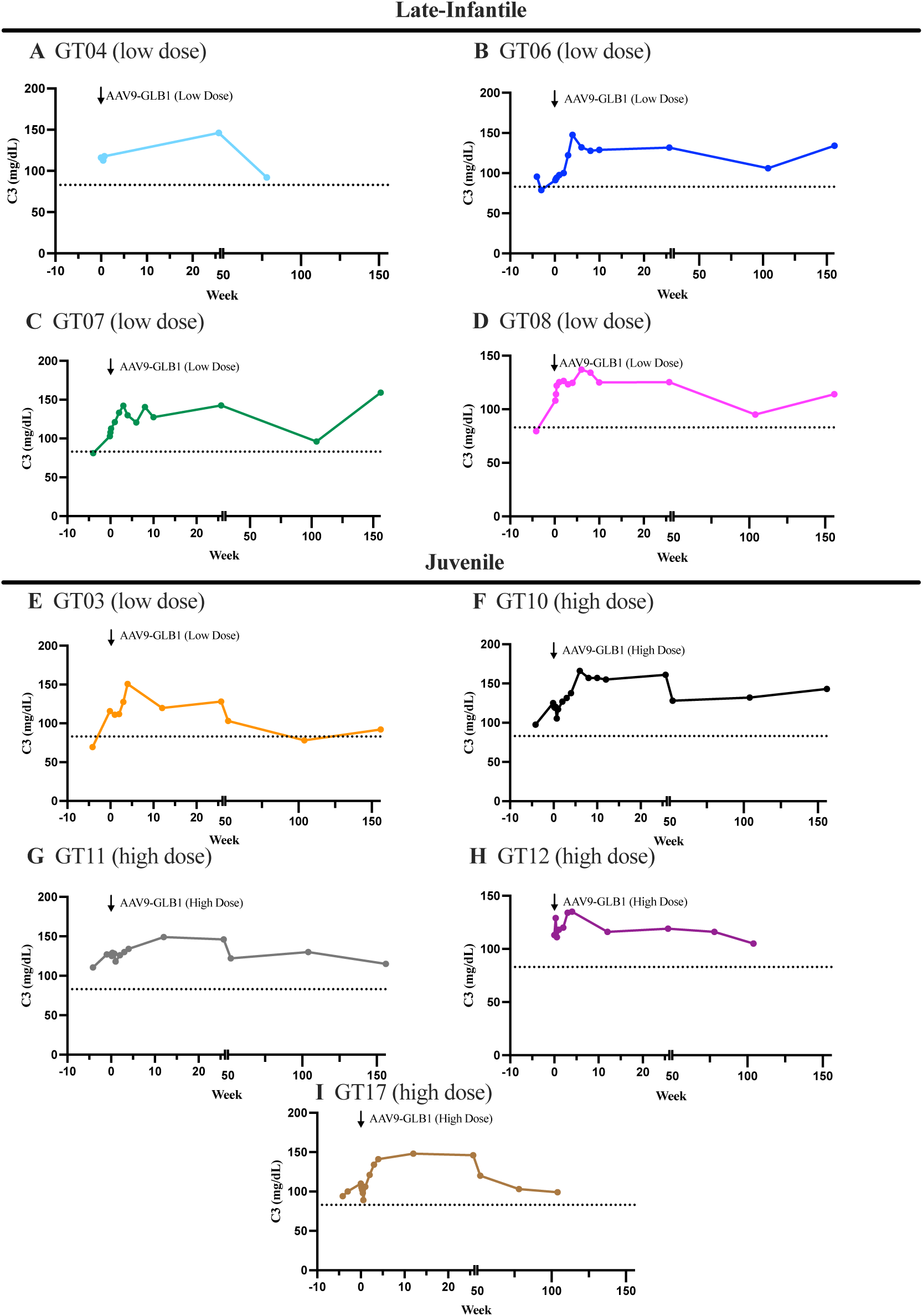
C3 levels in serum for each patient over the duration of the study. AAV-GLB1 gene transfer was completed on week 0 for each patient. The dotted line on each graph represents the lower limit of normal for C3 (83 mg/dL).

##### C4

**Figure H5.**
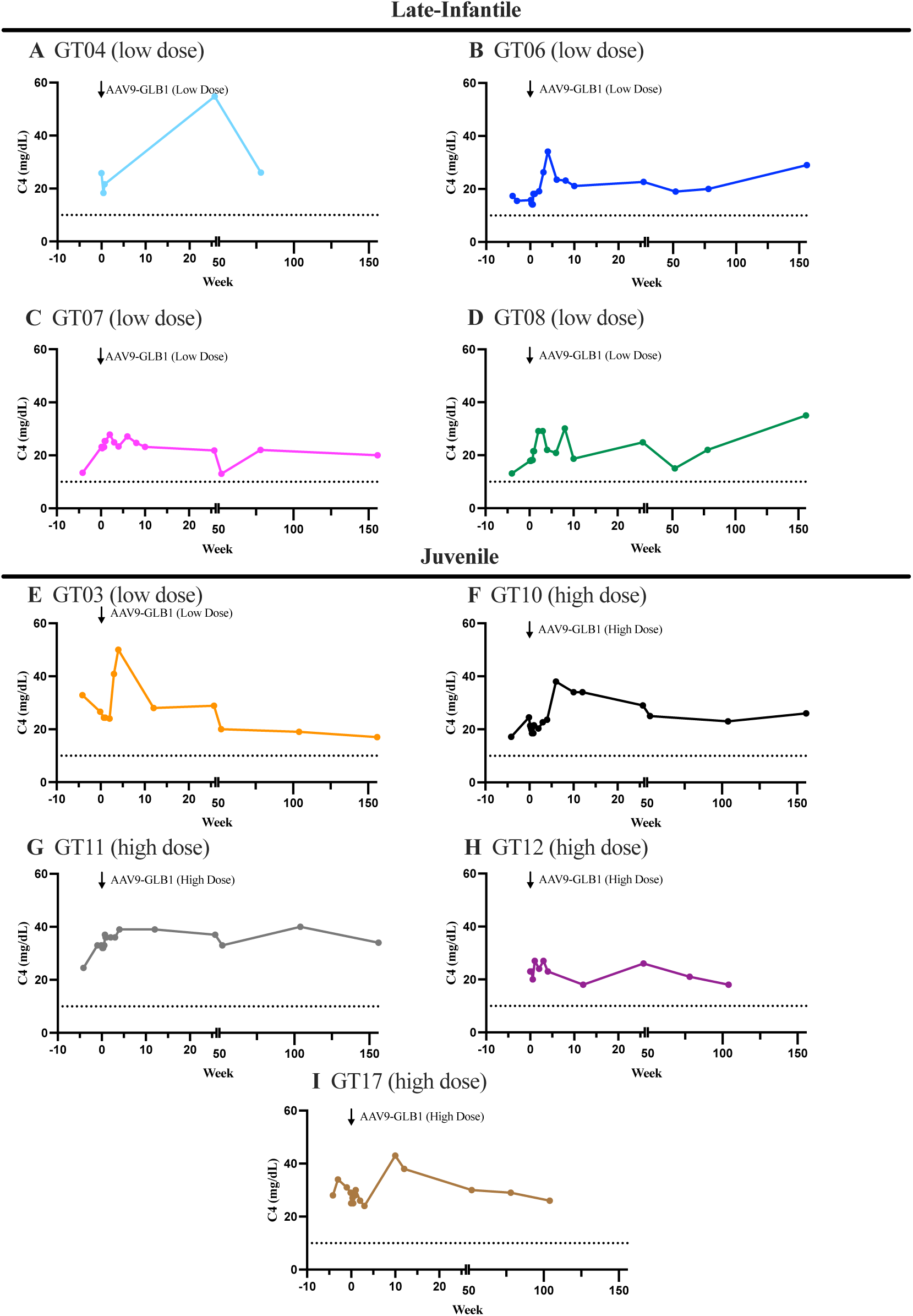
C4 levels in serum for each patient over the duration of the study. AAV-GLB1 gene transfer was completed on week 0 for each patient. The dotted line on each graph represents the lower limit of normal for C4 (10 mg/dL).

##### Platelet

**Figure H6.**
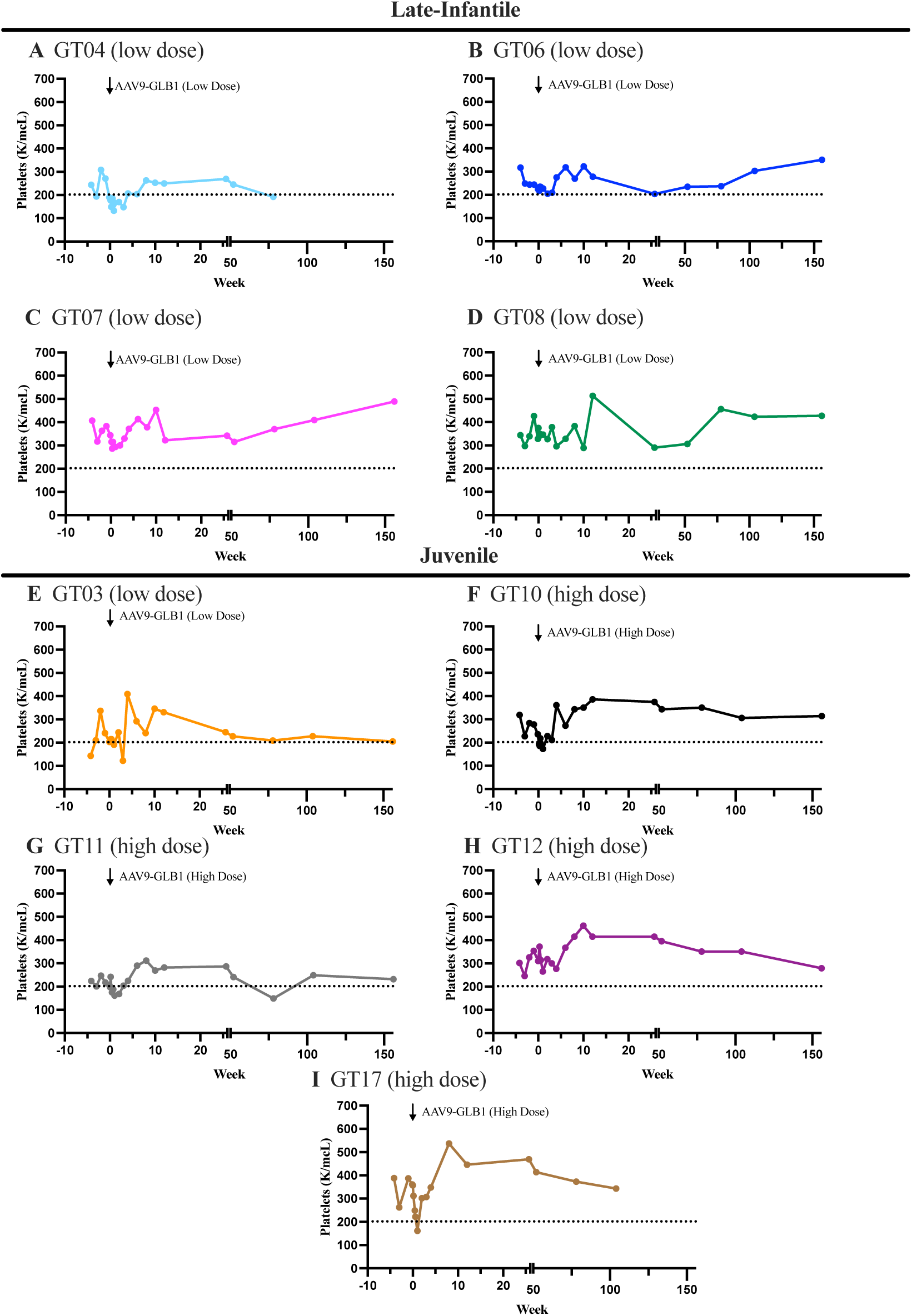
Platelet levels in serum for each patient over the duration of the study. AAV-GLB1 gene transfer was completed on week 0 for each patient. The dotted line on each graph represents the lower limit of normal for Platelets (202 K/mcL).

##### D-Dimer

**Figure H7.**
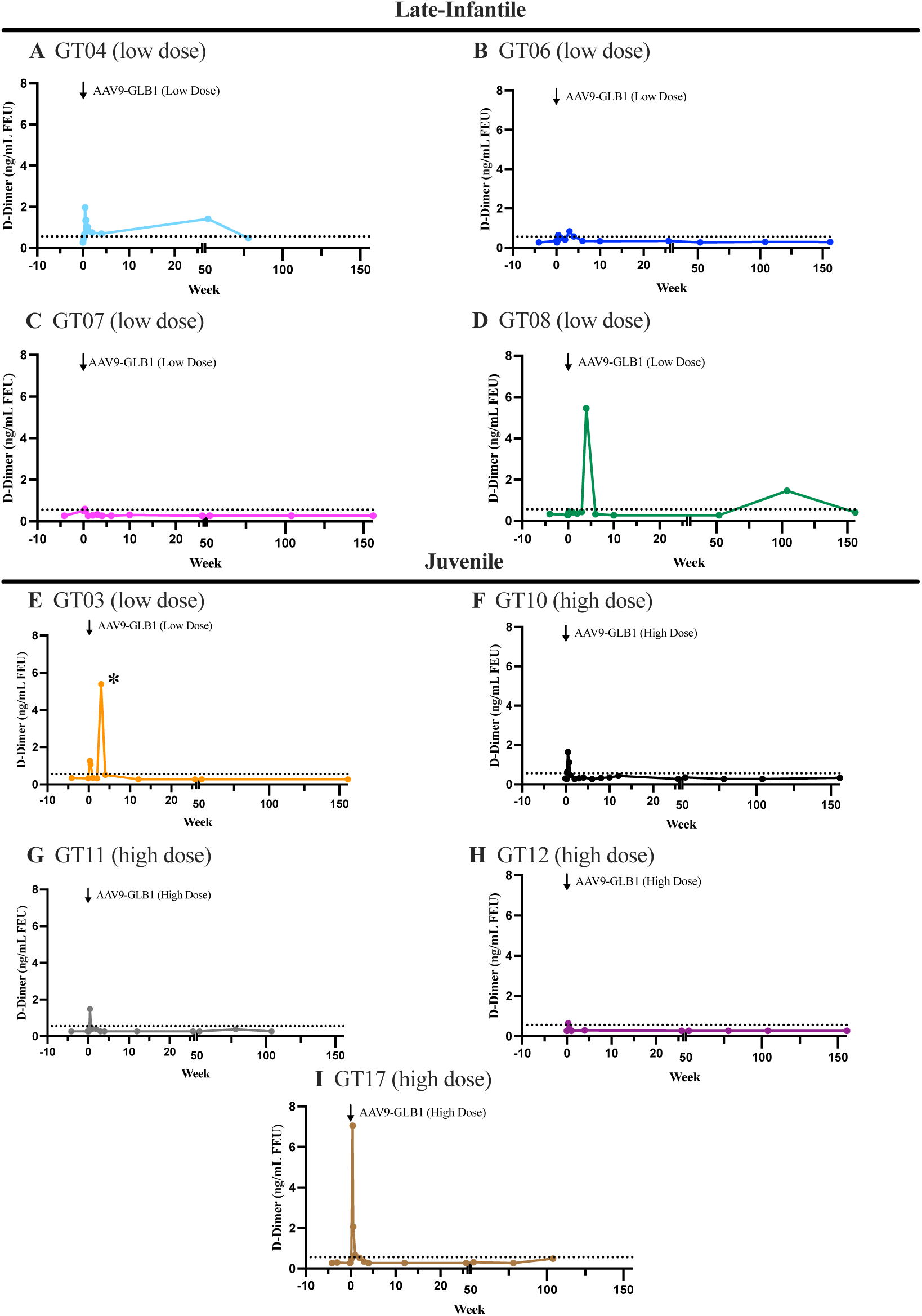
D-dimer levels in serum for each patient over the duration of the study. AAV-GLB1 gene transfer was completed on week 0 for each patient. The dotted line on each graph represents the upper limit of normal for D-dimer (16 U/L). The * for GT03 designates the D-dimer increase associated with central line sepsis.

#### Supplement I Viral Shedding

The overall goal of our viral shedding studies was to determine how long affected siblings should be separated to prevent early exposure to the virus. In our studies, feces contained the highest number of vector genome copies as shown in Figure J2. Ultimately, it was found that in all three media, viral shedding was complete around day 30. This indicates that caution should be taken by both medical staff and patient families until this pointto prevent sensitizing exposure to the viral capsid.

A dose response was observed in the Urine Viral shedding analysis (Figure I1), and there was not sufficient data to visualize a dose response in Feces or Saliva (Figures I2 & I3).

**Figure I1.**
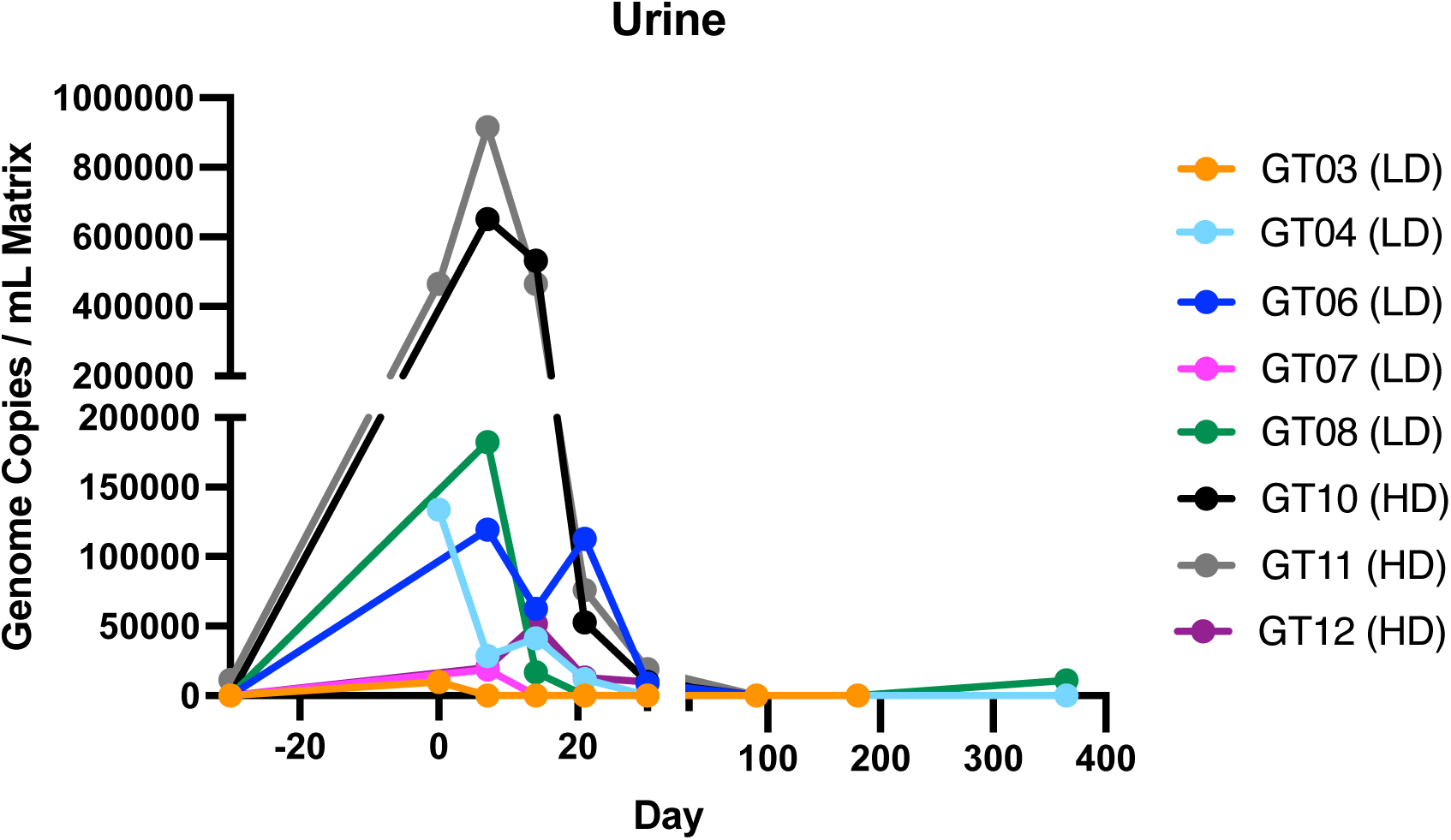
Viral Shedding in Urine. LD corresponds to participants who received the low dose (1.5×10^13^ vector genomes (vg) per kilogram of body weight [vg/kg]) administration of AAV9-GLB1 and HD corresponds to participants who received the high dose administration of AAV9-GLB1 (4.5×10^13^ vg/kg). Results are shown in Genome Copies per milliliter of the original matrix (Urine).

**Figure I2.**
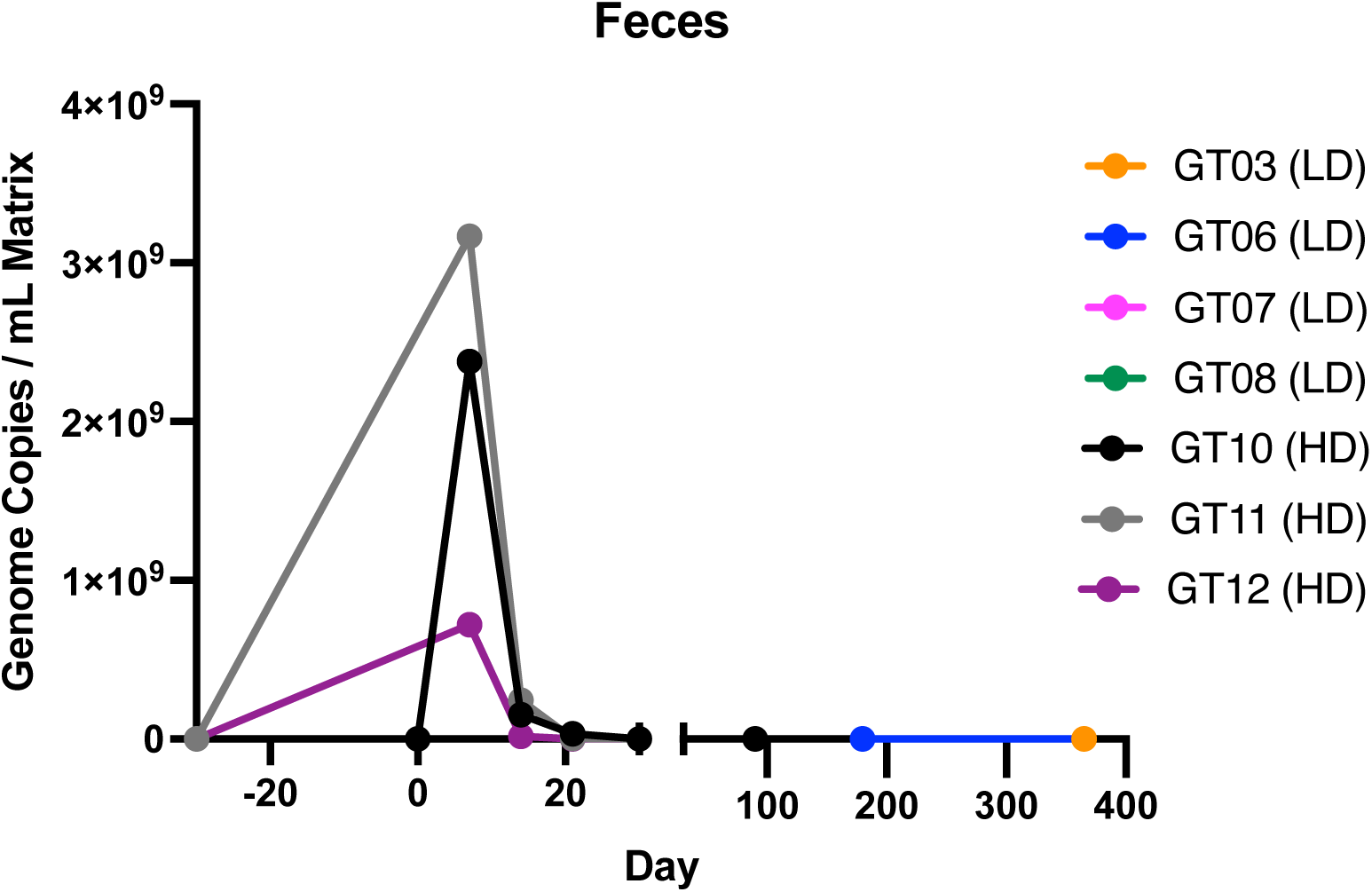
Viral Shedding in Feces. LD corresponds to participants who received the low dose (1.5×10^13^ vector genomes (vg) per kilogram of body weight [vg/kg]) administration of AAV9-GLB1 and HD corresponds to participants who received the high dose administration of AAV9-GLB1 (4.5×10^13^ vg/kg). Results are shown in Genome Copies per milliliter of the original matrix (Feces).

**Figure I3.**
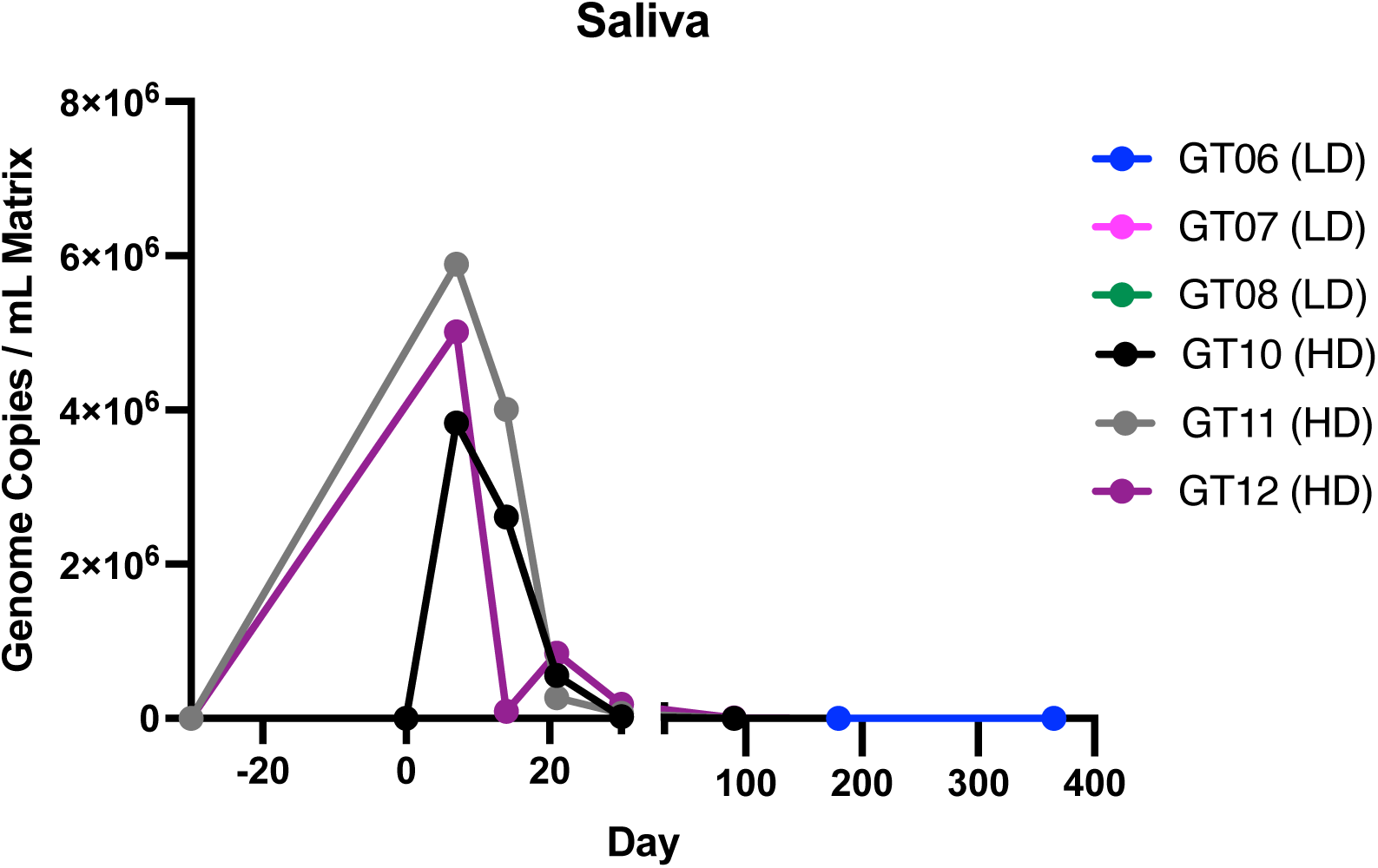
Viral Shedding in Saliva. LD corresponds to participants who received the low dose (1.5×10^13^ vector genomes (vg) per kilogram of body weight [vg/kg]) administration of AAV9-GLB1 and HD corresponds to participants who received the high dose administration of AAV9-GLB1 (4.5×10^13^ vg/kg). Results are shown in Genome Copies per milliliter of the original matrix (Saliva).

**Figure I4.**
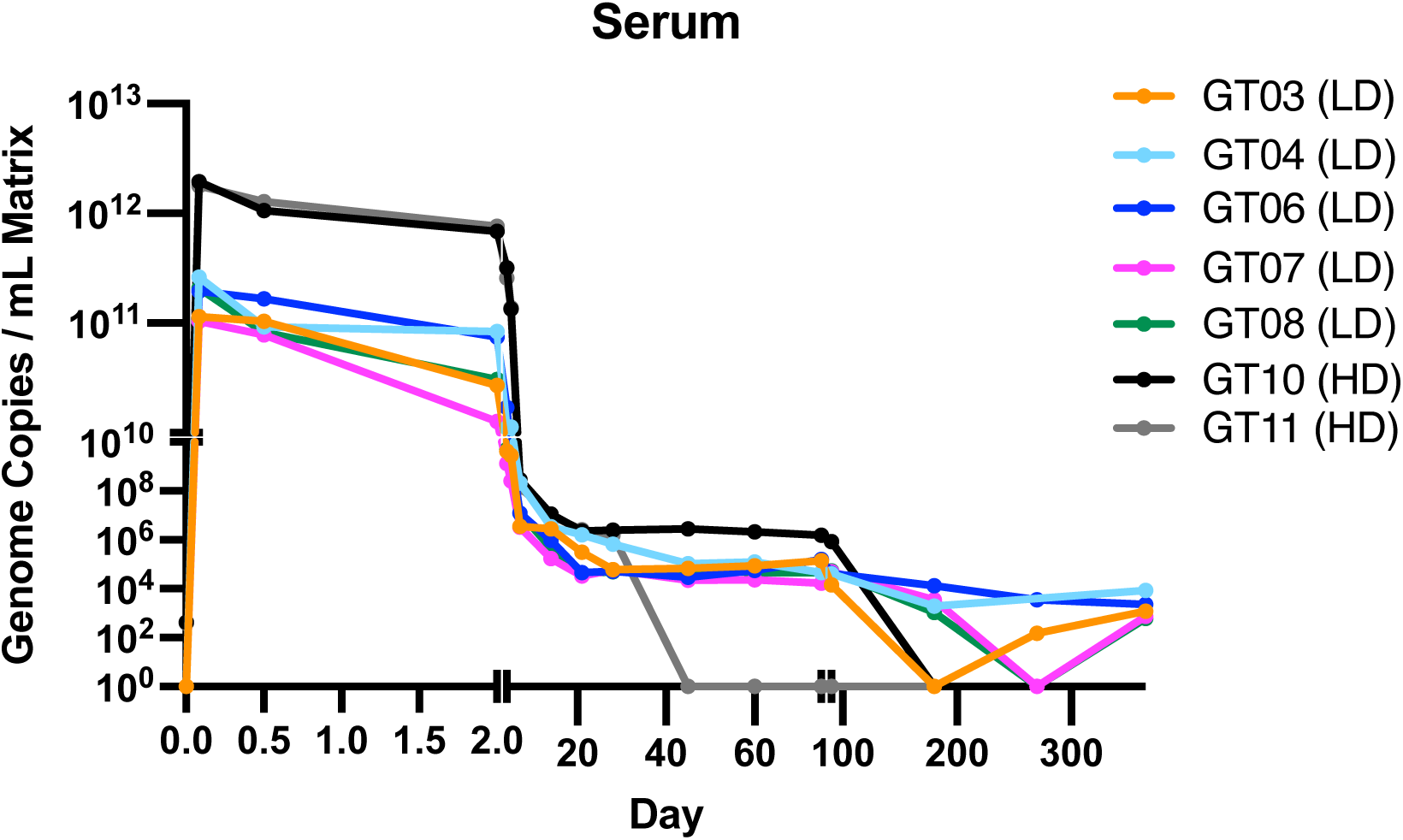
Viral Shedding in Serum. LD corresponds to participants who received the low dose (1.5×10^13^ vector genomes (vg) per kilogram of body weight [vg/kg]) administration of AAV9-GLB1 and HD corresponds to participants who received the high dose administration of AAV9-GLB1 (4.5×10^13^ vg/kg). Results are shown in Genome Copies per milliliter of the original matrix (Serum).

#### Supplement J Immune Response to the Vector

##### Anti-AAV9 IgG

**Figure J1.**
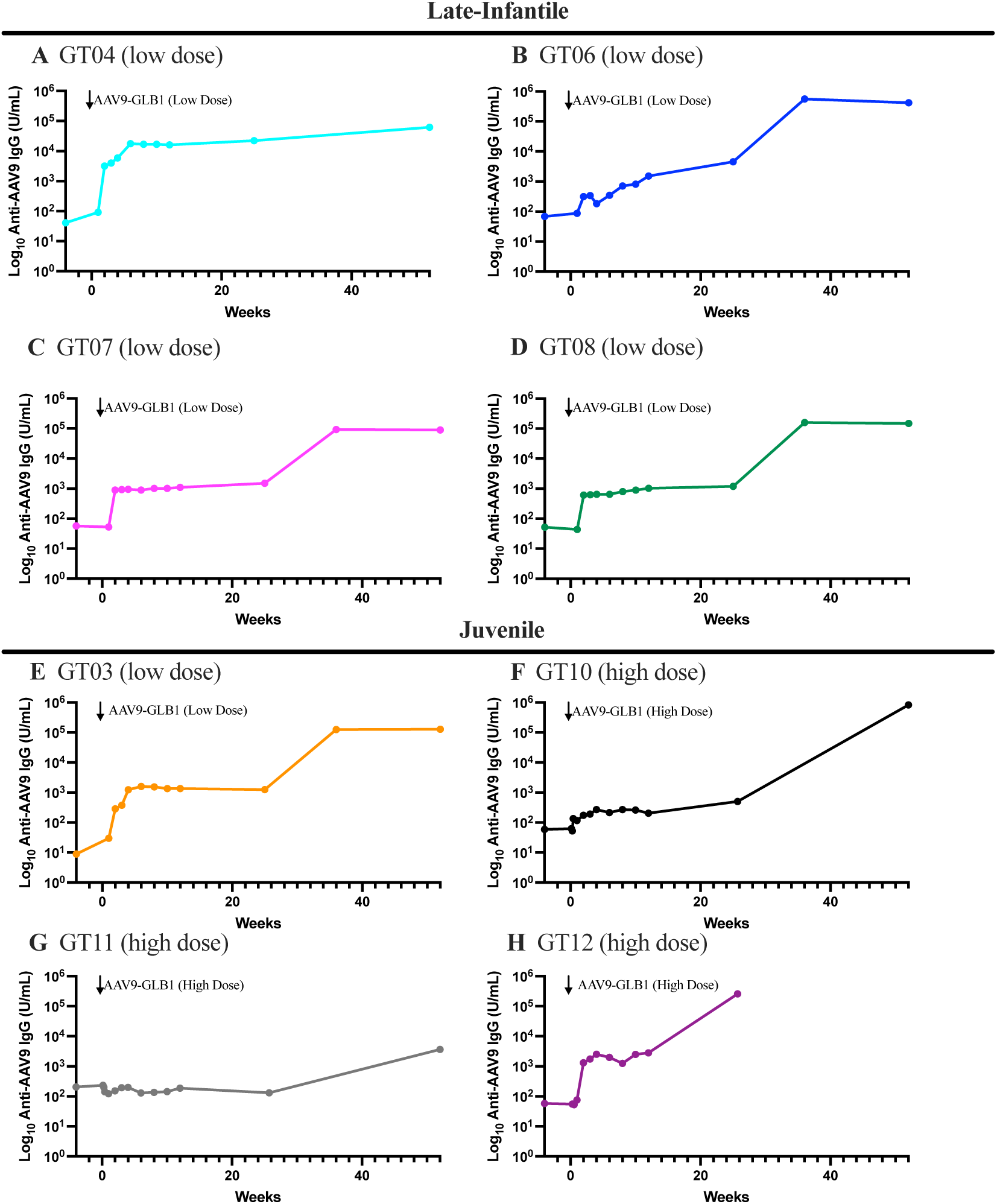
Anti-AAV9 IgG levels in serum.

##### Anti-AAV9 IgM

**Figure J2.**
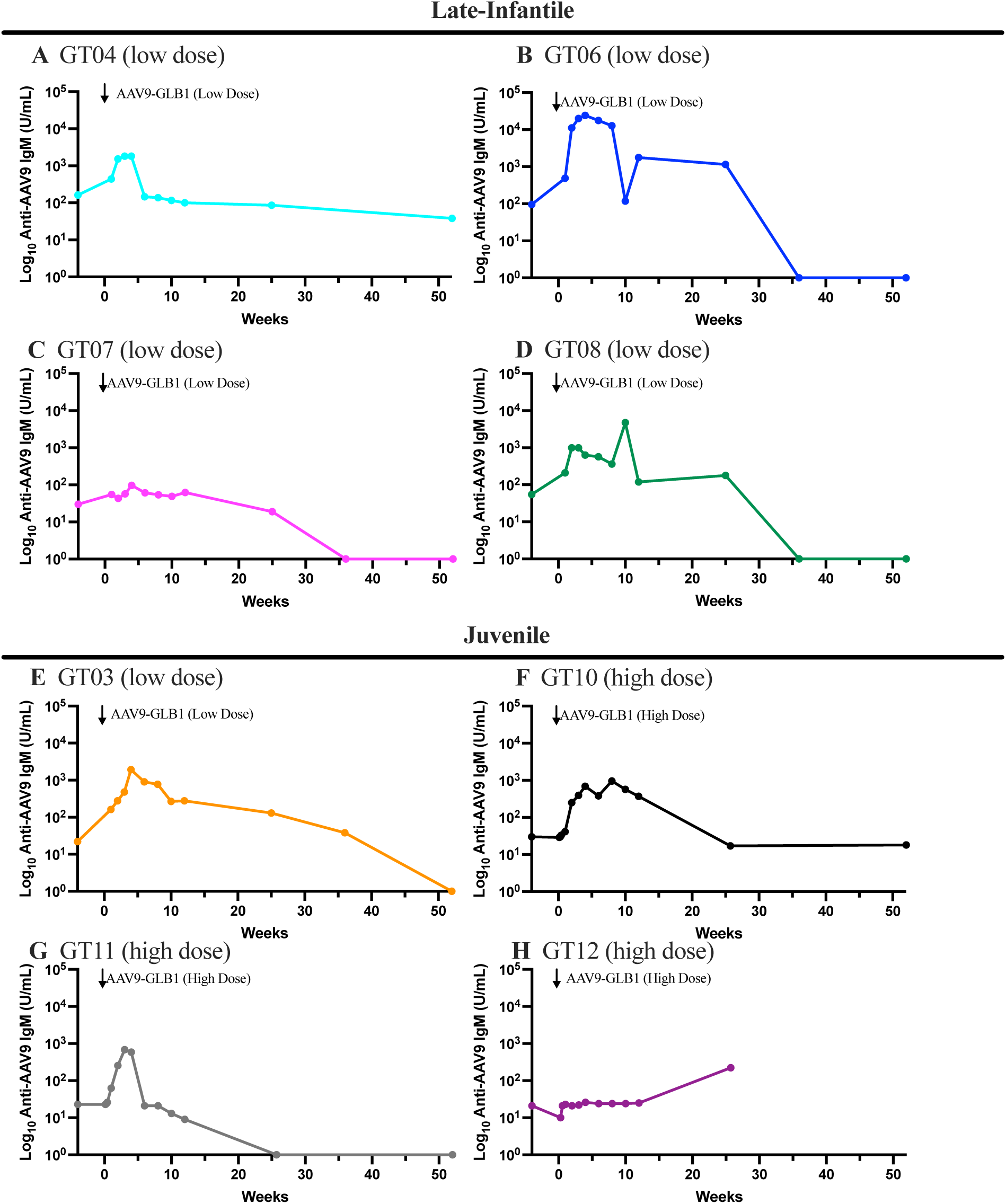
Anti-AAV9 IgM levels in serum.

**Figure J3.**
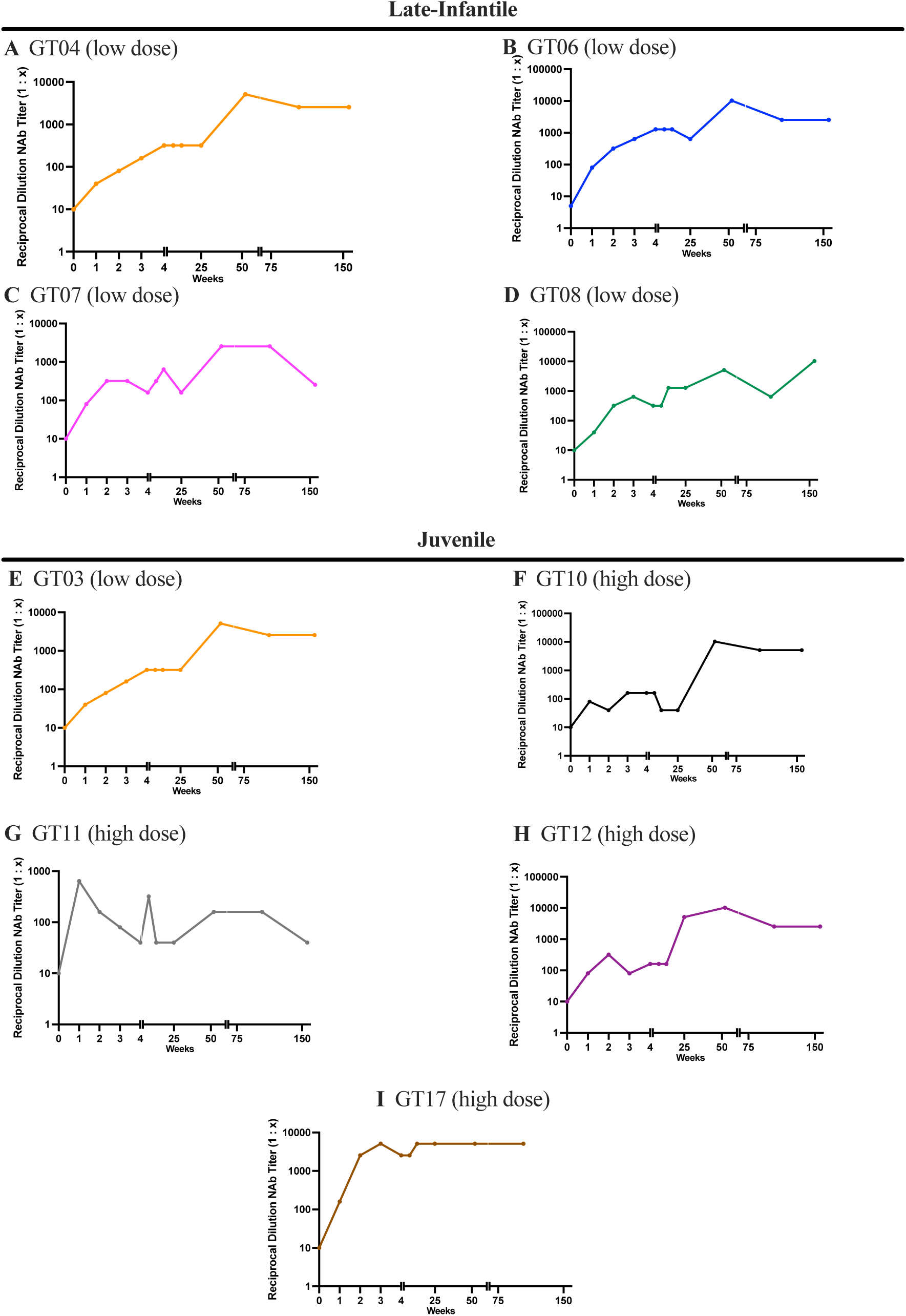
Neutralizing antibodies for all participants over time.

**Figure J4.**
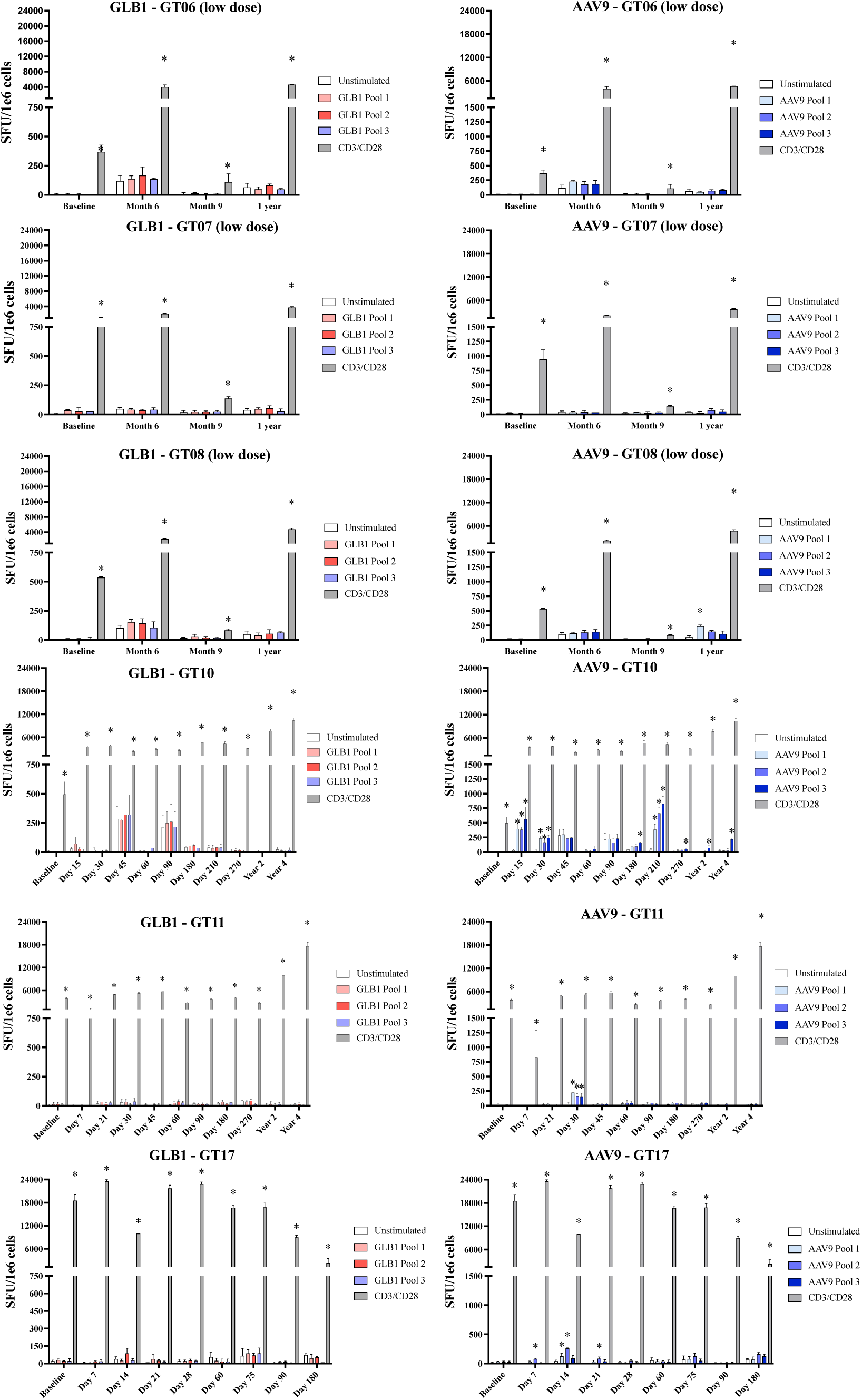
Transgene and Capsid specific ELISpots: IFN-γ ELISpot results for transgene specific (left) and capsid specific (right) immune responses. Positive responses designated by * and determined if they were ≥ 50 SFU/1e6 cells and at least 3X unstimulated negative control. CD3/CD28 stimulation was used as positive control. Positive and negative controls were run on all participants at all timepoints.

#### Supplement K Individualized β-galactosidase, GM1 Ganglioside, and H3N2B Levels

**Figure K1.**
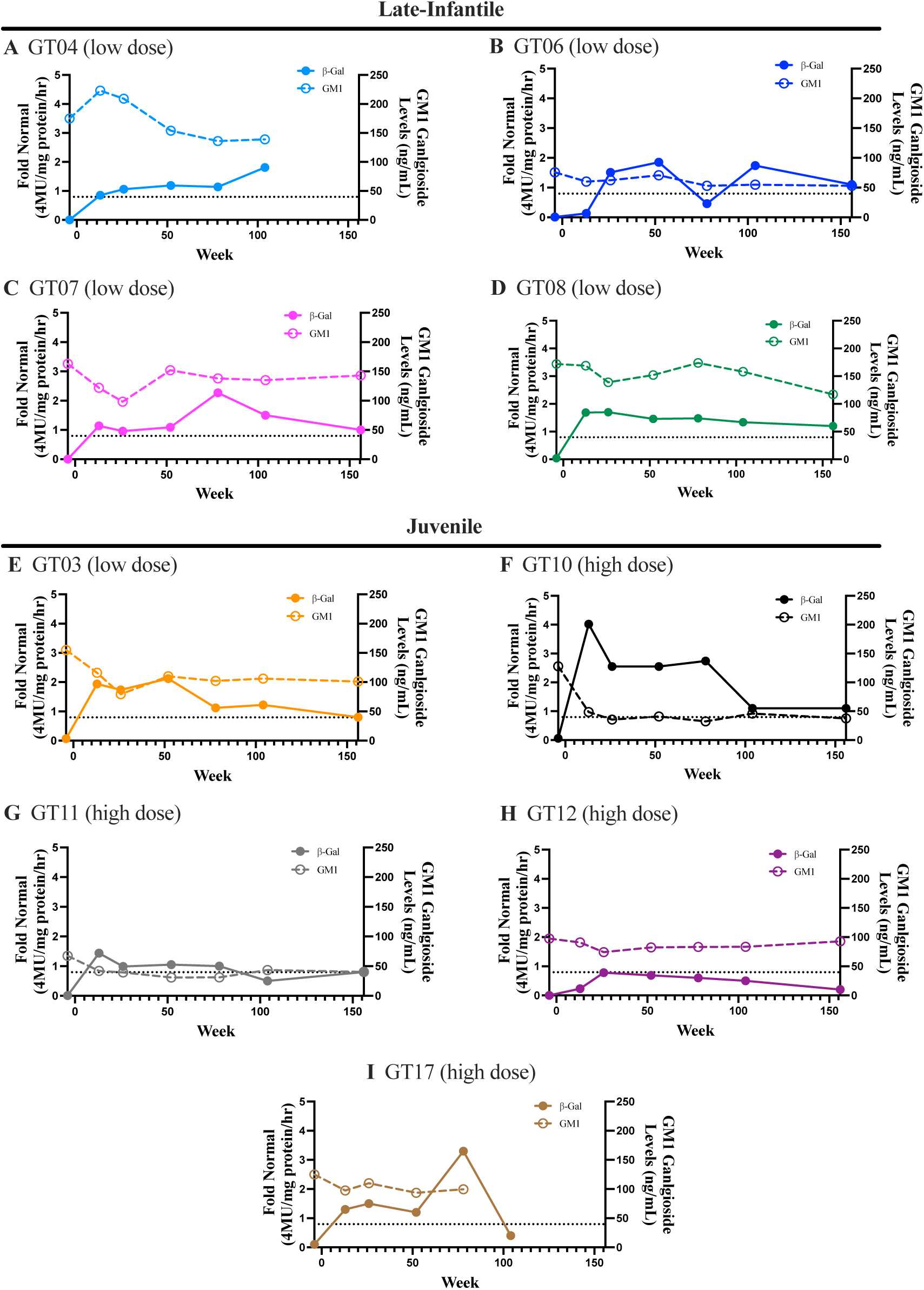
β-galactosidase (closed circles) and GM1 levels (open circles) in CSF. The dotted line on each graph represents the upper limit of normal for GM1 ganglioside levels in CSF (39.9 ng/mL at 1 standard deviation above normal).^37^

**Figure K2.**
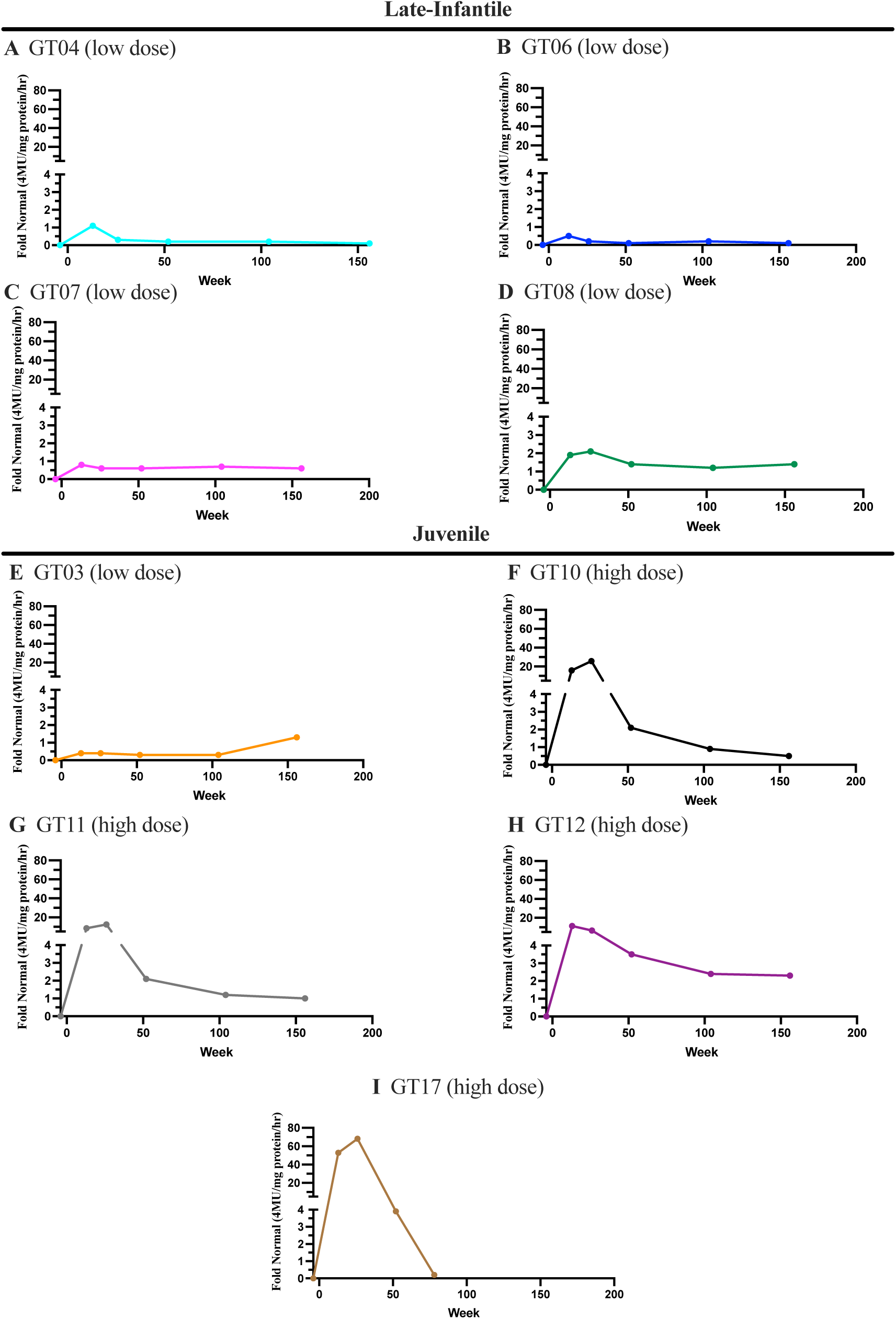
Serum β-galactosidase Levels for Both Dose Groups.

**Figure K3.**
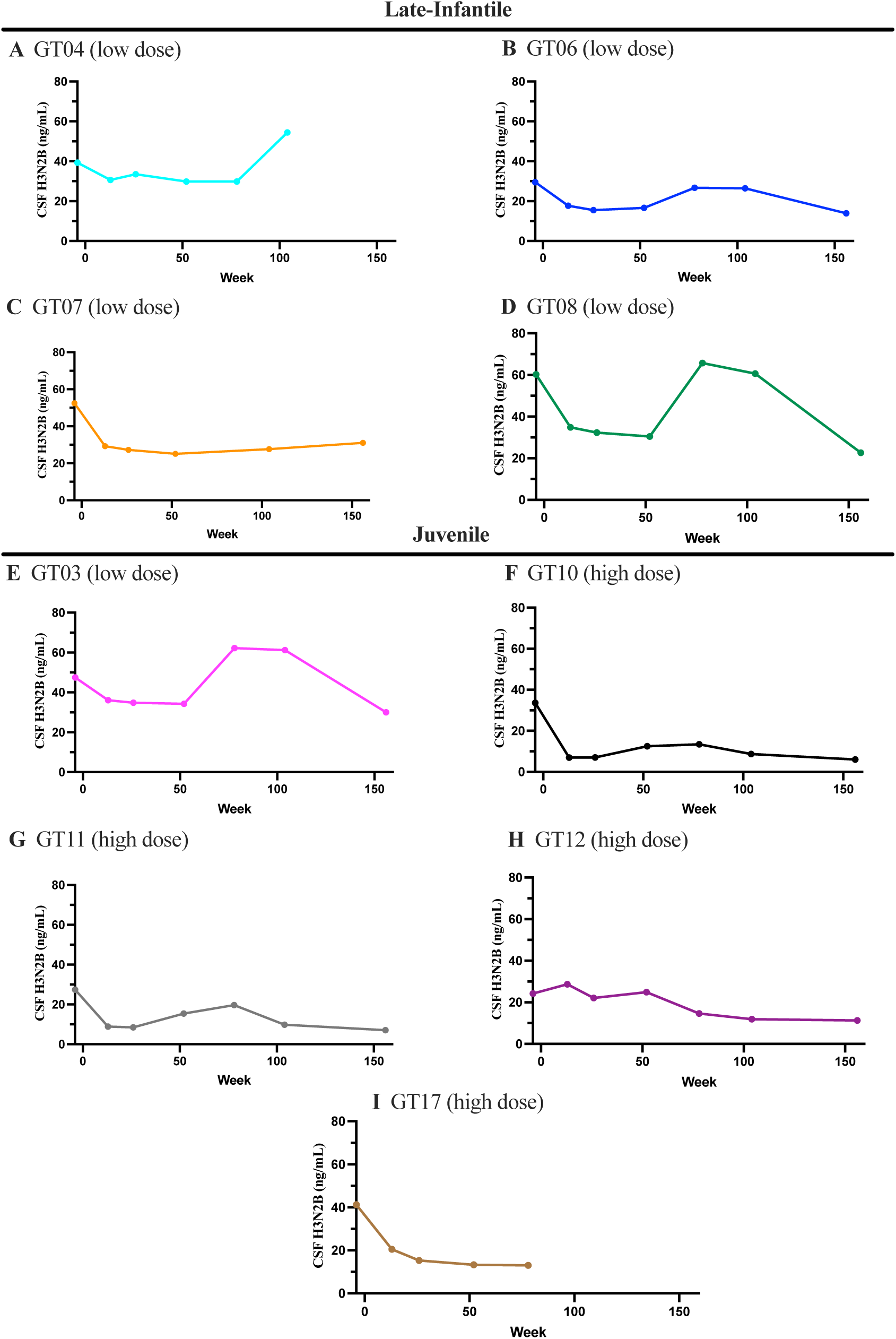
H3N2b Levels in CSF (ng/mL).

**Figure K4.**
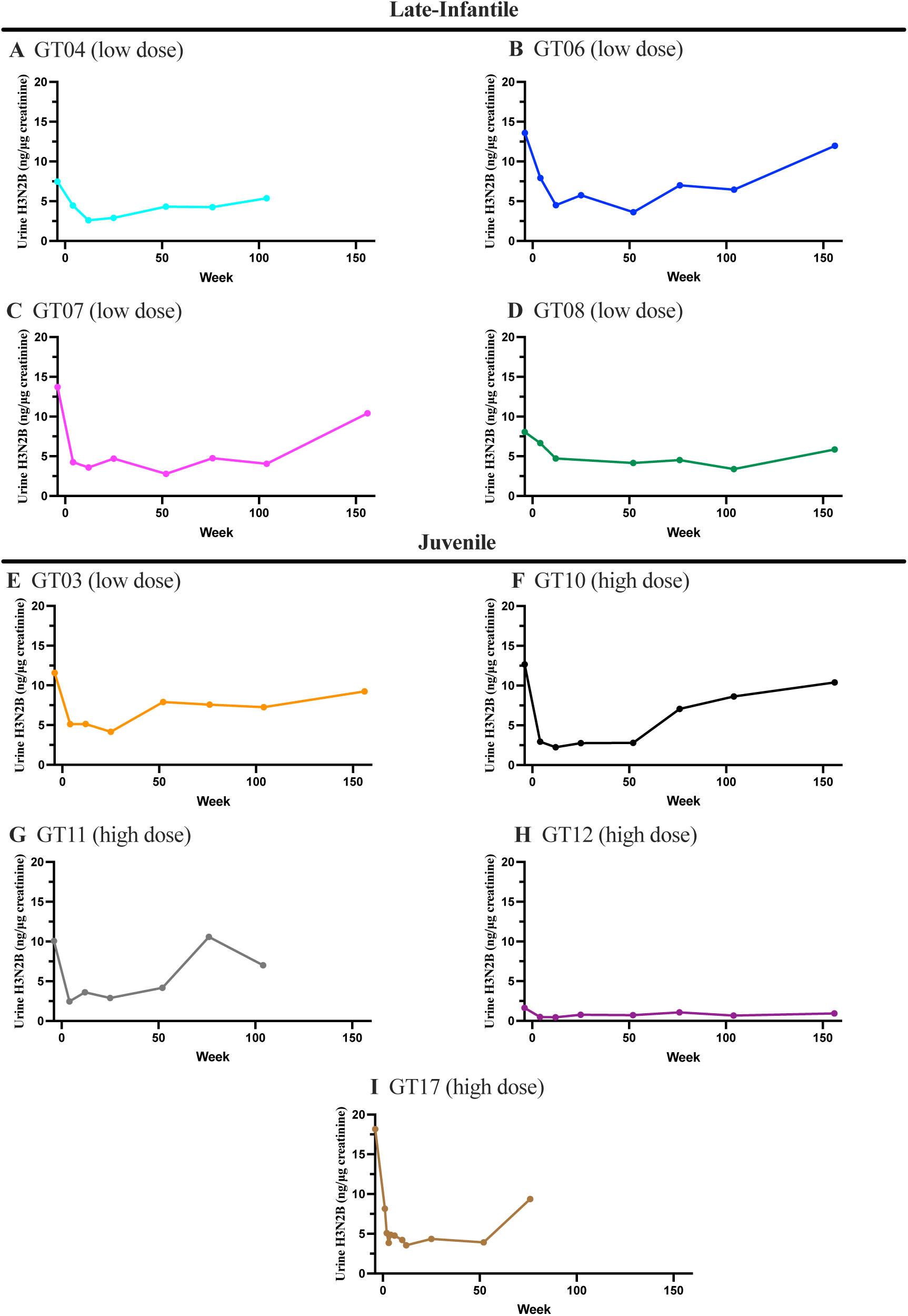
H3N2b Levels in Urine. H3N2b levels were normalized to human creatinine levels (ng/mL creatinine).

**Figure K5.**
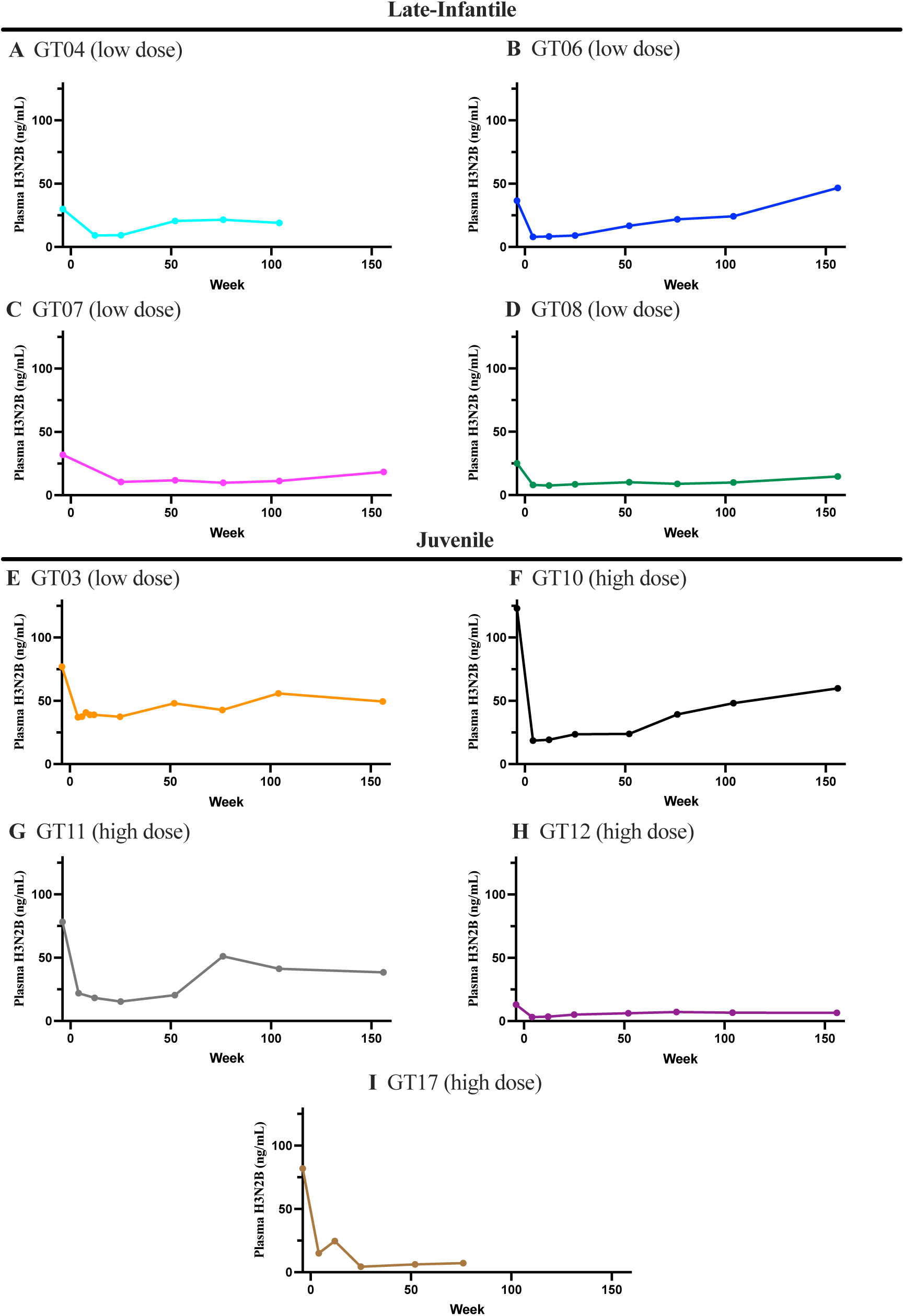
H3N2b Levels in Plasma (ng/mL).

#### Supplement L Clinical Outcome Assessments

##### Vineland Adaptive Behavior Growth Scale Values (GSV)

**Figure L1.**
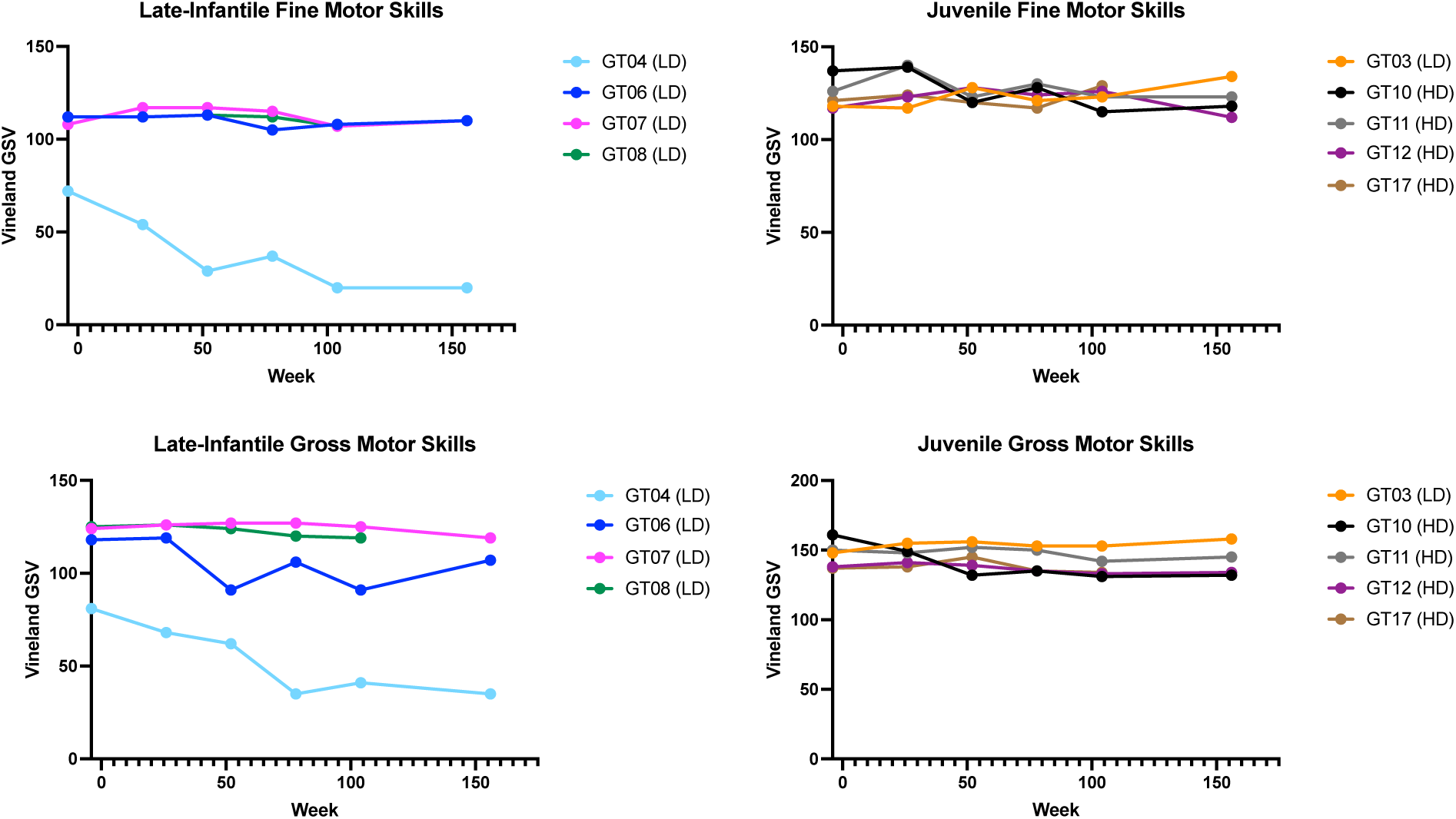
Vineland Adaptive Behavioral Growth Scale Values for fine and gross motor skills. Late-infantile GM1 participants are shown on the left, and juvenile participants are shown on the right with LD corresponding to participants who received low dose, and HD corresponding to high dose administration of AAV9-GLB1. In the late-infantile cohort, GT06, GT07, and GT08 demonstrated stability in both fine and gross motor skills, where GT04 showed a slight decline in both language domains. Juvenile participants showed stability in both fine and gross motor skills.

**Figure L2.**
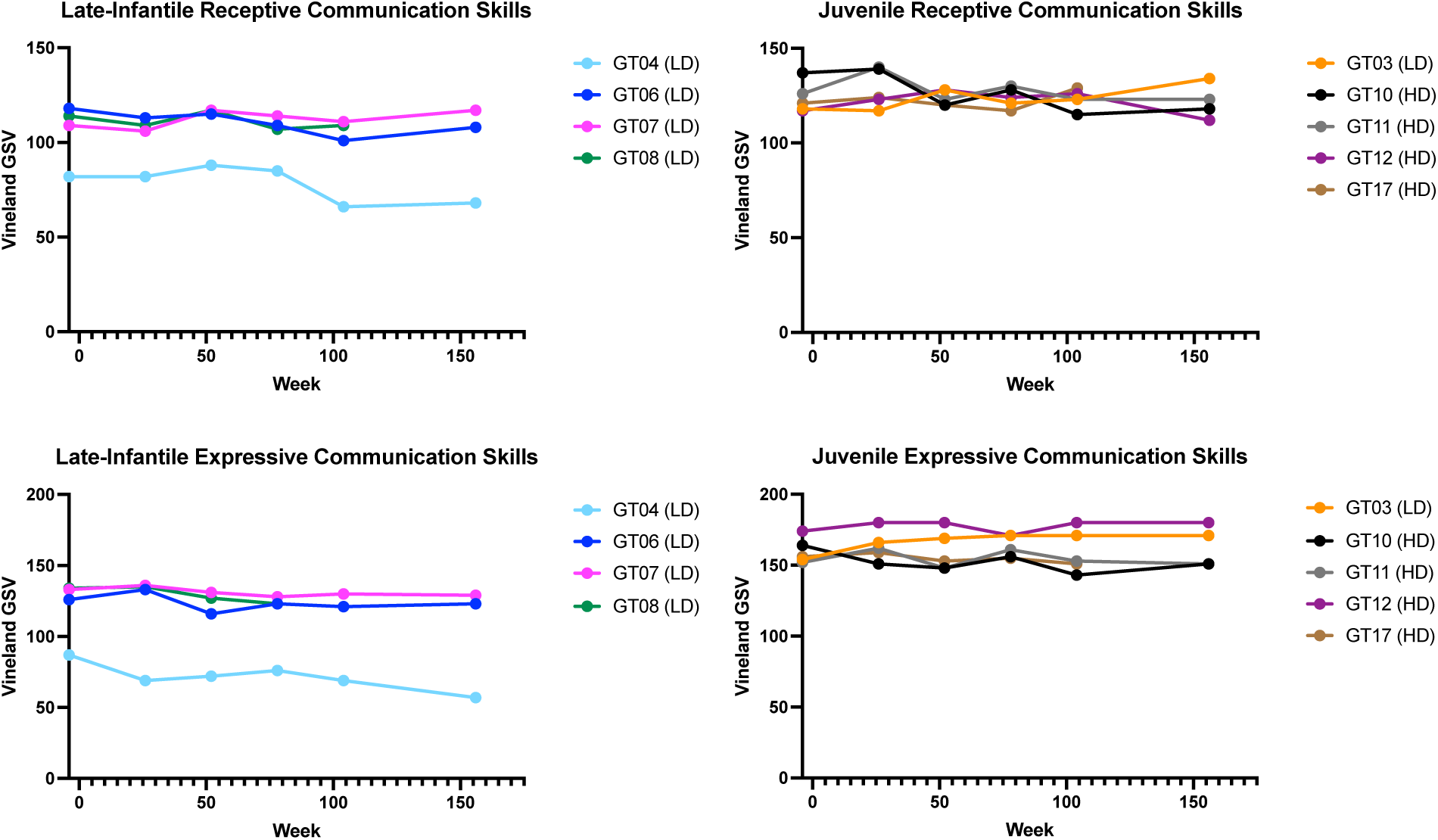
Vineland Adaptive Behavioral Growth Scale Values for Receptive and Expressive Communication skills. Late-infantile GM1 participants are shown on the left, and juvenile participants are shown on the right with LD corresponding to participants who received low dose, and HD corresponding to high dose administration of AAV9-GLB1. In the late-infantile cohort, GT06, GT07, and GT08 demonstrated stability in both expressive and receptive communication, where GT04 showed a slight decline in both communication domains. Juvenile participants showed stability in both expressive and receptive communication skills.

**Figure L3.**
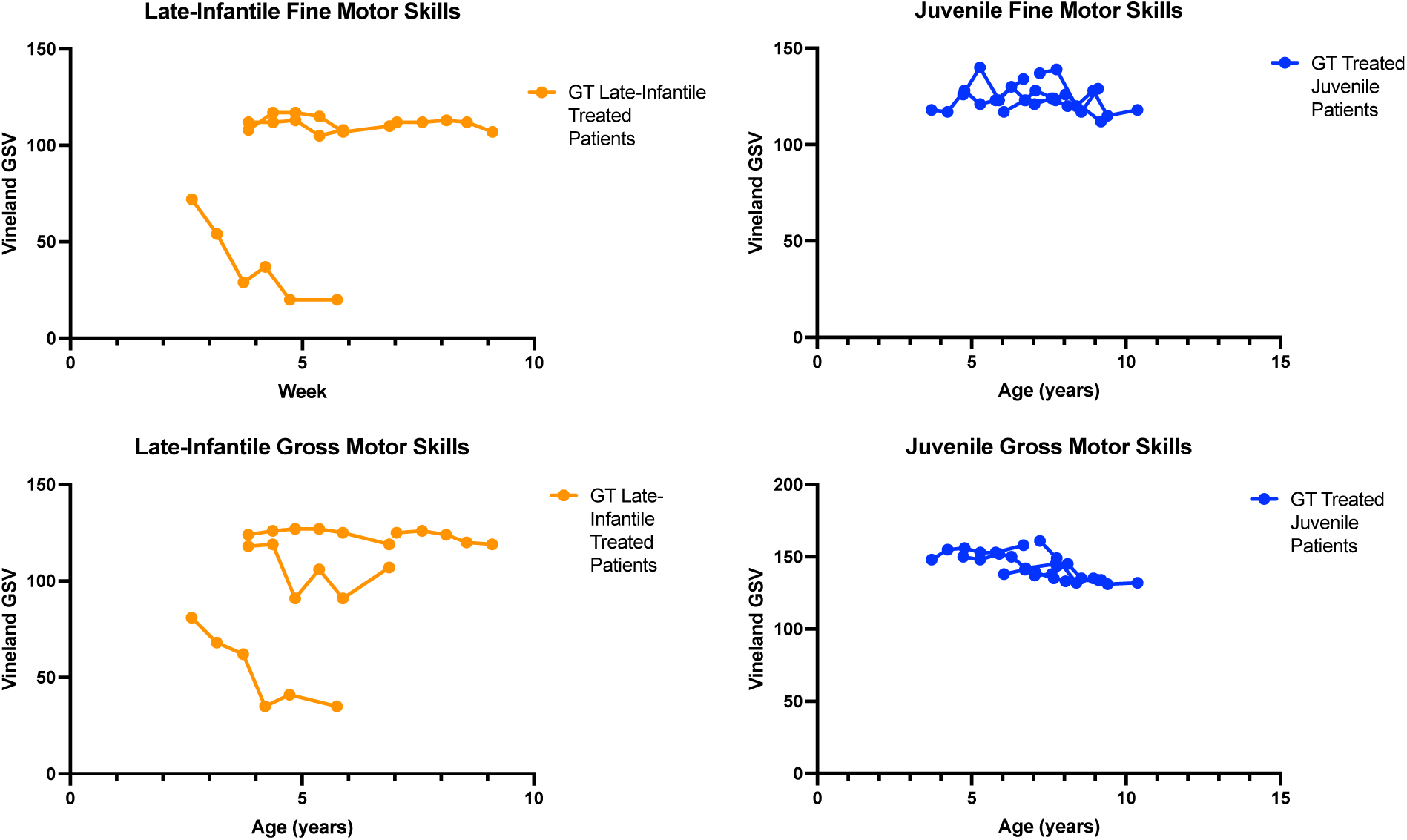
Vineland Adaptive Behavioral Growth Scale Values for fine and gross motor skills shown by patient age. Late-infantile GM1 participants are shown on the left, and juvenile participants are shown on the right with LD corresponding to participants who received low dose, and HD corresponding to high dose administration of AAV9-GLB1. Participant specific designations were redacted per MedArXiv requirements.

**Figure L4.**
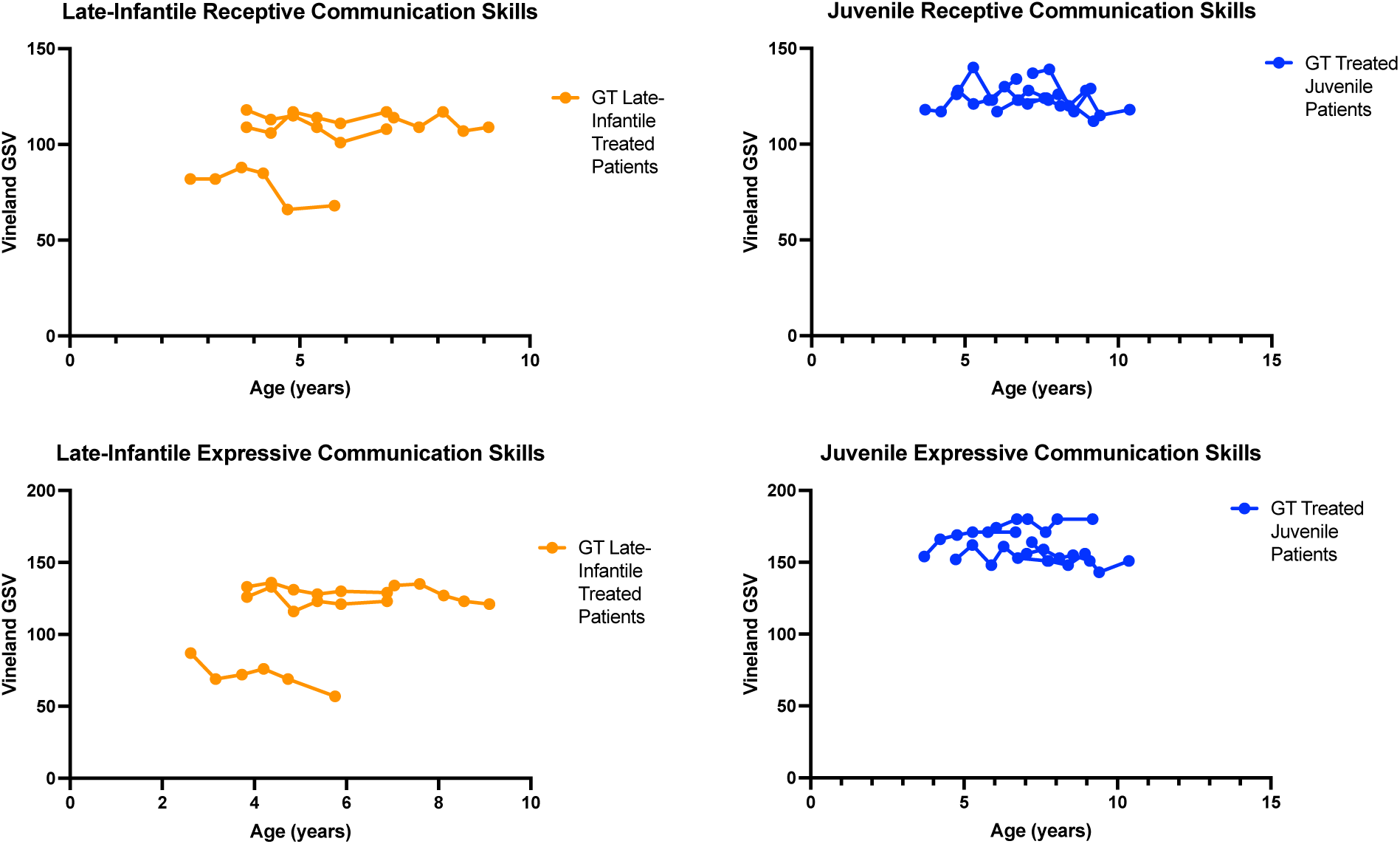
Vineland Adaptive Behavioral Growth Scale Values for Receptive and Expressive Communication skills shown by patient age. Late-infantile GM1 participants are shown on the left, and juvenile participants are shown on the right with LD corresponding to participants who received low dose, and HD corresponding to high dose administration of AAV9-GLB1. Participant specific designations were redacted per MedArXiv requirements.

**Figure L5.**
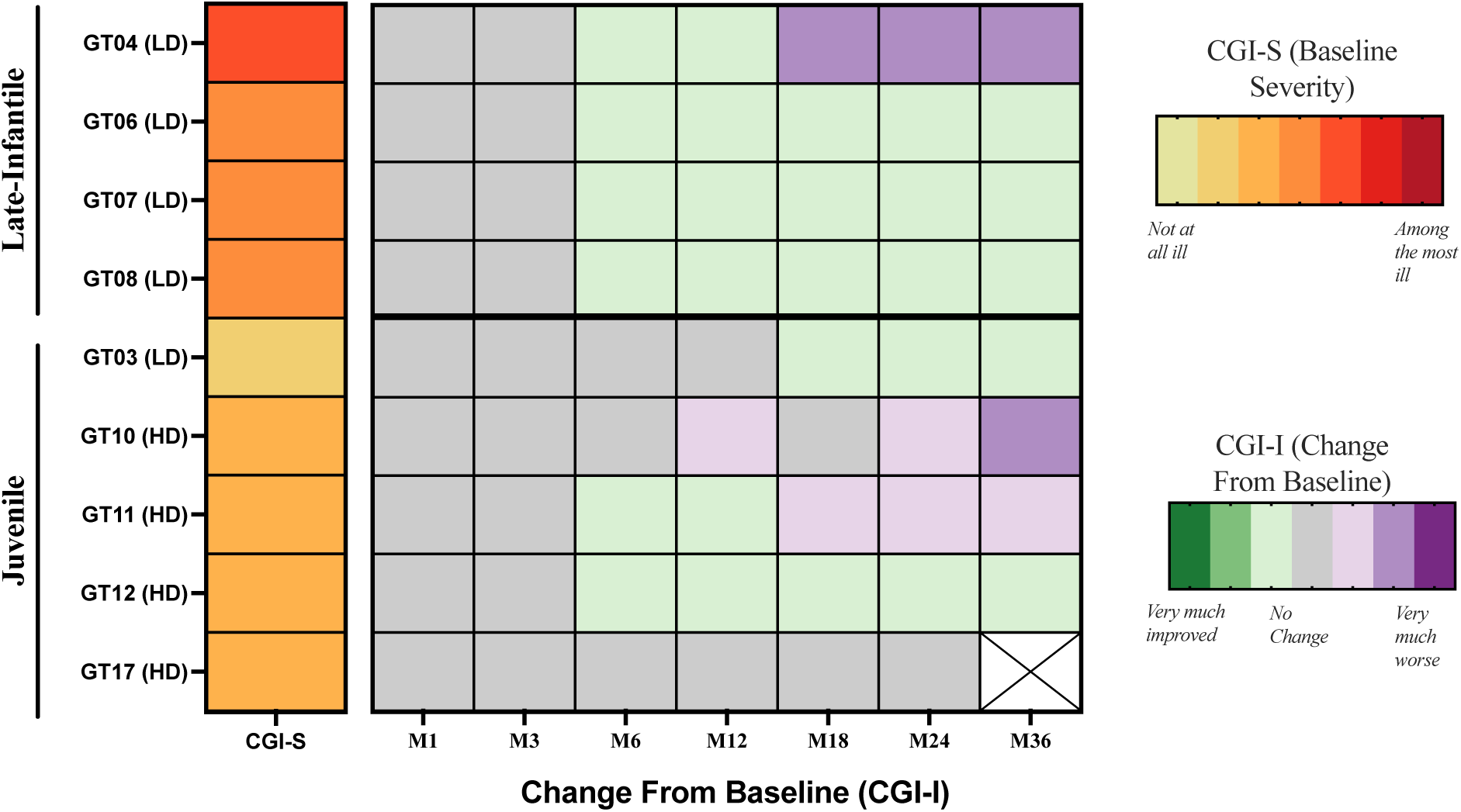
Baseline Clinical Global Impression scales of severity (CGI-S) and Clinical Global Impression of Improvement (CGI-I).

#### Supplement M Neuroimaging Results

##### T1-Weighted Results – Brain Volume

**Figure M1.**
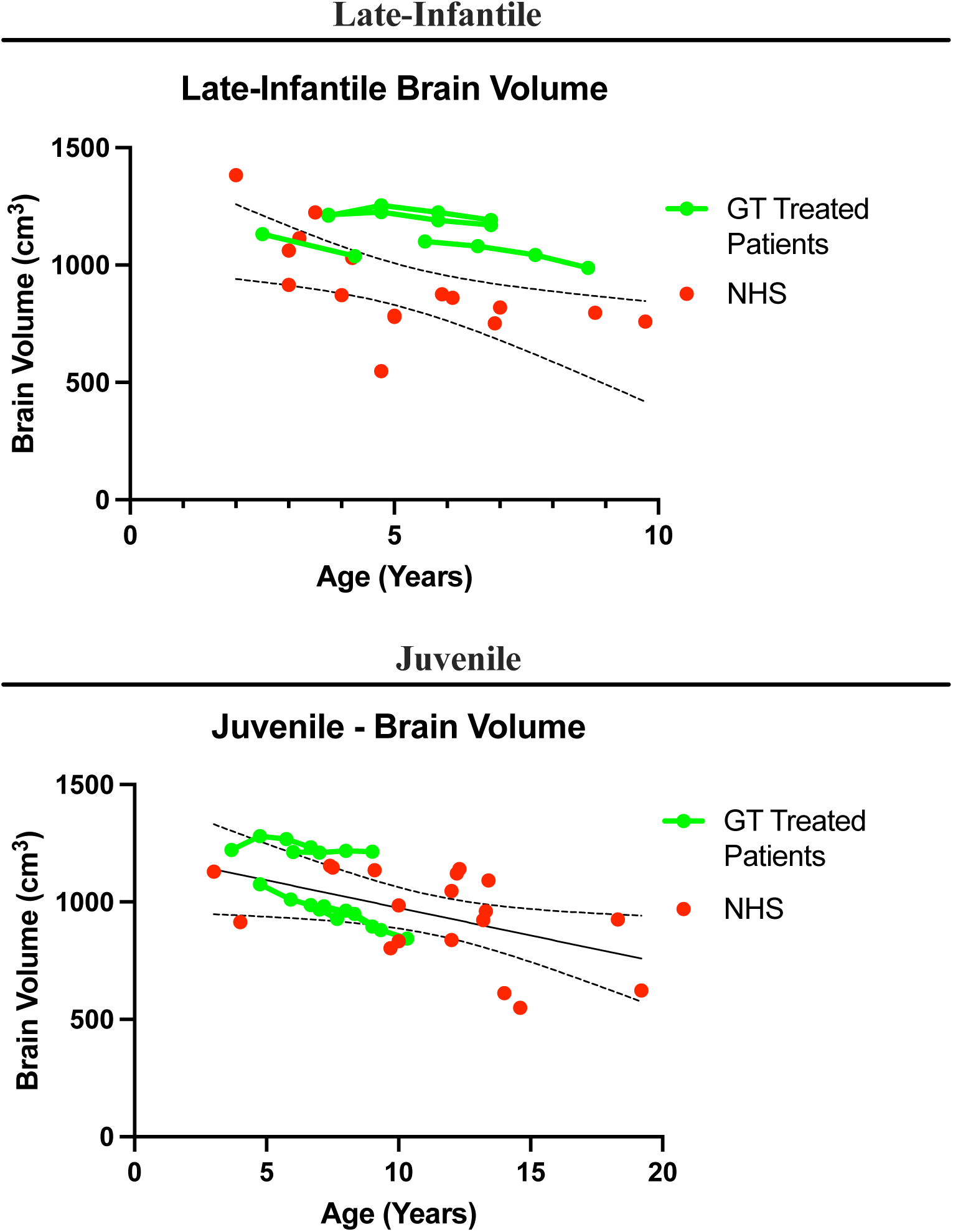
Quantification of cerebral atrophy in treated GM1 participants compared to untreated late-GM1 participants from the NHGRI natural history study (red). A simple linear regression was performed on the untreated natural history data to define the normal trajectory of both the late-infantile (*n* = 11) and juvenile (*n* = 18) diseases (with a 95% CI shown as the dotted line). Late-infantile participants GT07 and GT08 showed improvement in the rate of cerebral atrophy compared to the average change in untreated late-infantile participants. Juvenile participants GT03 and GT12 showed improvement in the rate of cerebral atrophy compared to the average change in untreated juvenile participants. Individual participant graphs were redacted per MedArXiv requirements.

##### T1-Weighted Results – Ventricle Volume

**Figure M2.**
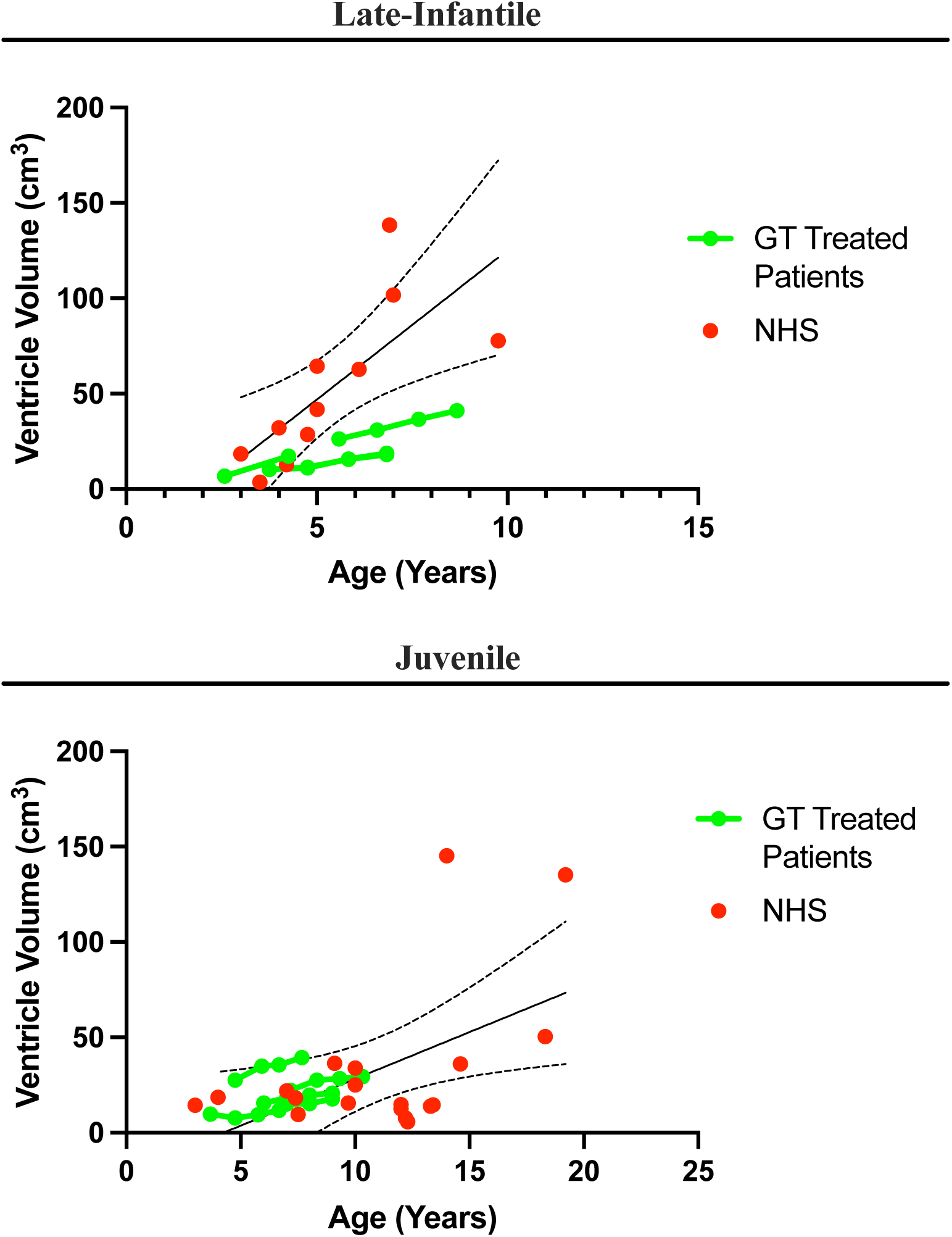
Quantification of ventricle volume in treated GM1 participants compared to untreated GM1 participants from the NHGRI natural history study (red). A simple linear regression was performed on the untreated natural history data to define the normal trajectory of both the late-infantile (*n* = 11) and juvenile (*n* = 18) diseases (with a 95% CI shown as the dotted line). Late-infantile participants GT04, GT06, GT07, and GT08 all showed improvement in the rate of ventricular enlargement compared to the average change in untreated late-infantile participants. Juvenile participants GT03, GT10, GT12, and GT17 all showed improvement in the rate of ventricular enlargement compared to the average change in untreated juvenile participants. Individual participant graphs were redacted per MedArXiv requirements.

##### T1-Weighted Results – Thalamic Volume

**Figure M3.**
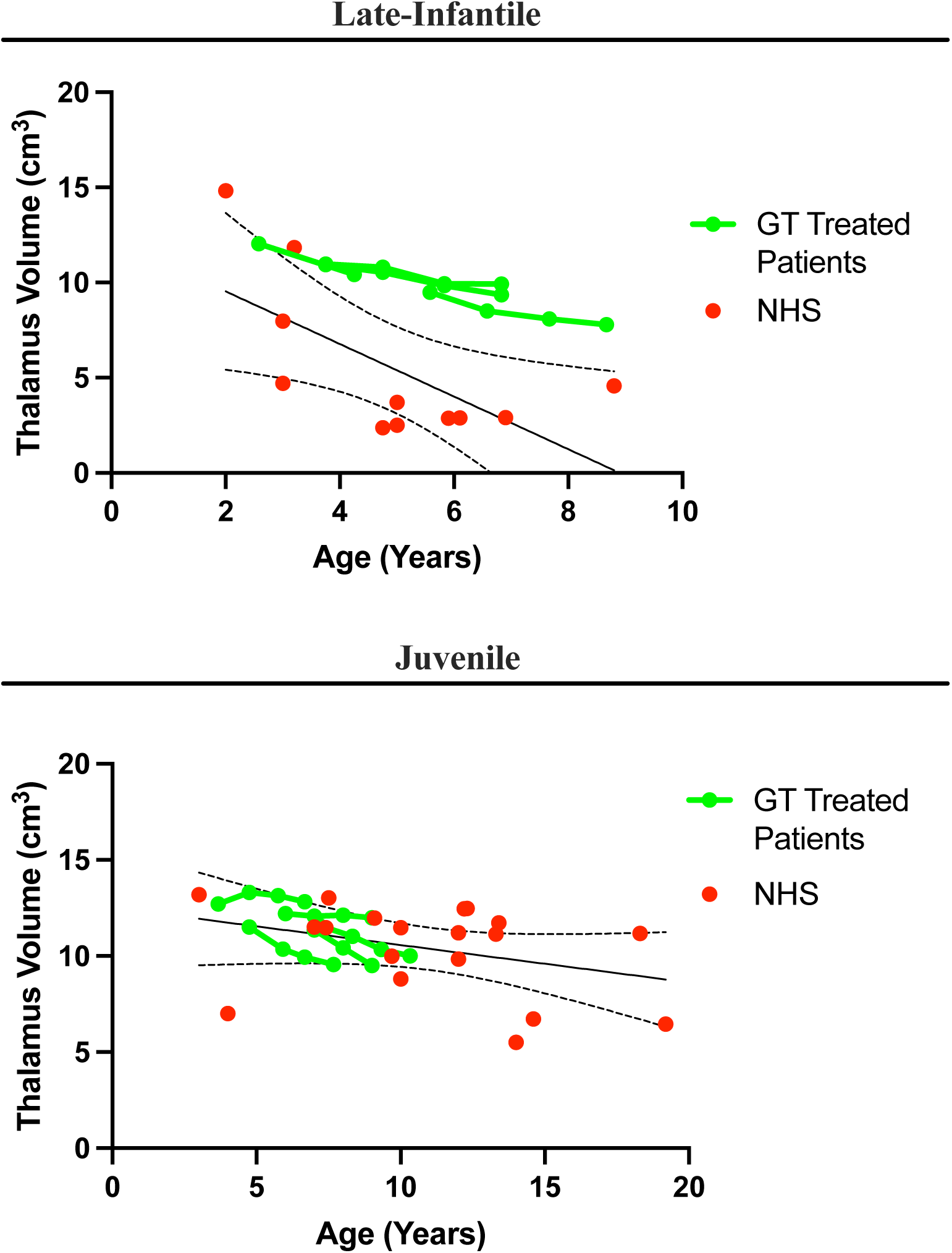
Quantification of thalamic volume in treated GM1 participants compared to untreated GM1 participants from the NHGRI natural history study (red). A simple linear regression was performed on the untreated natural history data to define the normal trajectory of both the late-infantile (*n* = 11) and juvenile (*n* = 18) diseases (with a 95% CI shown as the dotted line). Late-infantile participants GT04, GT06, GT07, and GT08 all showed improvement in the rate of thalamic atrophy compared to the average change in untreated late-infantile participants. Juvenile participants GT03 and GT12 showed improvement in the rate of thalamic atrophy compared to the average change in untreated juvenile participants. Individual participant graphs were redacted per MedArXiv requirements.

##### DT Results – Net Fiber Tract Metrics

**Figure M4.**
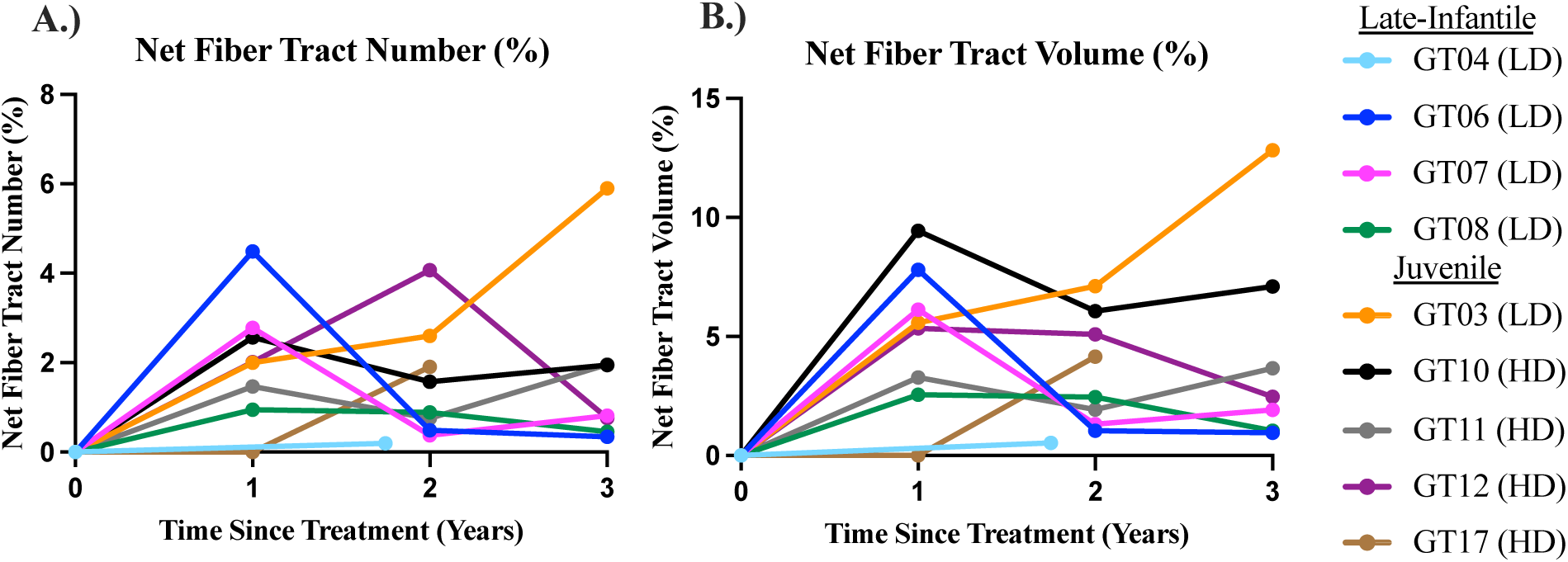
Quantification of Differential Tractography. A.) Net fiber number changes as a percentage relative to baseline tractography, individualized by patient. B.) Net fiber volume changes as a percentage relative to baseline tractography, individualized by patient.

##### MRS – LCSO – *N*-Acetylaspartete + *N*-Acetylaspartyl glutamate (NAA)

**Figure M5.**
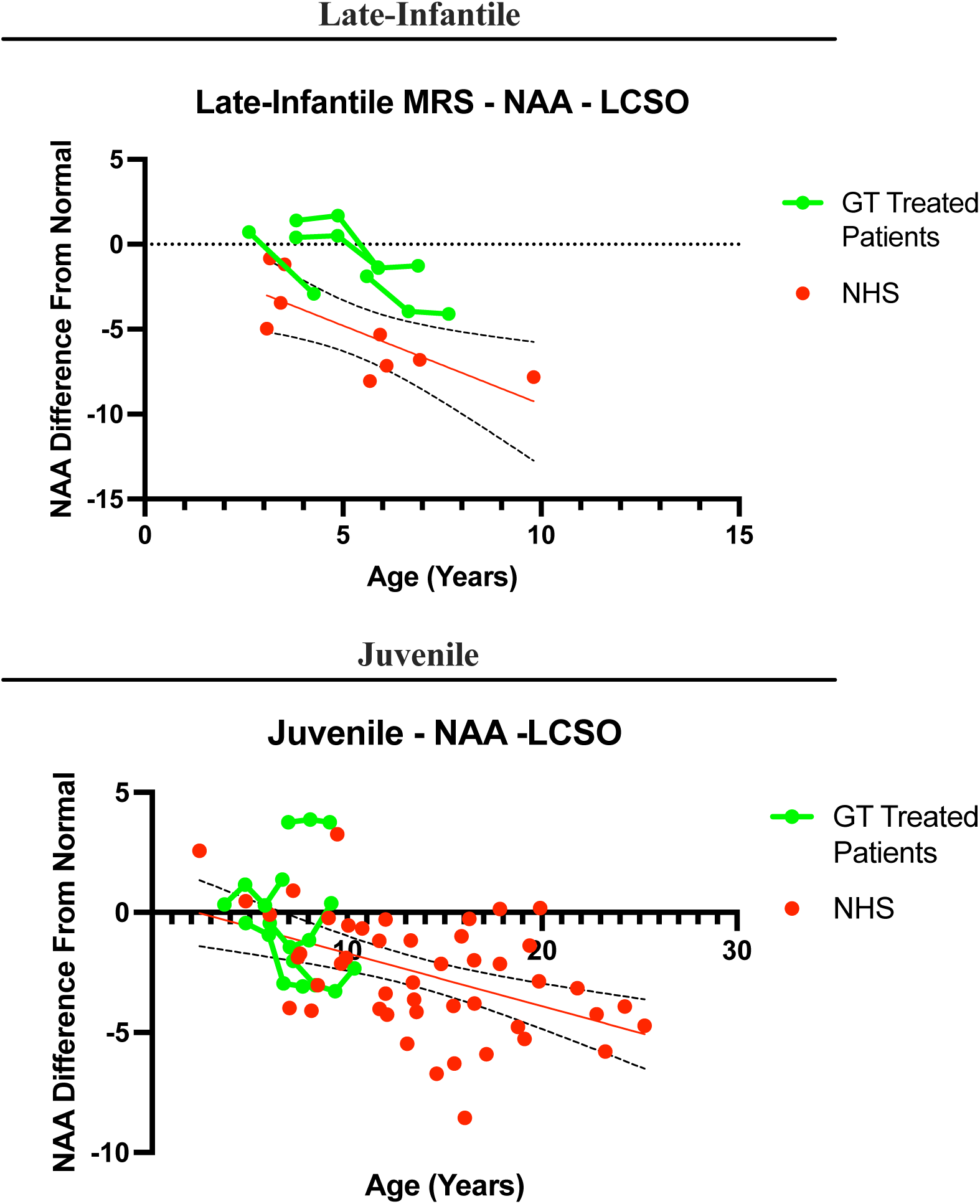
Quantification of *N*-Acetylaspartate + *N*-Acetylaspartyl glutamate (NAA) in the left centrum semiovale (LCSO) in treated GM1 participants compared to untreated GM1 participants from the NHGRI natural history study (red). A simple linear regression was performed on the untreated natural history data to define the normal trajectory of both the late-infantile (*n* = 9) and juvenile (*n* = 17) diseases (with a 95% CI shown as the dotted line). Late-infantile participants GT07 and GT08 showed stabilization of the NAA decreases associated with late-infantile GM1 disease progression in the year following gene transfer. GT08 showed a diminished rate of NAA compared to the untreated GM1 participants in the second year following gene transfer. Juvenile participants GT03 GT10, and GT12 all showed stabilization of the NAA decreases associated with juvenile GM1 disease progression. Individual participant graphs were redacted per MedArXiv requirements.

## References

1. Nicoli ER, Annunziata I, d’Azzo A, Platt FM, Tifft CJ, Stepien KM. GM1 Gangliosidosis-A Mini-Review. Front Genet. 2021;12:734878. Published 2021 Sep 3. doi:10.3389/fgene.2021.734878.

2. Feng Y, Huang Y, Zhao X, et al. Clinical and molecular characteristics of 11 Chinese probands with GM1 gangliosidosis. Metab Brain Dis. 2018;33(6):2051–2057. doi:10.1007/s11011-018-0315-2.

3. Karimzadeh P, Naderi S, Modarresi F, et al. Case reports of juvenile GM1 gangliosidosisis type II caused by mutation in GLB1 gene. BMC Med Genet. 2017;18(1):73. Published 2017 Jul 17. doi:10.1186/s12881-017-0417-4.

4. King KE, Kim S, Whitley CB, Jarnes-Utz JR. The juvenile gangliosidoses: A timeline of clinical change. Mol Genet Metab Rep. 2020;25:100676. Published 2020 Nov 14. doi:10.1016/j.ymgmr.2020.100676.

5. Kolstad J, Zoppo C, Johnston JM, et al. Natural history progression of MRI brain volumetrics in type II late-infantile and juvenile GM1 gangliosidosis patients. Mol Genet Metab. 2025;144(3):109025. doi:10.1016/j.ymgme.2025.109025.

6. Nestrasil I, Ahmed A, Utz JM, Rudser K, Whitley CB, Jarnes-Utz JR. Distinct progression patterns of brain disease in infantile and juvenile gangliosidoses: Volumetric quantitative MRI study. Mol Genet Metab. 2018;123(2):97–104. doi:10.1016/j.ymgme.2017.12.432

7. Laur D, Pichard S, Bekri S, et al. Natural history of GM1 gangliosidosis-Retrospective cohort study of 61 French patients from 1998 to 2019. J Inherit Metab Dis. 2023;46(5):972–981. doi:10.1002/jimd.12646.

8. Gross AL, Gray-Edwards HL, Bebout CN, et al. Intravenous delivery of adeno-associated viral gene therapy in feline GM1 gangliosidosis. Brain. 2022;145(2):655–669. doi:10.1093/brain/awab309.

9. Gray-Edwards HL, Maguire AS, Salibi N, et al. 7T MRI Predicts Amelioration of Neurodegeneration in the Brain after AAV Gene Therapy. Mol Ther Methods Clin Dev. 2019;17:258–270. Published 2019 Dec 24. doi:10.1016/j.omtm.2019.11.023.

10. Kell P, Sidhu R, Qian M, et al. A pentasaccharide for monitoring pharmacodynamic response to gene therapy in GM1 gangliosidosis. EBioMedicine. 2023;92:104627. doi:10.1016/j.ebiom.2023.104627.

11. D’Souza P, Farmer C, Johnston JM, et al. GM1 gangliosidosis type II: Results of a 10-year prospective study. Genet Med. 2024;26(7):101144. doi:10.1016/j.gim.2024.101144.

12. Lewis CJ, Johnston JM, Zaragoza Domingo S, et al. Retrospective assessment of clinical global impression of severity and change in GM1 gangliosidosis: a tool to score natural history data in rare disease cohorts. Orphanet J Rare Dis. 2025;20(1):125. Published 2025 Mar 14. doi:10.1186/s13023-025-03614-6.

13. Lewis CJ, Vardar Z, Kühn AL, et al. Differential tractography: an imaging marker for tissue degeneration in neurodegenerative diseases. Brain Communications, 2025, fcaf198, 10.1093/braincomms/fcaf198.

14. Galletta F, Cucinotta U, Marseglia L, et al. Hemophagocytic lymphohistiocytosis following gene replacement therapy in a child with type 1 spinal muscular atrophy. J Clin Pharm Ther. 2022;47(9):1478–1481. doi:10.1111/jcpt.13733.

15. Neurogene. Neurogene Provides Update on NGN-401 Gene Therapy Clinical Trial for Rett Syndrome. November 18, 2024 (https://ir.neurogene.com/news-releases/news-release-details/neurogene-provides-update-ngn-401-gene-therapy-clinical-trial).

16. Guillou J, de Pellegars A, Porcheret F, et al. Fatal thrombotic microangiopathy case following adeno-associated viral SMN gene therapy. Blood Adv. 2022;6(14):4266–4270. doi:10.1182/bloodadvances.2021006419.

17. Rocket Pharmaceuticals. Rocket Pharmaceuticals Provides Update on Phase 2 Clinical Trial of RP-A501 for Danon Disease. May 27, 2025 (https://ir.rocketpharma.com/news-releases/news-release-details/rocket-pharmaceuticals-provides-update-phase-2-clinical-trial-rp).

18. Lek A, Wong B, Keeler A, et al. Death after High-Dose rAAV9 Gene Therapy in a Patient with Duchenne’s Muscular Dystrophy. N Engl J Med. 2023;389(13):1203–1210. doi:10.1056/NEJMoa2307798.

19. Hordeaux J, Lamontagne RJ, Bandyopadhyay S, Bell P, Wilson JM, Flotte TR. Lung endothelial transduction in a patient that succumbed to acute respiratory distress syndrome following high-dose rAAV9 gene therapy. Mol Ther. 2025;33(6):2339–2342. doi:10.1016/j.ymthe.2025.05.017.

20. Ma X, Zhuang L, Ma W, et al. AAV9-Mediated Gene Therapy for Infantile-Onset Pompe’s Disease. N Engl J Med. 2025;392(24):2438–2446. doi:10.1056/NEJMoa2407766.

21. D’Antiga L, Beuers U, Ronzitti G, et al. Gene Therapy in Patients with the Crigler-Najjar Syndrome. N Engl J Med. 2023;389(7):620–631. doi:10.1056/NEJMoa2214084.

22. Flotte TR, Cataltepe O, Puri A, et al. AAV gene therapy for Tay-Sachs disease. Nat Med. 2022;28(2):251–259. doi:10.1038/s41591-021-01664-4.

23. Greenberg B, Taylor M, Adler E, et al. Phase 1 Study of AAV9.LAMP2B Gene Therapy in Danon Disease. N Engl J Med. 2025;392(10):972–983. doi:10.1056/NEJMoa2412392.

24. Hwu WL, Muramatsu S, Tseng SH, et al. Gene therapy for aromatic L-amino acid decarboxylase deficiency. Sci Transl Med. 2012;4(134):134ra61. doi:10.1126/scitranslmed.3003640.

25. Meliani A, Boisgerault F, Hardet R, et al. Antigen-selective modulation of AAV immunogenicity with tolerogenic rapamycin nanoparticles enables successful vector re-administration. Nat Commun. 2018;9(1):4098. Published 2018 Oct 5. doi:10.1038/s41467-018-06621-3.

26. Sessa M, Lorioli L, Fumagalli F, et al. Lentiviral haemopoietic stem-cell gene therapy in early-onset metachromatic leukodystrophy: an ad-hoc analysis of a non-randomised, open-label, phase 1/2 trial. Lancet. 2016;388(10043):476–487. doi:10.1016/S0140-6736(16)30374-9.

27. Merkel SF, Andrews AM, Lutton EM, et al. Trafficking of adeno-associated virus vectors across a model of the blood-brain barrier; a comparative study of transcytosis and transduction using primary human brain endothelial cells. J Neurochem. 2017;140(2):216–230. doi:10.1111/jnc.13861.

28. Weismann, C. M., Ferreira, J., Keeler, A. M., Su, Q., Qui, L., Shaffer, S. A., Xu, Z., Gao, G., & Sena-Esteves, M. (2015). Systemic AAV9 gene transfer in adult GM1 gangliosidosis mice reduces lysosomal storage in CNS and extends lifespan. Human molecular genetics, 24(15), 4353–4364. 10.1093/hmg/ddv168.

29. McCurdy VJ, Johnson AK, Gray-Edwards HL, et al. Sustained normalization of neurological disease after intracranial gene therapy in a feline model. Sci Transl Med. 2014;6(231):231ra48. doi:10.1126/scitranslmed.3007733.

30. Bharucha-Goebel DX, Todd JJ, Saade D, et al. Intrathecal Gene Therapy for Giant Axonal Neuropathy. N Engl J Med. 2024;390(12):1092–1104. doi:10.1056/NEJMoa2307952.

31. Winston G, Kharas N, Svenningsson P, Jha A, Kaplitt GK. Gene therapy for Parkinson’s disease: trials and technical advances. Lancet Neurol. 2025;24(6):548–556. 10.1016/S1474-4422(25)00125-5.

32. Huang Q, Chan KY, Wu J, et al. An AAV capsid reprogrammed to bind human transferrin receptor mediates brain-wide gene delivery. Science. 2024;384(6701):1220–1227. doi:10.1126/science.adm8386.

## Supplementary References

1. D’Souza P, Farmer C, Johnston JM, et al. GM1 gangliosidosis type II: Results of a 10-year prospective study. Genet Med. 2024;26(7):101144. doi:10.1016/j.gim.2024.101144.

2. Patterson, Marc C. “Gangliosidoses.” Handbook of clinical neurology 113 (2013): 1707–1708.

3. Regier, Debra S., Cynthia J. Tifft, and Caroline E. Rothermel. “GLB1-related disorders.” (2021).

4. Flotte TR, Cataltepe O, Puri A, et al. AAV gene therapy for Tay-Sachs disease. Nat Med. 2022;28(2):251–259. doi:10.1038/s41591-021-01664-4.

5. Kell P, Sidhu R, Qian M, et al. A pentasaccharide for monitoring pharmacodynamic response to gene therapy in GM1 gangliosidosis. EBioMedicine. 2023;92:104627. doi:10.1016/j.ebiom.2023.104627.

6. Leonard M, Dunn J, Smith G. A clinical biomarker assay for the quantification of d3-creatinine and creatinine using LC-MS/MS. Bioanalysis. 2014;6(6):745–759. doi:10.4155/bio.13.323.

7. Leon-Astudillo C, Coleman K, Salabarria SM, et al. Quantification and comparison of anti-AAV9 and anti-AAVrh74 antibodies in plasma and human milk: Implications for AAV-based gene therapy candidacy. J Neuromuscul Dis. Published online April 10, 2025. doi:10.1177/22143602251324857

8. Sparrow SS, Cicchetti DV, Saulnier CA. Vineland-3: Vineland adaptive behavior scales. PsychCorp; 2016.

9. Farmer C, Ludwig, NN, Thurm A. A tutorial on person ability scores for the intellectual and developmental disabilities clinician. International Journal of Developmental Disabilities, (2025)1–10. 10.1080/20473869.2025.2486428.

10. Farmer C, Thurm A, Troy JD, Kaat AJ. Comparing ability and norm-referenced scores as clinical trial outcomes for neurodevelopmental disabilities: a simulation study. J Neurodev Disord. 2023;15(1):4. Published 2023 Jan 17. doi:10.1186/s11689-022-09474-6.

11. Bates D, Mächler M, Bolker B, Walker S. Fitting linear mixed-effects models using lme4. arXiv preprint arXiv:14065823. 2014.

12. Kuznetsova A, Brockhoff PB, Christensen RH. lmerTest package: tests in linear mixed effects models. Journal of statistical software. 2017;82:1–26. in R version 4.4.2.

13. Guy W. ECDEU assessment manual for psychopharmacology. US Department of Health, Education, and Welfare, Public Health Service, 1976.

14. Busner J, Targum SD. The clinical global impressions scale: applying a research tool in clinical practice. Psychiatry (Edgmont). 2007;4(7):28–37.

15. Lewis CJ, Johnston JM, Zaragoza Domingo S, et al. Retrospective assessment of clinical global impression of severity and change in GM1 gangliosidosis: a tool to score natural history data in rare disease cohorts. Orphanet J Rare Dis. 2025;20(1):125. Published 2025 Mar 14. doi:10.1186/s13023-025-03614-6.

16. National Human Genome Research Institute. Natural History of Glycosphingolipid Storage Disorders and Glycoprotein Disorders. ClinicalTrials.gov identifier: NCT00029965. https://clinicaltrials.gov/study/NCT00029965.

17. National Human Genome Research Institute. A Phase 1/2 Study of Intravenous Gene Transfer with an AAV9 Vector Expressing Human Beta-galactosidase in Type I and Type II GM1 Gangliosidosis. ClinicalTrials.gov identifier: NCT03952637. https://clinicaltrials.gov/study/NCT03952637.

18. Kolstad J, Zoppo C, Johnston JM, et al. Natural history progression of MRI brain volumetrics in type II late-infantile and juvenile GM1 gangliosidosis patients. Mol Genet Metab. 2025;144(3):109025. doi:10.1016/j.ymgme.2025.109025.

19. Lewis CJ, Vardar Z, Kühn AL, et al. Differential tractography: an imaging marker for tissue degeneration in neurodegenerative diseases. Brain Communications, 2025, fcaf198, 10.1093/braincomms/fcaf198.

20. Li X, Morgan PS, Ashburner J, Smith J, Rorden C. The first step for neuroimaging data analysis: DICOM to NIfTI conversion. J Neurosci Methods. 2016;264:47–56. doi:10.1016/j.jneumeth.2016.03.001.

21. Manjón JV, Romero JE, Vivo-Hernando R, et al. vol2Brain: A New Online Pipeline for Whole Brain MRI Analysis. Front Neuroinform. 2022;16:862805. Published 2022 May 24. doi:10.3389/fninf.2022.862805.

22. Lewis CJ, Johnston JM, D’Souza P, et al. A Case for Automated Segmentation of MRI Data in Neurodegenerative Diseases: Type II GM1 Gangliosidosis. NeuroSci. 2025;6(2):31. Published 2025 Apr 3. doi:10.3390/neurosci6020031.

23. Yeh FC, Zaydan IM, Suski VR, et al. Differential tractography as a track-based biomarker for neuronal injury. Neuroimage. 2019;202:116131. doi:10.1016/j.neuroimage.2019.116131.

24. Tournier JD, Smith R, Raffelt D, et al. MRtrix3: A fast, flexible and open software framework for medical image processing and visualisation. Neuroimage. 2019;202:116137. doi:10.1016/j.neuroimage.2019.116137.

25. Andersson JLR, Sotiropoulos SN. An integrated approach to correction for off-resonance effects and subject movement in diffusion MR imaging. Neuroimage. 2016;125:1063–1078. doi:10.1016/j.neuroimage.2015.10.019.

26. Smith SM, Jenkinson M, Woolrich MW, et al. Advances in functional and structural MR image analysis and implementation as FSL. Neuroimage. 2004;23 Suppl 1:S208–S219. doi:10.1016/j.neuroimage.2004.07.051.

27. Dhollander T, Raffelt D, Connelly A. Unsupervised 3-tissue response function estimation from single-shell or multi-shell diffusion MR data without a co-registered T1 image. ISMRM Workshop on Breaking the Barriers of Diffusion MRI, 2016, 5.

28. Jenkinson M, Beckmann CF, Behrens TE, Woolrich MW, Smith SM. FSL. Neuroimage. 2012;62(2):782–790. doi:10.1016/j.neuroimage.2011.09.015.

29. Woolrich MW, Jbabdi S, Patenaude B, et al. Bayesian analysis of neuroimaging data in FSL. Neuroimage. 2009;45(1 Suppl):S173–S186. doi:10.1016/j.neuroimage.2008.10.055.

30. Yeh FC, Wedeen VJ, Tseng WY. Generalized q-sampling imaging. IEEE Trans Med Imaging. 2010;29(9):1626–1635. doi:10.1109/TMI.2010.2045126.

31. Provencher SW. Estimation of metabolite concentrations from localized in vivo proton NMR spectra. Magnetic resonance in medicine. 1993;30(6):672–679.

32. Ernst T, Kreis R, Ross B. Absolute quantitation of water and metabolites in the human brain. I. Compartments and water. Journal of magnetic resonance, Series B. 1993;102(1):1–8.

33. Srinivasan R, Sailasuta N, Hurd R, Nelson S, Pelletier D. Evidence of elevated glutamate in multiple sclerosis using magnetic resonance spectroscopy at 3 T. Brain. 2005;128(5):1016–1025.

34. Baker EH, Levin SW, Zhang Z, Mukherjee AB. Evaluation of disease progression in INCL by MR spectroscopy. Annals of clinical and translational neurology. 2015;2(8):797–809.

35. Baker EH, Basso G, Barker PB, Smith MA, Bonekamp D, Horská A. Regional apparent metabolite concentrations in young adult brain measured by 1H MR spectroscopy at 3 Tesla. Journal of Magnetic Resonance Imaging: An Official Journal of the International Society for Magnetic Resonance in Medicine. 2008;27(3):489–499.

36. Horska A, Calhoun V, Bradshaw D, Barker P. Rapid method for correction of CSF partial volume in quantitative proton MR spectroscopic imaging. Magnetic Resonance in Medicine: An Official Journal of the International Society for Magnetic Resonance in Medicine. 2002;48(3):555–558.

37. Izumi T, Ogawa T, Koizumi H, Fukuyama Y. Normal developmental profiles of CSF gangliotetraose-series gangliosides from neonatal period to adolescence. Pediatr Neurol. 1993;9(4):297–300. doi:10.1016/0887-8994(93)90067-m.

